# Evaluating the performance of polygenic indices of neuropsychiatric conditions and brain endophenotypes in four UK population samples

**DOI:** 10.64898/2026.07.07.26357467

**Authors:** Anna Dearman, Pascal Vrtička, Jamie Moore, Meena Kumari, Leonard Schalkwyk

## Abstract

Neuropsychiatric polygenic indices (NPGIs) are used as genetic predictors of poor mental health. However, NPGIs are also associated with environmental factors which could affect mental health in adulthood, including the rearing environment. Hence, their “genetic” effects are both direct and environmentally mediated. There is a need to identify alternative genetic predictors without environmental signal. Endophenotype-based polygenic indices (EPGIs) trained on brain structure and function are under-studied alternatives which, due to their relative biological proximity, may exhibit associations with mental health outcomes which are less environmentally mediated than those of NPGIs. Using four representative UK samples (*Understanding Society*; UKHLS, NCDS, BCS70 and MCS) we employ sex-stratified path models to estimate the direct and environmentally mediated effects of eleven NPGIs and 30 EPGIs on adult mental health, focussing on the rearing environment. The depression NPGI is consistently associated with mental health symptoms across most sex-stratified sub-samples (best meta-analysis β = 0.091, p 0.001) but demonstrates 1.6 - 24.5% environmental mediation. Seven other NPGIs and three EPGIs show sample- and sex-specific associations with mental health symptoms. NPGIs for attention deficit hyperactivity disorder, depression and substance use disorder are robustly associated with measures of the rearing environment, which in turn are frequently associated with mental health symptoms. Sensitivity analyses find that NPGI associations vary substantially depending on who is included in the sample. In conclusion, the rearing environment likely mediates a substantial portion of NPGIs’ so-called “genetic” effects on mental health symptoms, but EPGIs are not currently powerful enough to replace them.

## Introduction

Mental health conditions constitute a substantial individual and societal burden, contributing an estimated 14.6% of years lived with disability globally (1), although this figure may be an underestimation (2). In England, the prevalence of common mental disorders - such as anxiety and depression - amongst adults has been estimated at one in six (3). Mental health conditions are multifactorial - the average heritability is estimated at 46% (4), with major depression estimated at around 31-42% (5), hence both genetic and environmental factors play a substantial role in their aetiology. Genetic liability towards a mental health outcome is often measured using polygenic indices (PGIs) a.k.a. polygenic (risk) scores, which are usually trained on neuropsychiatric conditions (neuropsychiatric PGIs; NPGIs). NPGIs have been associated with sub-clinical symptom measures including depressive symptoms (6), thought problems (7) and externalizing behaviour (8), and with intermediate phenotypes (endophenotypes (9)) such as structural (10) and functional (11) brain measures. Efforts to understand how mental health is influenced by the interplay of genetic and environmental factors include a growing literature on gene-environment interaction (GxE) which uses statistical interactions between NPGIs and environmental exposures (12,13). It is important to use accurate measures of genetic liability in such studies, however NPGIs are unsuitable due to their lack of biological clarity, their dependence on sample characteristics, and the risk that their associations are misinterpreted as evidence for genetic essentialism and determinism (14). A key argument against such interpretations is the presence of gene-environment correlation (rGE) (15) wherein NPGIs exhibit statistical associations with environmental factors which may in turn have an independent effect on mental health outcomes (16–18). Hence, NPGIs contain a combination of biological and environmental signal, which could upwardly bias their estimated “genetic” effects on mental health.

Despite this, the biological signal contained within NPGIs is likely to reflect biological processes which are genuinely pertinent to mental health. NPGIs are calculated based on genome-wide association studies (GWAS) of neuropsychiatric conditions including neurodevelopmental (e.g. attention deficit hyperactivity disorder; ADHD (19), and autism spectrum disorder; ASD (20)) and mental health (e.g. major depressive disorder; MDD (21), bipolar disorder; BD (22), post-traumatic stress disorder; PTSD (23), eating disorders; ED (24), anxiety disorders; AD (25), schizophrenia; SCZ (26), obsessive-compulsive disorder; OCD (27), and substance use disorders; SUD (8)) conditions. These GWAS have enhanced our understanding of the genetic contribution to poor mental health, with downstream analyses overwhelmingly implicating brain tissues as mediators of these genetic associations. A recent review (28) highlights the main mediating cell types for SCZ, BD and MDD, specifically glutamatergic neurons which connect cortical and subcortical regions, and GABAergic neurons which connect structures within each of these regions. However, other tissues have been implicated by downstream analyses of neuropsychiatric GWAS: genetic loci linked to the “cross-disorder” factor (resulting from a meta-analysis of eight conditions (29)) and depression (21) are significantly enriched for genes expressed in the pituitary gland and skeletal muscle, respectively. Hence, the biological signal remains heterogeneous and difficult to interpret.

One way to reduce this biological complexity could be to use PGIs based on GWAS of endophenotypes, instead of neuropsychiatric conditions. GWAS have been performed on a range of brain phenotypes including molecular and neuroimaging-based measures, many of which have demonstrated genomic overlap with neuropsychiatric conditions, making them candidate endophenotypes for poor mental health. It is relatively unknown whether endophenotype PGIs (EPGIs) are powerful enough to associate with mental health symptoms in population samples. Nominal associations have been observed between depression remission and PGIs for both caudate volume and protein levels (30,31), and PGIs of AI-derived neuroimaging phenotypes have nominally improved prediction of mental and behavioural disorders (32). An EPGI which robustly associates with mental health symptoms would be easier to interpret than an NPGI because the former could reasonably be conceived as a proxy for its respective endophenotype, e.g. cerebellar volume or global white matter integrity. Brain endophenotypes have demonstrated lower polygenicity than neuropsychiatric conditions (33,34) supporting the notion that they are less complex and more amenable to genetic characterisation. It remains unknown whether these properties could translate into EPGIs which exhibit less environmental signal than NPGIs.

As mentioned, NPGIs also contain environmental signal because they exhibit rGE: statistical associations with environmental exposures from across the life course (16,17,35), including those occurring during childhood and adolescence (18,36,37). Passive rGE refers to genetic associations with the parentally-influenced rearing environment, as opposed to active and evocative rGE wherein the individual’s own behaviour contributes to the observed gene-environment correlation. The rearing environment includes common experiences such as parental divorce and measures of socioeconomic disadvantage which, while not necessarily harmful, are linked to worse mental health in adulthood when examined at the population level (38–40). One possible reason could be that socioeconomic disadvantage (41), including parental unemployment (42) has been associated with adverse childhood experiences (ACEs), which in turn have been associated with higher odds of adverse mental health outcomes in adulthood (43). Furthermore, parental divorce is sometimes considered an ACE (44) and has been positively associated with offspring depression in adulthood (45). Parental marital status and measures of socioeconomic disadvantage during childhood have previously demonstrated statistical associations with offspring NPGIs in UK population samples (18). Furthermore, some of these associations differed across UK birth cohorts born a few decades apart (18). Hence, NPGIs are unsuitable measures of “genetic” effects not only because they contain environmental signal, but because such signal likely varies across sociocultural contexts.

Several approaches exist for disentangling the environmental signal in GWAS summary statistics, including genomic structural equation modelling, which allows the summary statistics for one GWAS to be statistically “partialled out” from another. Marees et al used this technique to remove the influence of socioeconomic position from the “heritability” (variance explained by genetic factors) of behavioural and neuropsychiatric traits, finding a substantial attenuation for ADHD and smoking behaviours (46). Another approach is within-family GWAS, whereby modelling kinship during GWAS has been shown to substantially attenuate the heritability of behavioural traits such as household income, while increasing it for biological traits such as asthma (47). Alternatively, path models can be used to model the direct and mediated effects of a PGI on an outcome via one or more environmental exposures, as demonstrated in youth samples for both educational achievement (48) and more recently for mental health (49). These studies estimated environmental mediation of 40% and up to 27.5%, respectively, for the effects of PGIs on these outcomes. These findings are all consistent with the notion that PGIs indexing behavioural and neuropsychiatric phenotypes contain environmental signal which could upwardly bias “genetic” associations with mental health outcomes.

In order to assess the performance of NPGIs and EPGIs, we use path models to estimate the direct and environmentally-mediated effects of eleven NPGIs and thirty EPGIs on adult mental health symptoms in four representative UK population samples. For PGIs significantly associated with mental health, effect size is estimated using random effects meta-analysis. Environmental mediation is estimated using three rearing environment exposures available in all four samples: whether the participant lived with both parents (family intact), whether the father was employed (father employed) and whether the mother completed secondary education (mother educated). To examine rGE in the data, we report each exposure’s associations with PGIs. An extensive set of sensitivity analyses is performed to examine the impacts of inclusion criteria, measurement (of PGIs, mother’s education, and mental health symptoms), covariate adjustment, model specification and age at symptom measurement. We hypothesise that NPGIs will exhibit greater environmental mediation and rGE than EPGIs, that environmental mediation and rGE will differ across samples, and that the representativeness of each sample will impact the number and direction of associations observed.

## Methods

### Samples

*Understanding Society* a.k.a. the UK Household Longitudinal Study (UKHLS) was established in 2009 and comprises around 100,000 participants from 40,000 UK households. The annual main survey collects data from adult participants (16+) on a range of topics. In waves 2 and 3 (2010–2012), blood samples were collected during a nurse visit, resulting in genetic data for around 9,900 participants. We perform separate analyses on two sub-samples due to their differing sample characteristics: the British Household Panel Survey (BHPS) which began in 1991 and was absorbed into UKHLS in 2009, and the General Population Sample (GPS) which began in 2009. BHPS had likely undergone substantial non-random attrition over approximately two decades prior to the nurse visit, while GPS was relatively newly-recruited.

The 1958 National Child Development Study (NCDS) contains over 17,000 individuals born in the same week in 1958. The biomedical survey was carried out at age 44, with over 6,300 participants providing genotype data. The 1970 British Cohort Study (BCS70) contains around 17,000 individuals born in the same week in 1970. The biomedical survey was carried out at age 46, with over 5,500 participants providing genotype data. There is a non-negligible amount of missing childhood data, such that a 2004 technical report found greater non-response in BCS70 compared to NCDS, especially when comparing the two cohorts’ first and third follow-up waves (50). The Millennium Cohort Study (MCS) contains almost 19,000 individuals born between 2000-2002. The study design includes over-sampling of Black and Minority Ethnic (BAME) participants (51). Saliva samples were collected for genotyping from over 7,500 participants when they were aged 14, along with over 12,000 parents.

At the time of data application, eleven, ten and six follow-up surveys had been released for NCDS, BCS70 and MCS, respectively. These were conducted between the ages of 7-62 for NCDS, 5-51 for BCS70, and 3-17 for MCS.

### Survey measures

In UKHLS, age was operationalised by scaling year of birth, which was exponentiated to create age^2^. For MCS, age was recorded at biospecimen collection. Sex was self-reported and verified by genotyping for UKHLS, and was recorded during the first survey for each of the three birth cohorts. For MCS, ethnicity was reported by the main respondent (usually mother) during the first survey.

“Family intact”, “father employed” and “mother educated” were binarized based on available data relating to childhood and adolescence. Family intact was based on variables indicating who was living with, or caring for, the respondent. Father employed was based on variables indicating the participants’ father’s labour market status, occupation, or financial contribution to the household. Mother educated was based on variables which indicate the participant’s mother’s completion of secondary education, or age of school completion. An alternative measure which included vocational qualifications was used in sensitivity analyses where possible. Full details can be found in the supplementary methods, online code and supplementary tables 1 – 4.

Continuous measures of psychological distress, taken from the earliest adult (16+) survey available, were used as outcomes, except for BCS70 which had substantial missingness at age 16. For UKHLS, wave 2 (2010–2012) was used. For NCDS, BCS70 and MCS, the age 23, 26 and 17 questionnaires were used, respectively. UKHLS used the 12-item General Health Questionnaire (GHQ-12) (52), NCDS and BCS70 used the 24-item Malaise inventory (53) (which captures psychological distress and somatic symptoms (54)) and MCS used the 6-item Kessler scale (55). In NCDS and BCS70, additional measures were examined in sensitivity analyses. Full details in the supplementary methods, online code and supplementary table 5.

### Genotype data QC

The samples differed in terms of genotyping platform, imputation method and quality control (QC) performed by the respective study teams prior to this study (see supplementary tables 6 and 7). To ensure harmonisation, we carried out further QC on all datasets using PLINK2 following the steps outlined in Coleman et al (56). Individuals of non-European ancestry were excluded prior to further QC in all samples except MCS. Twins and families with multiple mothers and fathers were excluded from MCS. Variants were removed if they were duplicates, or if an rsID could not be assigned.

Missingness checks were performed to ensure no variant or individual had >1% missing data. To avoid the impact of related trios on some of the following QC steps, MCS was then split into a children file set and a parents file set. Variants were removed based on Hardy-Weinberg equilibrium testing (p 0.00001). Linkage disequilibrium (LD)-pruned datasets were used to calculate relatedness and heterozygosity. Estimated third-degree relatives (or closer) were removed, as were individuals with high heterozygosity. Variants with minor allele frequencies (MAF) of <1% were then removed. To maximise similarity of PGI calculation, each dataset was filtered to retain only the remaining 3,673,101 variants which were present in all four datasets. The MCS children and MCS parents datasets were then restricted to participants of European ancestry.

Additional versions of the UKHLS and MCS datasets were generated for sensitivity analyses. UKHLS was MAF filtered to 0.5%. A version of MCS which included individuals of all ancestries was retained. These datasets will hereafter be referred to as UKHLS-rare and MCS-full, respectively.

Full details can be found in the supplementary methods, online code and supplementary table 8.

### PGI selection

Summary statistics for AD, ADHD, ASD, BD, cross-disorder, ED, MDD, OCD, PTSD, SCZ and SUD were downloaded from the Psychiatric Genomics Consortium (PGC) website https://pgc.unc.edu/for-researchers/download-results/ [accessed 7^th^ June 2024] and used to derive NPGIs.

Endophenotype selection was based on a convenient, semi-systematic approach. We focussed on endophenotypes measured in the human brain, with GWAS results uploaded to the NHGRI-EBI GWAS catalog (57). Full summary statistics had to be easily accessible, with signed coefficients to enable PGI calculation. Using genome-wide approaches such as LD score regression (LDSC), study authors must have demonstrated significant genomic overlap between the endophenotype and a mental health-related behavioural outcome, i.e. neuropsychiatric condition, relevant personality trait such as neuroticism, or relevant sub-clinical behaviour such as alcohol consumption. For convenience, the study title must have included the word “brain”. The study must not have focussed on a topic other than neuropsychiatric conditions or endophenotype measurement. Thirty endophenotypes (regional brain volumes, white matter microstructure phenotypes, cerebrovascular lesions, resting-state brain activity phenotypes, and longitudinal change in brain structure) from seven publications (58–64) were included (table 1).

For further details on PGI selection, see supplementary methods, supplementary table 9, and supplementary figure 1.

**Table 1.**
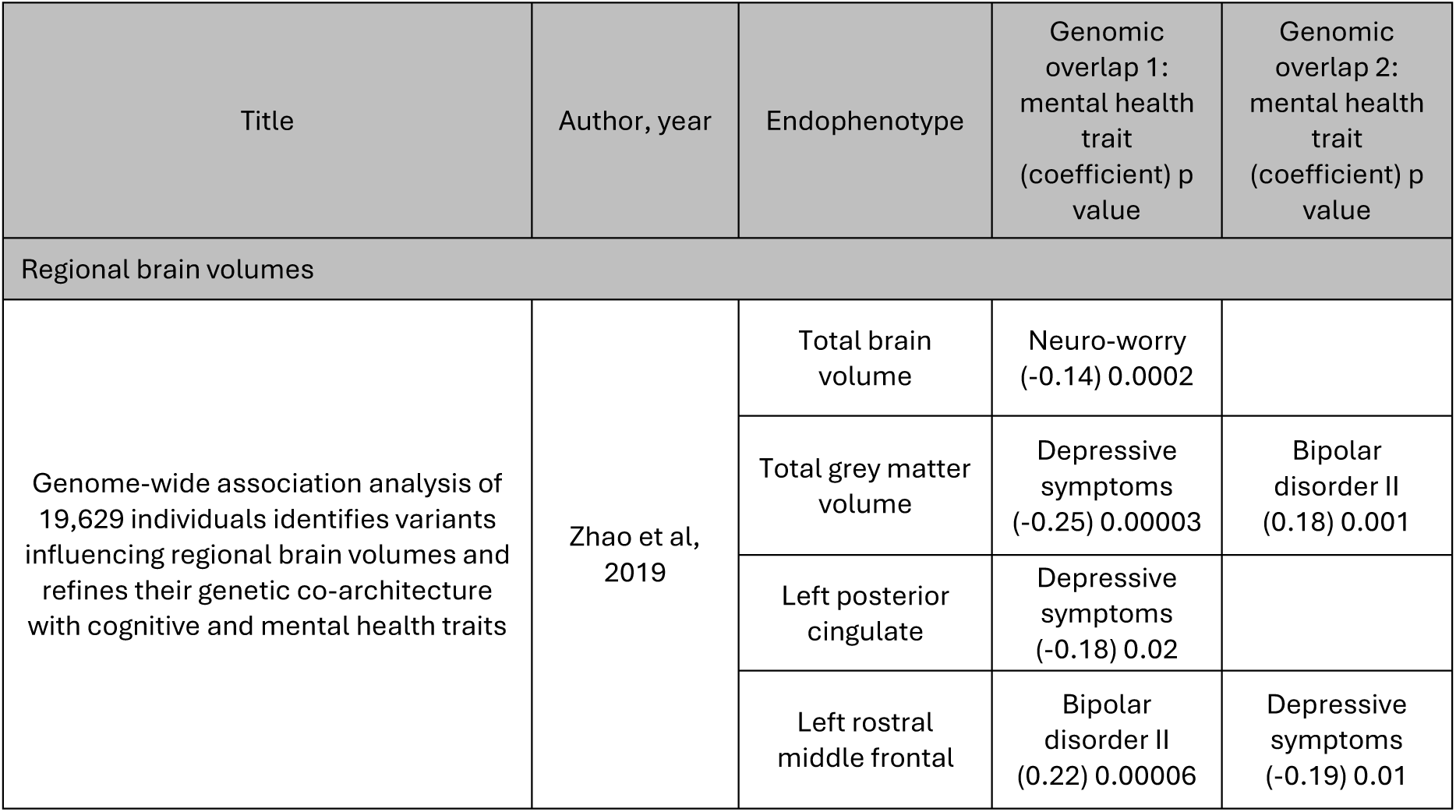
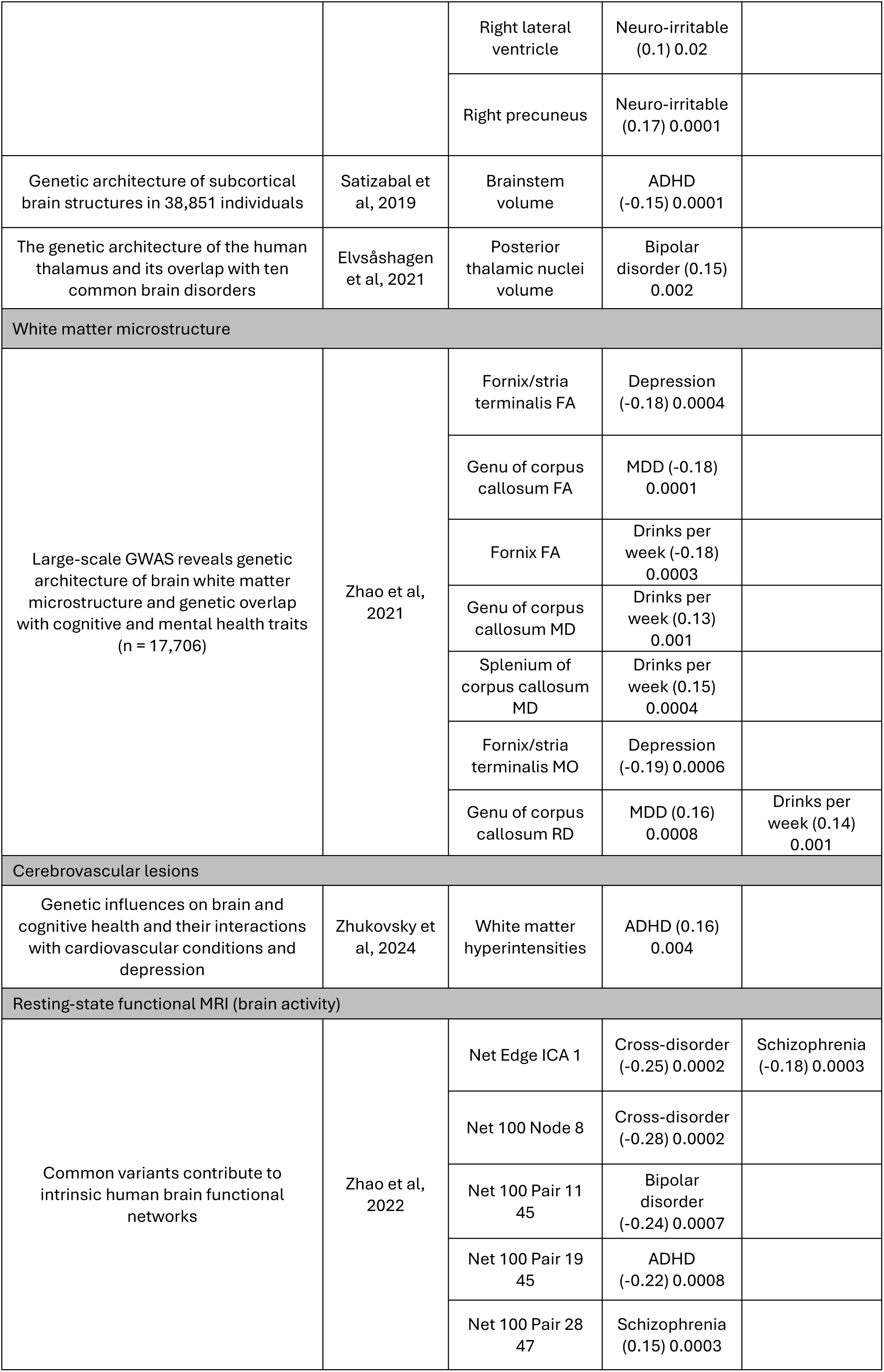
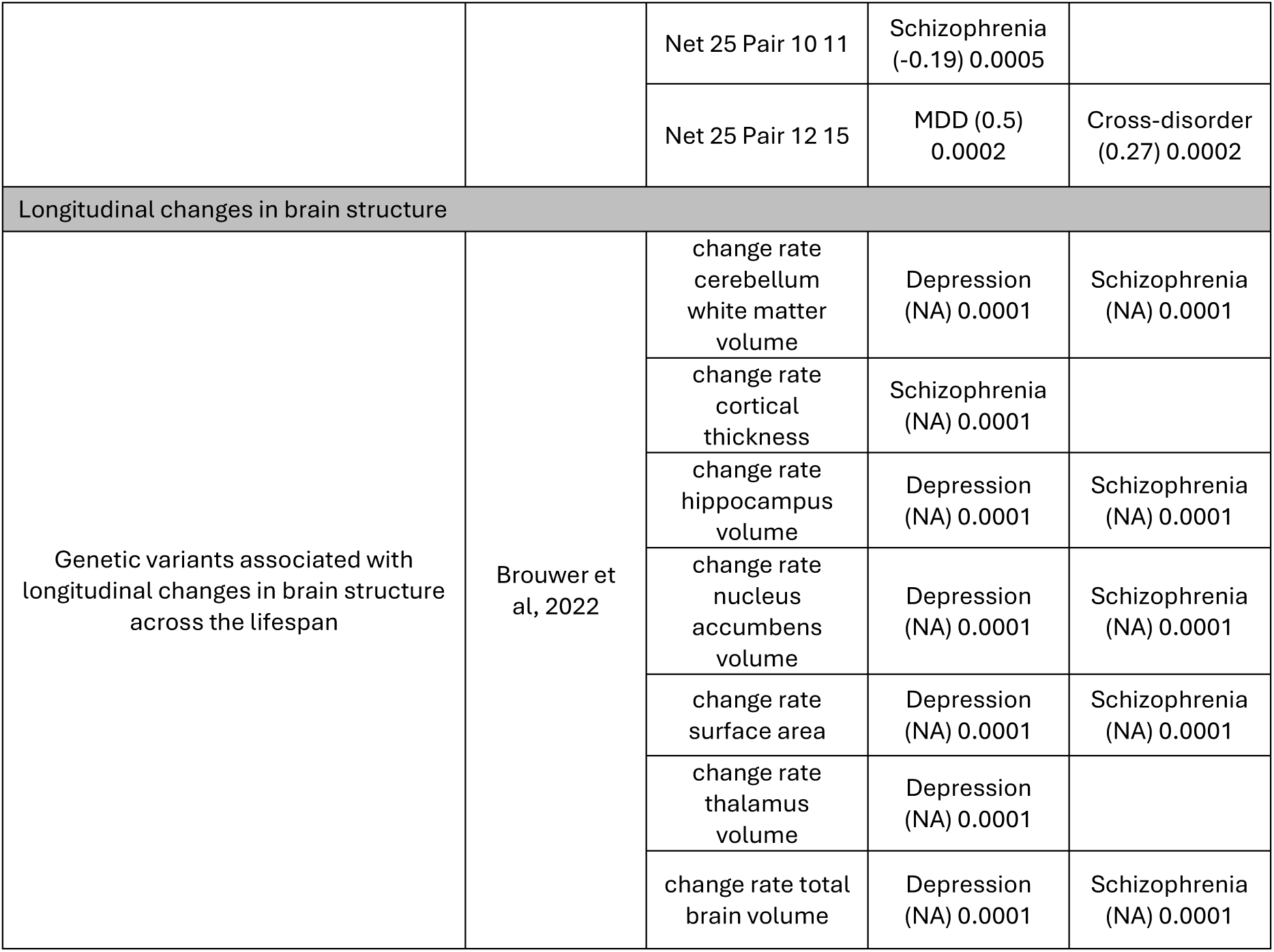
Genome-wide association studies (GWAS) and their respective endophenotypes whose summary statistics were used for deriving polygenic indices. N.B. Brouwer and colleagues did not report effect sizes in their genomic overlap analyses. ADHD = attention deficit hyperactivity disorder, FA = fractional anisotropy, ICA = independent components analysis, MD = mean diffusivity, MDD = major depressive disorder, MO = mode of anisotropy, MRI = magnetic resonance imaging, NA = not applicable, Neuro = neuroticism, RD = radial diffusivity.

| Title | Author, year | Endophenotype | Genomic overlap 1: mental health trait (coefficient) p value | Genomic overlap 2: mental health trait (coefficient) p value |
| --- | --- | --- | --- | --- |
| Regional brain volumes |  |  |  |  |
| Genome-wide association analysis of 19,629 individuals identifies variants influencing regional brain volumes and refines their genetic co-architecture with cognitive and mental health traits | Zhao et al, 2019 | Total brain volume | Neuro-worry (-0.14) 0.0002 |  |
|  |  | Total grey matter volume | Depressive symptoms (-0.25) 0.00003 | Bipolar disorder II (0.18) 0.001 |
|  |  | Left posterior cingulate | Depressive symptoms (-0.18) 0.02 |  |
|  |  | Left rostral middle frontal | Bipolar disorder II (0.22) 0.00006 | Depressive symptoms (-0.19) 0.01 |
|  |  | Right lateral ventricle | Neuro-irritable (0.1) 0.02 |  |
|  |  | Right precuneus | Neuro-irritable (0.17) 0.0001 |  |
| Genetic architecture of subcortical brain structures in 38,851 individuals | Satizabal et al, 2019 | Brainstem volume | ADHD (-0.15) 0.0001 |  |
| The genetic architecture of the human thalamus and its overlap with ten common brain disorders | Elvsåshagen et al, 2021 | Posterior thalamic nuclei volume | Bipolar disorder (0.15) 0.002 |  |
| White matter microstructure |  |  |  |  |
| Large-scale GWAS reveals genetic architecture of brain white matter microstructure and genetic overlap with cognitive and mental health traits (n = 17,706) | Zhao et al, 2021 | Fornix/stria terminalis FA | Depression (-0.18) 0.0004 |  |
|  |  | Genu of corpus callosum FA | MDD (-0.18) 0.0001 |  |
|  |  | Fornix FA | Drinks per week (-0.18) 0.0003 |  |
|  |  | Genu of corpus callosum MD | Drinks per week (0.13) 0.001 |  |
|  |  | Splenium of corpus callosum MD | Drinks per week (0.15) 0.0004 |  |
|  |  | Fornix/stria terminalis MO | Depression (-0.19) 0.0006 |  |
|  |  | Genu of corpus callosum RD | MDD (0.16) 0.0008 | Drinks per week (0.14) 0.001 |
| Cerebrovascular lesions |  |  |  |  |
| Genetic influences on brain and cognitive health and their interactions with cardiovascular conditions and depression | Zhukovsky et al, 2024 | White matter hyperintensities | ADHD (0.16) 0.004 |  |
| Resting-state functional MRI (brain activity) |  |  |  |  |
| Common variants contribute to intrinsic human brain functional networks | Zhao et al, 2022 | Net Edge ICA 1 | Cross-disorder (-0.25) 0.0002 | Schizophrenia (-0.18) 0.0003 |
|  |  | Net 100 Node 8 | Cross-disorder (-0.28) 0.0002 |  |
|  |  | Net 100 Pair 11 45 | Bipolar disorder (-0.24) 0.0007 |  |
|  |  | Net 100 Pair 19 45 | ADHD (-0.22) 0.0008 |  |
|  |  | Net 100 Pair 28 47 | Schizophrenia (0.15) 0.0003 |  |
|  |  | Net 25 Pair 10 11 | Schizophrenia<br>(-0.19) 0.0005 |  |
|  |  | Net 25 Pair 12 15 | MDD (0.5)<br>0.0002 | Cross-disorder<br>(0.27) 0.0002 |
| Longitudinal changes in brain structure |  |  |  |  |
| Genetic variants associated with longitudinal changes in brain structure across the lifespan | Brouwer et al, 2022 | change rate cerebellum white matter volume | Depression (NA) 0.0001 | Schizophrenia (NA) 0.0001 |
|  |  | change rate cortical thickness | Schizophrenia (NA) 0.0001 |  |
|  |  | change rate hippocampus volume | Depression (NA) 0.0001 | Schizophrenia (NA) 0.0001 |
|  |  | change rate nucleus accumbens volume | Depression (NA) 0.0001 | Schizophrenia (NA) 0.0001 |
|  |  | change rate surface area | Depression (NA) 0.0001 | Schizophrenia (NA) 0.0001 |
|  |  | change rate thalamus volume | Depression (NA) 0.0001 |  |
|  |  | change rate total brain volume | Depression (NA) 0.0001 | Schizophrenia (NA) 0.0001 |

### Generating PGIs

PGIs were generated using PRSice (v 2.3.5) (65). Six GWAS p-value thresholds (Pts) of 0.0001, 0.01, 0.05, 0.1, 0.5 and 1.0 were used as per the standard clumping and thresholding approach (66). PGIs were scaled and centered prior to analysis. Full details can be found in the supplementary methods. The MCS-full PGIs contained more variants than the European-only datasets, likely due to greater ancestral heterogeneity and thus different LD structure in the former. See supplementary methods and supplementary table 10 for details.

### Statistical analyses

All analyses were performed using R (v 4.4.0) in RStudio (v 2024.04.2 build 764). All were sex-stratified due to known sex differences in a) rates and pathophysiological mechanisms of mental health conditions (67,68), b) environmental risk factors (69–71), and c) neuroimaging measures of brain structure and function (68,72–74) including with respect to mental health (75). Complete case analysis was used. Due to partial collinearity between “family intact” and “father employed” resulting from participants whose father passed away, participants were excluded from the main analysis if either parent was deceased (for brevity, such participants are hereafter referred to as “bereaved”), but were included in a sensitivity analysis. UKHLS was split into two sub-samples prior to analysis: GPS and BHPS.

Pearson’s correlation was used to examine collinearity amongst PGIs and ancestry PCs.

Linear regression was used to estimate “total genetic effects” of each PGI (symptoms ∼ PGI), and the independent associations of all three exposures (symptoms ∼ family intact + father employed + mother educated). For GPS and BHPS, models were adjusted for age and age^2^. As PGIs and symptoms were scaled and centered, coefficients can be interpreted as betas. The *sem* function from the *lavaan* package (v 0.6-18) was used to run path models for each PGI. A direct path from PGI to mental health symptoms was used to estimate “direct genetic effects”. To model “indirect genetic effects”, paths from PGI to each of the three exposures were included, along with paths from each of the three exposures to mental health symptoms (see figure 1 for schematic diagram). The robust weighted least squares estimator (WLSMV) was used, and the exposures were treated as ordered variables. Coefficients were standardized using the *standardizedSolution* function, making them comparable with the regression betas. In GPS and BHPS, all paths from the PGI to another variable were adjusted for age and age^2^. Sensitivity analyses examined the impact of omitting this adjustment.

**Figure 1.**
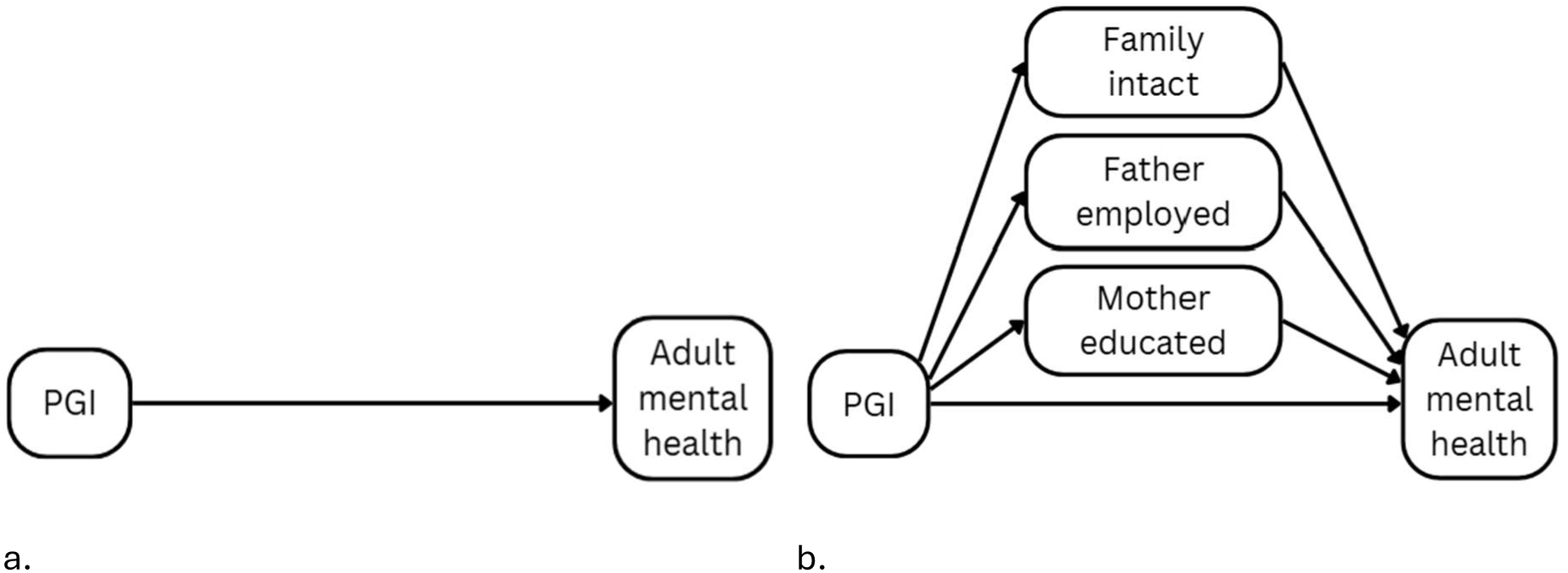
Schematic diagram of models. a. Linear regression estimating total genetic effects. b. Path models estimating direct and indirect genetic effects. PGI = polygenic index.

Coefficients and p values from the “direct genetic effects” paths were used to determine which PGIs were significantly associated with psychological distress. Environmental mediation was estimated using the formula *100 * ((total – direct) / total)* and will hereafter be referred to as “per cent mediated”. To minimise unstable estimates, this was only calculated where at least one of the associations (regression or path model) was at least nominally significant (p < 0.05). Positive estimates suggest environmental mediation while values of 0 or below are interpreted as total lack of mediation, i.e. direct genetic effects. To examine rGE, coefficients from paths between PGIs and exposures were also examined.

The main analyses are summarised in table 2.

**Table 2.**
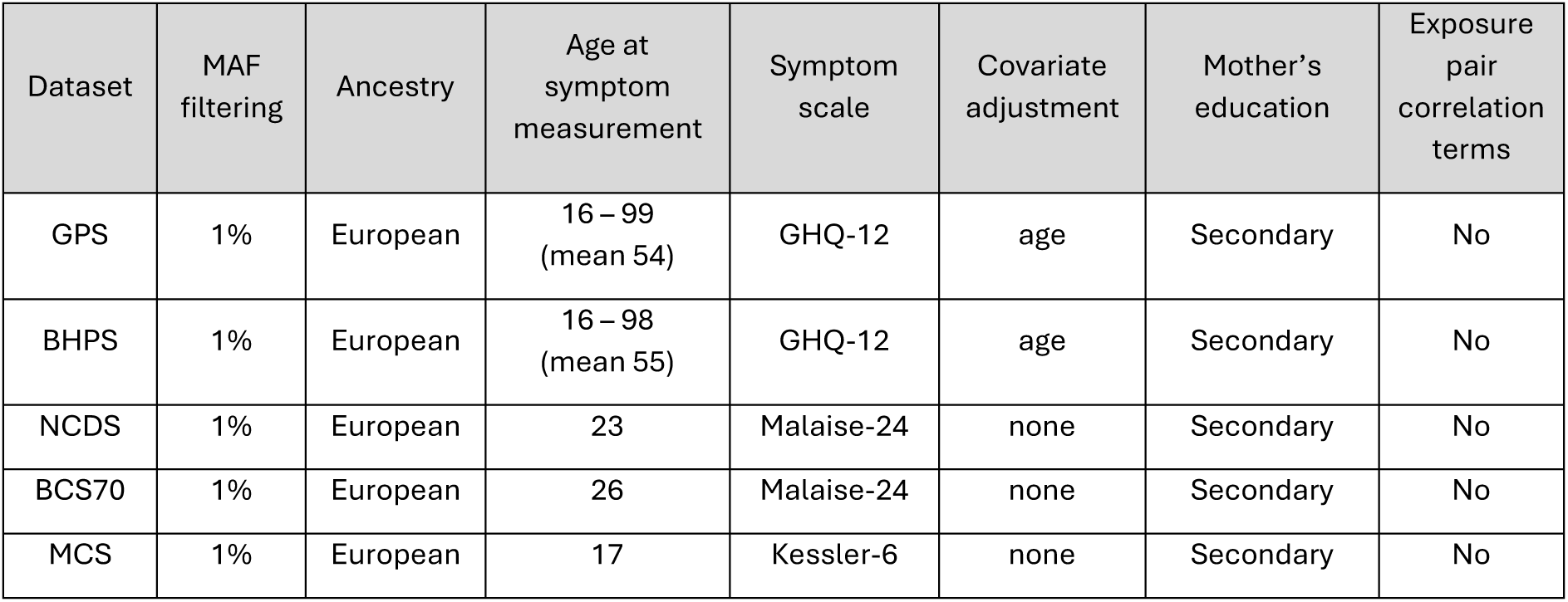
Details of main analyses. BCS70 = the 1970 British Cohort Study, BHPS = British Household Panel Survey, GHQ = general health questionnaire, MAF = minor allele frequency, MCS = Millennium Cohort Study, NCDS = the 1958 National Child Development Study.

All PGI analyses were run once per combination of PGI phenotype, P_t_, sex, and sample, i.e. 41 * 6 * 2 * 5, respectively. To correct for multiple testing, Bonferroni adjustment (76) was used with a numerator (alpha) of 0.05 and a denominator of 41 (number of phenotypes) i.e. 0.05/41 or 0.00122. This relatively lenient denominator was chosen due to substantial collinearity between the PGIs across P_t_s and phenotypes (see figure 2 and supplementary figure 2), and the relative stringency of the Bonferroni approach.

Meta-analyses were performed to obtain an overall estimate of PGIs’ associations with psychological distress. Sex-specific meta-analyses were performed on the direct path estimates using the *rma.mv* function from the *metafor* package (v 4.8-0) using a t distribution and restricted maximum likelihood estimation. As the datasets exhibit a range of age distributions and sociocultural contexts, “dataset” was set as a random effect to model the assumption that samples are drawn from heterogeneous populations.

### Sensitivity analyses

Sensitivity analyses were performed to examine the impacts of inclusion criteria, operationalisation (of PGIs, mother’s education, and symptom scale used), covariate adjustment, model specification and age at symptom measurement. For full details, see supplementary methods and supplementary table 11.

## Results

### Descriptive Statistics

Sample size ranged from n=682 to n=2,962. Rates of father employed and family intact were lowest, and mother educated highest, in MCS. See table 3. For samples used in sensitivity analyses, see supplementary tables 12-15.

**Table 3.**
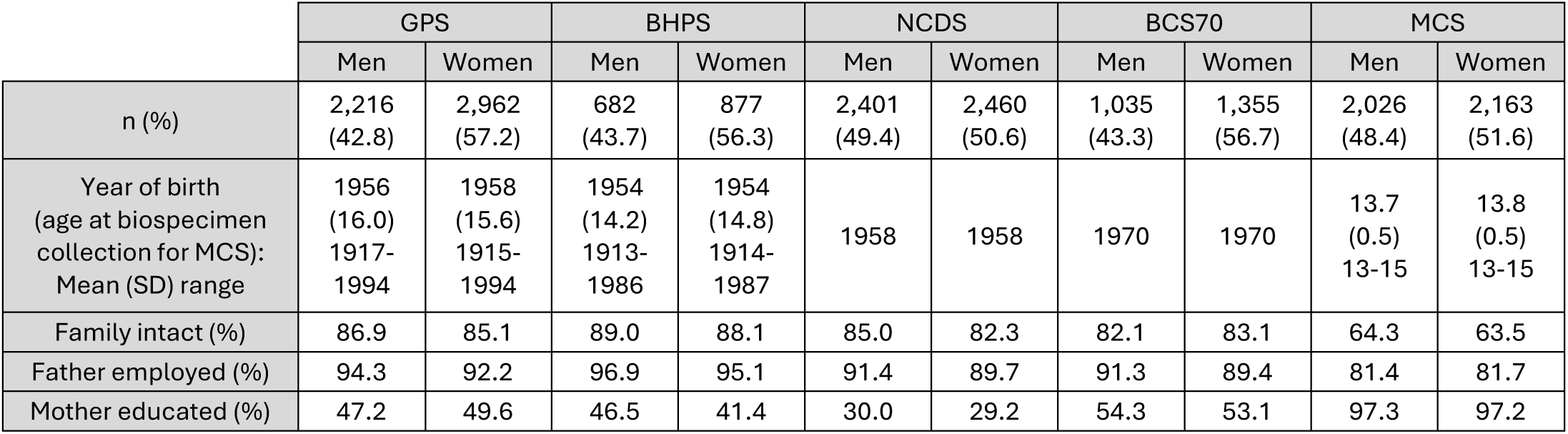
Descriptive statistics for the main analytical sample.

|  | GPS |  | BHPS |  | NCDS |  | BCS70 |  | MCS |  |
| --- | --- | --- | --- | --- | --- | --- | --- | --- | --- | --- |
|  | Men | Women | Men | Women | Men | Women | Men | Women | Men | Women |
| n (%) | 2,216<br>(42.8) | 2,962<br>(57.2) | 682<br>(43.7) | 877<br>(56.3) | 2,401<br>(49.4) | 2,460<br>(50.6) | 1,035<br>(43.3) | 1,355<br>(56.7) | 2,026<br>(48.4) | 2,163<br>(51.6) |
| Year of birth<br>(age at biospecimen<br>collection for MCS):<br>Mean (SD) range | 1956<br>(16.0)<br>1917-1994 | 1958<br>(15.6)<br>1915-1994 | 1954<br>(14.2)<br>1913-1986 | 1954<br>(14.8)<br>1914-1987 | 1958 | 1958 | 1970 | 1970 | 13.7<br>(0.5)<br>13-15 | 13.8<br>(0.5)<br>13-15 |
| Family intact (%) | 86.9 | 85.1 | 89.0 | 88.1 | 85.0 | 82.3 | 82.1 | 83.1 | 64.3 | 63.5 |
| Father employed (%) | 94.3 | 92.2 | 96.9 | 95.1 | 91.4 | 89.7 | 91.3 | 89.4 | 81.4 | 81.7 |
| Mother educated (%) | 47.2 | 49.6 | 46.5 | 41.4 | 30.0 | 29.2 | 54.3 | 53.1 | 97.3 | 97.2 |

### PGI correlations

In the combined UKHLS sample, we examined pairwise correlations between PGIs (P_t_ 1.0 only) and ancestry PCs. PC 1 is correlated with five NPGIs: SCZ (0.38), BD (0.22), cross-disorder (0.17), MDD (0.13) and ED (-0.13) and five EPGIs: total grey matter volume (GM) (-0.13) and four corpus callosum microstructure traits (-0.16 to 0.15). PC 3 is correlated with ASD (-0.11), brainstem volume (-0.10) and fornix/stria terminalis fractional anisotropy (FXST FA) (0.12). See figure 2 and supplementary figure 2 for correlation heatmaps.

**Figure 2.**
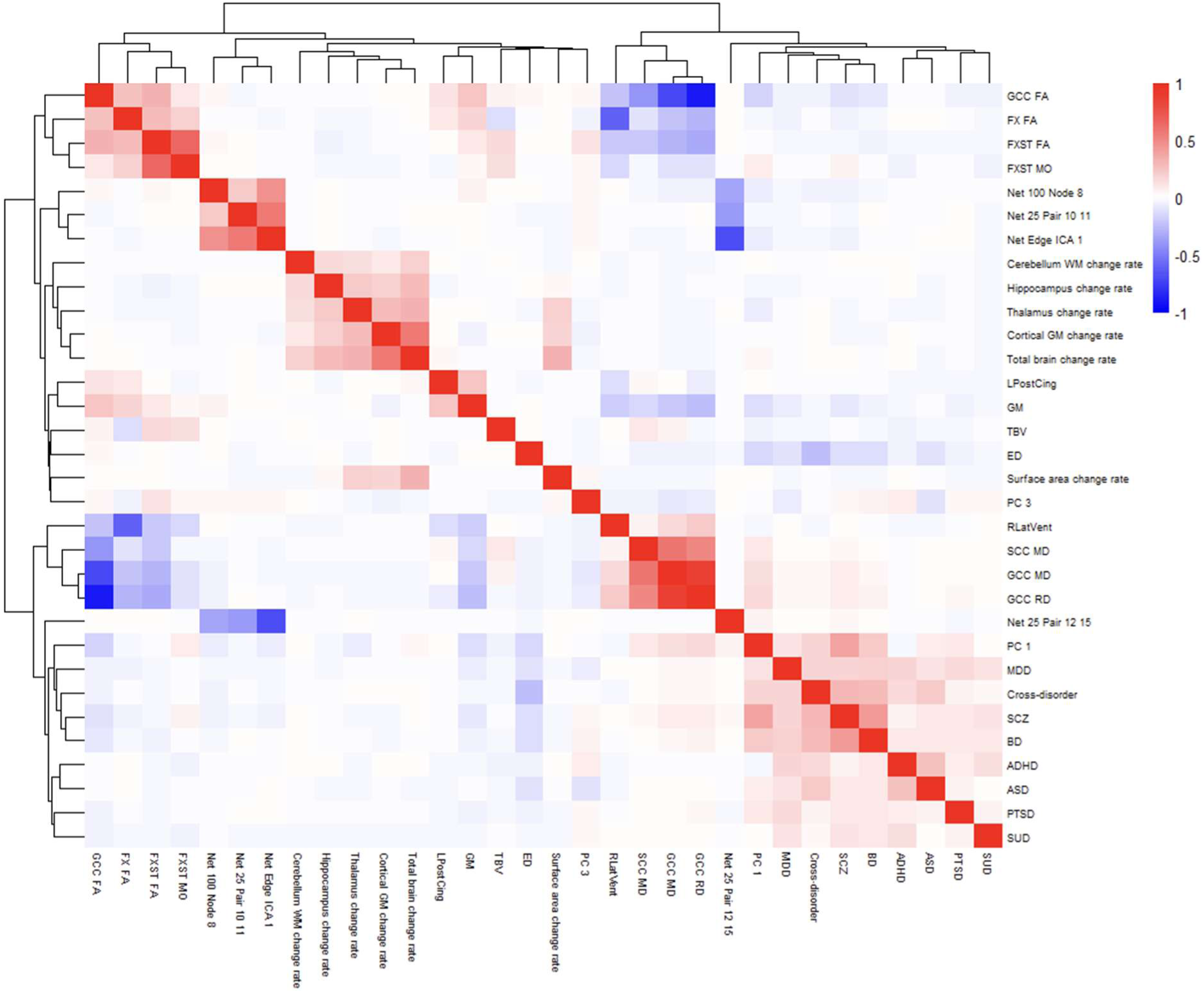
Correlation heatmap of PGIs and ancestry PCs in UKHLS (GPS and BHPS). For readability, plot is restricted to PGIs and PCs with at least four absolute correlations greater than 0.1, and to P_t_ 1.0. ADHD = attention deficit hyperactivity disorder, ASD = autism spectrum disorder, BD = bipolar disorder, ED = eating disorder, FA = fractional anisotropy, FXST = fornix/stria terminalis, GCC = genu of corpus callosum, GM = grey matter volume, ICA = independent components analysis, LPostCing = left posterior cingulate, MD = mean diffusivity, MDD = major depressive disorder, MO = mode of anisotropy, PC = principal component (of ancestry), PTSD = post-traumatic stress disorder, RLatVent = right lateral ventricle, SCC = splenium of corpus callosum, SCZ = schizophrenia, SUD = substance use disorders, TBV = total brain volume, WM = white matter.

### Exposure associations with psychological distress

Mutually-adjusted linear regressions show that each exposure was protective (p < 0.05) in multiple sex-stratified samples (figure 3). For path model results and sensitivity analysis results, see supplementary figure 3 and supplementary tables 16-22.

**Figure 3.**
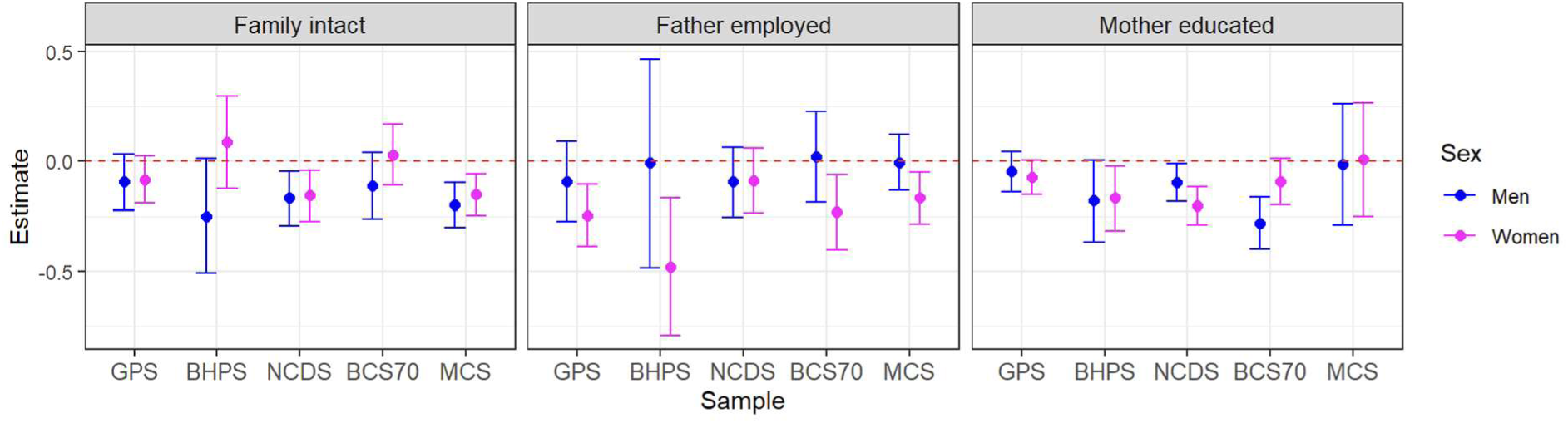
Exposure associations with psychological distress as estimated in mutually-adjusted linear regressions. Error bars represent 95% confidence intervals. Family intact was protective for both sexes in NCDS and MCS (p =< 0.008), father employed was protective in women across most datasets (p =< 0.008 excluding NCDS), and mother educated was protective in at least one sex for three samples: BHPS (women, p = 0.025), NCDS (both p =< 0.029) and BCS70 (men, p = 3.74e-6). BCS70 = the 1970 British Cohort Study, BHPS = British Household Panel Survey, GPS = general population sample of Understanding Society, MCS = Millennium Cohort Study, NCDS = the 1958 National Child Development Study.

### PGI associations with psychological distress

In the main analyses, eleven PGIs were significantly associated with psychological distress: eight NPGIs (AD, ADHD, ASD, cross-disorder, MDD, OCD, PTSD and SUD) and three EPGIs (left rostral middle frontal volume (LRostMidFront), GM and total brain volume (TBV)). See figure 4.

**Figure 4.**
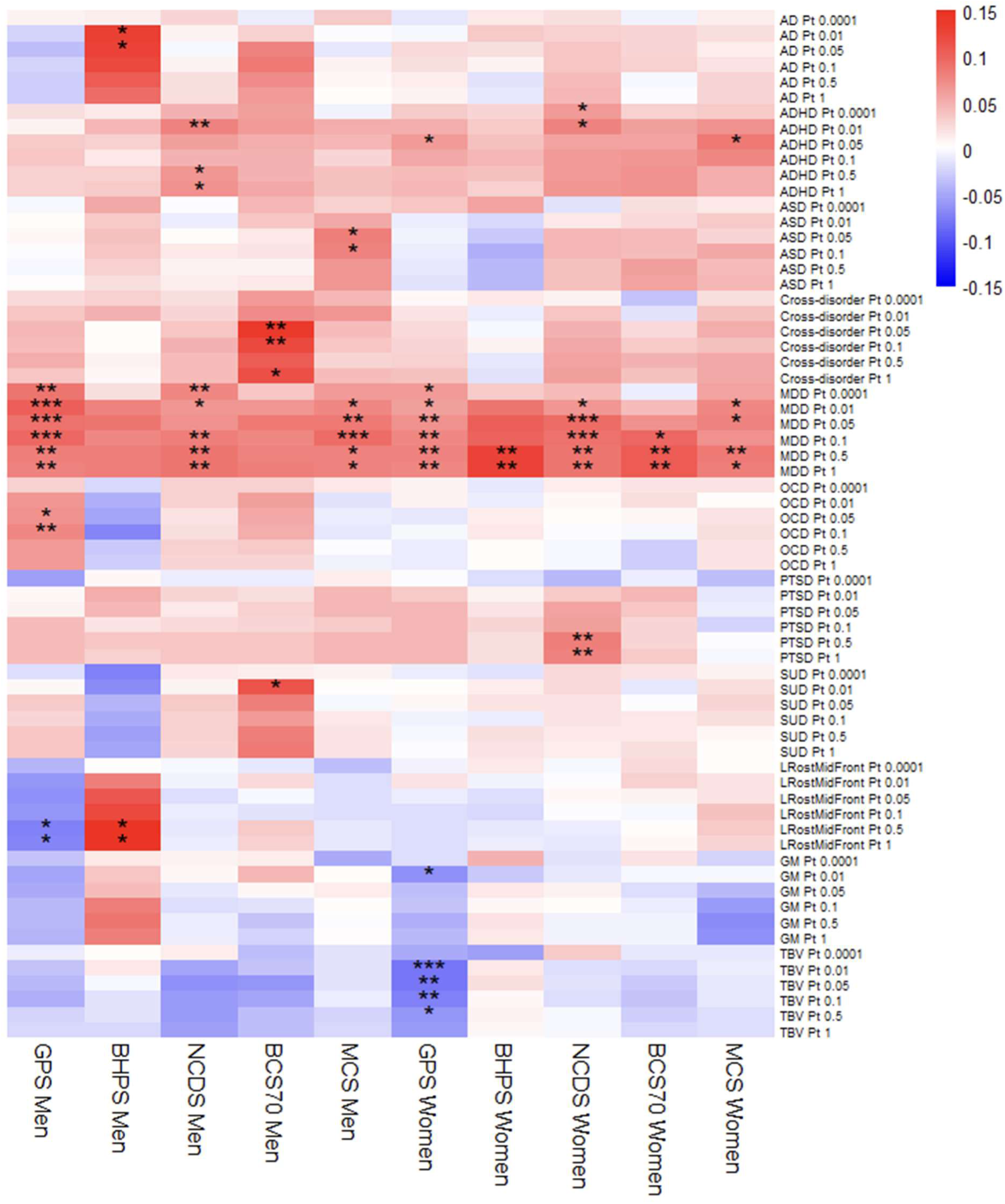
Associations between PGIs and mental health symptoms. * = p < 0.05/41, ** = p < 0.01/41, *** = p < 0.001/41. AD = anxiety disorders, ADHD = attention deficit hyperactivity disorder, ASD = autism spectrum disorder, BCS70 = the 1970 British Cohort Study, BHPS = British Household Panel Survey, GM = total grey matter volume, GPS = general population sample of Understanding Society, LRostMidFront = left rostral middle frontal region volume, MCS = Millennium Cohort Study, MDD = major depressive disorder, NCDS = the 1958 National Child Development Study, OCD = obsessive-compulsive disorder, Pt = p value threshold, PTSD = post-traumatic stress disorder, SUD = substance use disorders, TBV = total brain volume.

The MDD PGI was the most consistent, showing a positive association across eight out of ten sex-stratified samples, i.e. with the exceptions of men in BHPS and BCS70.

Meta-analyses were performed to estimate the magnitude of direct genetic effects on mental health. Only two P_t_s survived Bonferroni correction, both for the MDD NPGI in women: P_t_ 0.5 β=0.091 p=0.001 (see supplementary figure 4), and P_t_ 1.0 β=0.089 p=0.001. There was no evidence of heterogeneity (both p > 0.67). See supplementary results and supplementary table 23 for all meta-analysis results.

The total variance explained by all explanatory variables in psychological distress was small (see table 4). For path models, it ranged from 2.4 - 6.4% when using the most strongly associated MDD NPGI. In linear regressions, estimates were lower (1.6 - 4.3%) and, for MDD NPGI alone, lower still (0.8% to 2.0%). See figure 5.

**Table 4.** Maximum variance explained in psychological distress (%). BCS70 = the 1970 British Cohort Study, BHPS = British Household Panel Survey, GPS = general population sample of Understanding Society, MCS = Millennium Cohort Study, MDD = major depressive disorder, NCDS = the 1958 National Child Development Study, NPGI = neuropsychiatric polygenic index, P_t_ = p value threshold.

|  | GPS |  |  |  | BHPS |  |  |  | NCDS |  |  |  | BCS70 |  |  |  | MCS |  |  |  |
| --- | --- | --- | --- | --- | --- | --- | --- | --- | --- | --- | --- | --- | --- | --- | --- | --- | --- | --- | --- | --- |
| | $P_t$ | 1 | 2 | 3 | $P_t$ | 1 | 2 | 3 | $P_t$ | 1 | 2 | 3 | $P_t$ | 1 | 2 | 3 | $P_t$ | 1 | 2 | 3 |
| Men | 0.01 | 2.4 | 1.9 | 1.2 | 0.05 | 3.4 | 2.6 | 1.0 | 1.0 | 3.0 | 1.6 | 0.9 | 0.05 | 4.5 | 3.2 | 0.9 | 0.1 | 2.7 | 1.9 | 1.0 |
| Women | 0.5 | 3.8 | 2.3 | 0.8 | 1.0 | 6.4 | 4.3 | 2.0 | 0.05 | 4.4 | 2.6 | 1.1 | 1.0 | 3.2 | 2.1 | 1.3 | 0.5 | 4.0 | 2.2 | 0.9 |

<sup>1</sup> by the combination of MDD NPGI (optimal $P_t$ per sex-stratified sample), family intact, father employed and mother educated (and, for GPS and BHPS, age and age<sup>2</sup>) in *path models*.
<sup>2</sup> by the combination of MDD NPGI (optimal $P_t$ per sex-stratified sample), family intact, father employed and mother educated (and, for GPS and BHPS, age and age<sup>2</sup>) in *linear regression models*.
<sup>3</sup> by MDD NPGI (optimal $P_t$ per sex-stratified sample) *independent of* family intact, father employed and mother educated (and, for GPS and BHPS, age and age<sup>2</sup>) in *linear regression models*.

**Figure 5.**
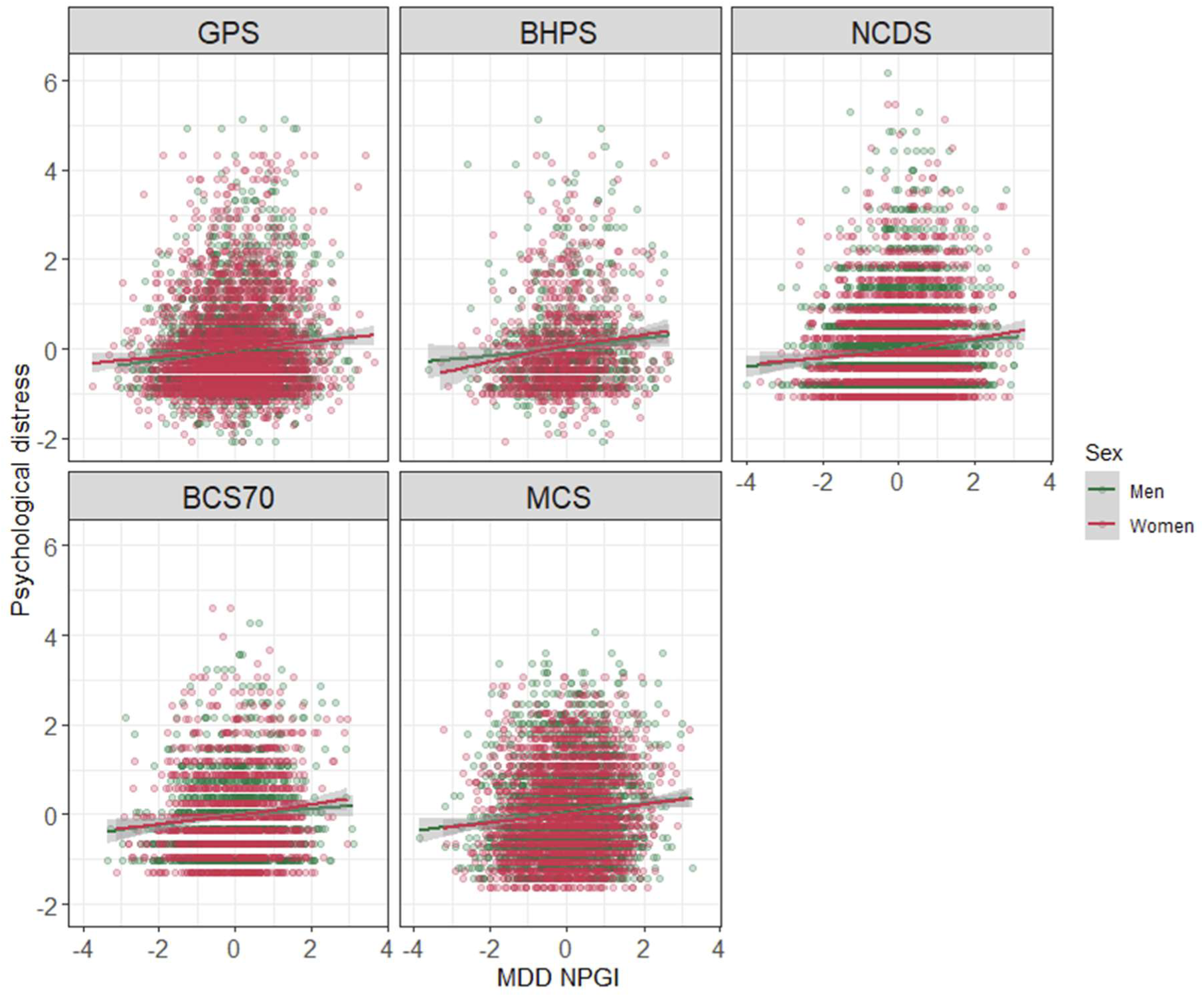
Bivariate correlation between psychological distress and MDD NPGI, using the optimal P_t_ per sex-stratified sample (optimal P_t_ = 0.01 for GPS men, 0.05 for BHPS men, NCDS women and BCS70 men, 0.1 for MCS men, 0.5 for GPS women and MCS women, and 1.0 for BHPS women, NCDS men and BCS70 women). BCS70 = the 1970 British Cohort Study, BHPS = British Household Panel Survey, GPS = general population sample of Understanding Society, MCS = Millennium Cohort Study, MDD = major depressive disorder, NCDS = the 1958 National Child Development Study, NPGI = neuropsychiatric polygenic index, P_t_ = p value threshold.

The PGI with the second-most consistent associations with psychological distress was ADHD, mostly showing positive associations in women (GPS, NCDS and MCS), but also in NCDS men. Other associations were specific to one sex-stratified sample. Of the NPGIs, five were associated in men (OCD in GPS, AD in BHPS, cross-disorder and SUD in BCS70, and ASD in MCS) and one in women (PTSD in NCDS). Of the EPGIs, one was associated in men (LRostMidFront in the GPS and BHPS sub-samples of UKHLS, with opposite directions of association across samples) and two in women (GM and TBV in GPS).

As EPGIs only exhibited associations in the age-representative sub-samples of UKHLS (as opposed to the other samples where symptoms were measured between the ages of 17 – 26), we ran *post hoc* exploratory linear regressions stratified by generation to examined whether associations were driven by the older participants. Amongst GPS women, the coefficient for GM EPGI is greatest in the youngest stratum (millennials aged 16-30 at the time of symptom measurement) though the association is non-significant. TBV PGI exhibits a suggestive negative association with psychological distress in GPS men aged 46-66 at the time of symptom measurement (p 0.01). Unlike in the four oldest strata, the coefficient for LRostMidFront in the youngest stratum of BHPS men is concordant with that of GPS men, however there are only n=15 and the association is non-significant. For full results, see supplementary table 24.

Heatmaps of sensitivity analysis results can be found in supplementary figures 5-10. Some of the main analyses’ less robust associations were replicated in at least one sex. Focussing on EPGIs, the LRostMidFront EPGI was associated with psychological distress in BCS70 men at age 16, GM EPGI in MCS-full women and TBV EPGI in NCDS men at age 50. When MCS was subset to participants whose parents both provided genetic data, all associations between PGIs and mental health were lost. Adjusting for parental PGIs made no further difference to the associations. Looking at different ages in NCDS and BCS70, the MDD NPGI tended to remain associated with malaise across the life course whereas the ADHD NPGI did not. A few other associations changed in the sensitivity analyses, but use of rare alleles, adjusting for ancestry PCs, and lack of adjustment for age made little difference.

### Environmental mediation of PGI associations with psychological distress

The extent of environmental mediation was estimated for PGIs with at least nominal associations with psychological distress in regression or path models. The two most robustly associated NPGIs, MDD and ADHD, consistently showed environmental mediation with estimates ranging from 1.6 - 24.5%, and 6.8 - 49.7%, respectively. All associations were mediated for SUD (7.4 – 51.6%), all were direct for OCD (-12.1 – -5.4%). Focussing on significant associations, those of ASD, GM and PTSD were consistently mediated, and those of cross-disorder and TBV were consistently direct. See figure 6 and supplementary table 25.

**Figure 6.**
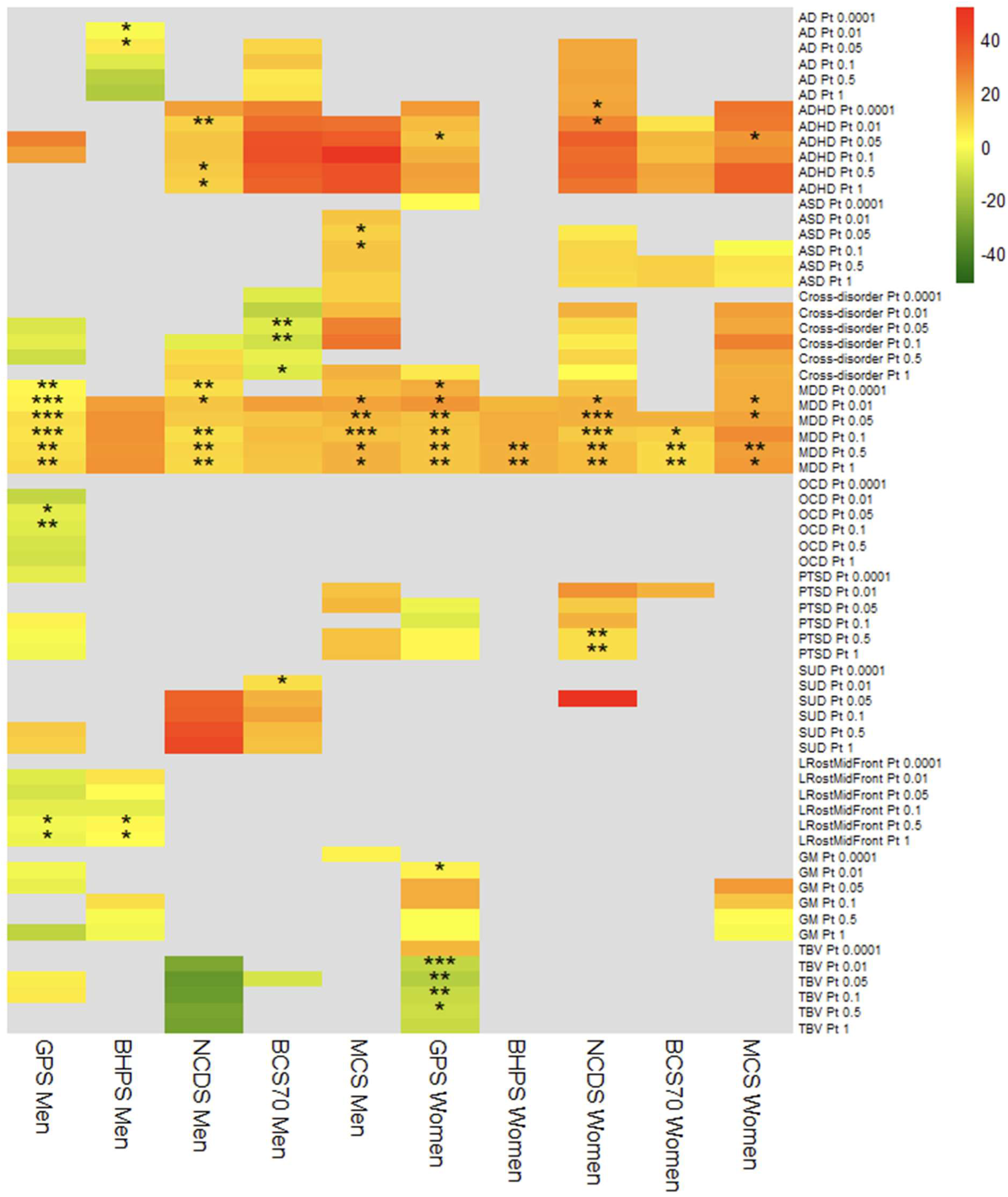
Estimates of “per cent mediated”, i.e. the extent to which polygenic index (PGI) associations with psychological distress are mediated by the rearing environment. Values of 0 and below represent no environmental mediation while positive values indicate some environmental mediation. Values were not calculated when both the linear regression (for estimating total genetic effects) and the path model (for estimating direct genetic effects) had p values greater than 0.05. * = p < 0.05/41, ** = p < 0.01/41, *** = p < 0.001/41. AD = anxiety disorders, ADHD = attention deficit hyperactivity disorder, ASD = autism spectrum disorder, BCS70 = the 1970 British Cohort Study, BHPS = British Household Panel Survey, GM = total grey matter volume, GPS = general population sample of Understanding Society, LRostMidFront = left rostral middle frontal region volume, MCS = Millennium Cohort Study, MDD = major depressive disorder, NCDS = the 1958 National Child Development Study, OCD = obsessive-compulsive disorder, Pt = p value threshold, PTSD = post-traumatic stress disorder, SUD = substance use disorders, TBV = total brain volume.

See supplementary figures 10-15 for heatmaps of per cent mediated estimates in sensitivity analyses. Mediation estimates were mostly attenuated by modelling pairwise correlation terms between the exposures, and with increasing age for NCDS and BCS70.

### Gene-environment correlation

For NPGIs, associations were generally negative such that increased genetic risk was associated with lower chance of family intact, father employed or mother educated. ADHD NPGI was associated with all three exposures, MDD NPGI with two (family intact and father employed) and SUD NPGI with two (family intact and mother educated). Sex differences were observed for several rGE associations, such that associations were significant for one sex but not the other in the same dataset.

### Family intact

Family intact was significantly associated with four NPGIs and one EPGI in at least one sex-stratified sample. The sample with the highest number of associations was MCS, while none were observed in BCS70. See figure 7a for main analysis results, and supplementary figures 17-22 for sensitivity analysis results.

**Figure 7.**
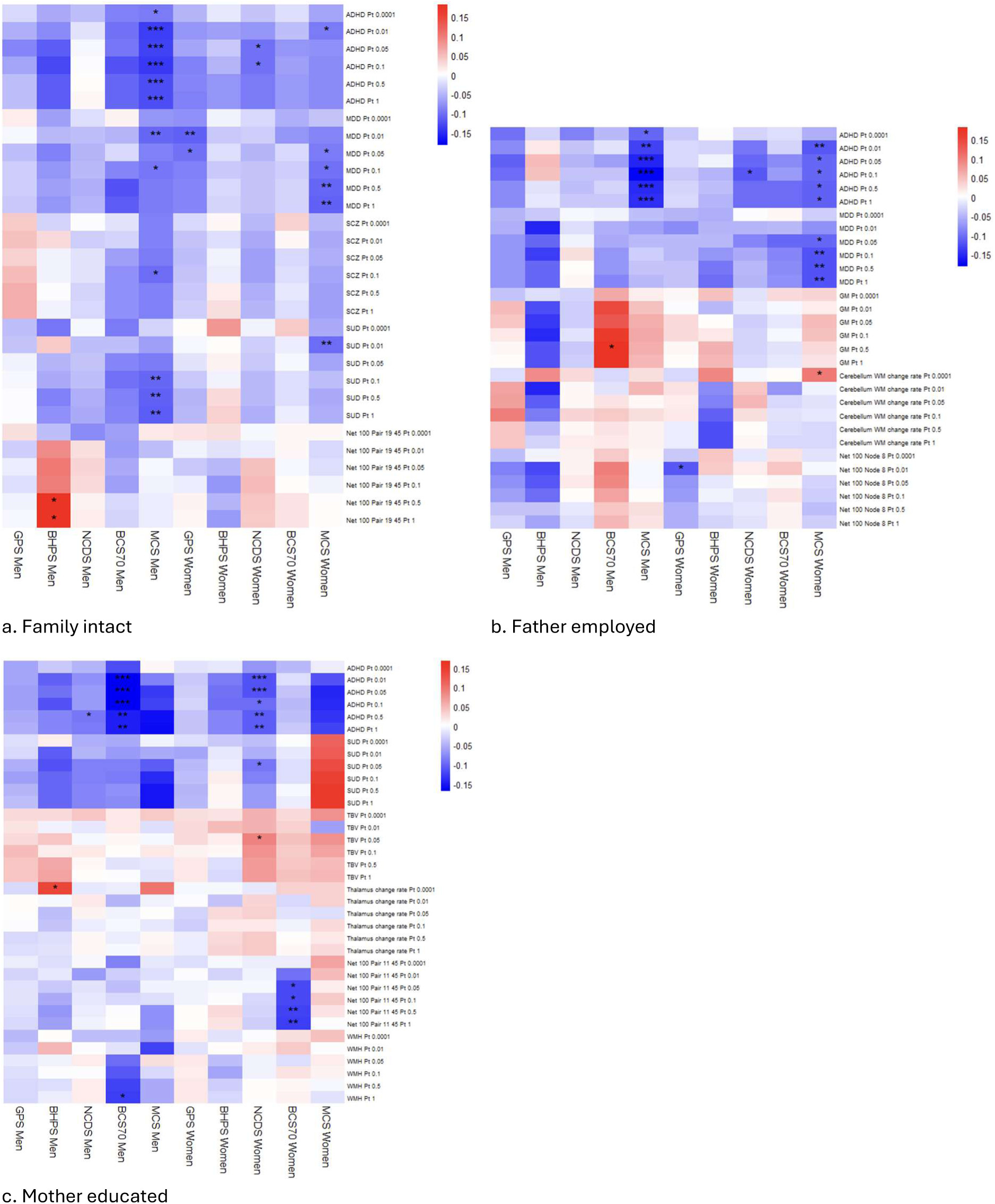
Associations between polygenic indices (PGIs) and exposures from the rearing environment. * = p < 0.05/41, ** = p < 0.01/41, *** = p < 0.001/41. ADHD = attention deficit hyperactivity disorder, BCS70 = the 1970 British Cohort Study, BHPS = British Household Panel Survey, GM = grey matter, GPS = general population sample of Understanding Society, MCS = Millennium Cohort Study, MDD = major depressive disorder, NCDS = the 1958 National Child Development Study, Pt = p value threshold, SCZ = schizophrenia, SUD = substance use disorders, TBV = total brain volume, WM = white matter, WMH = white matter hyperintensities.

### Father employed

Two NPGIs and three EPGIs were significantly associated with father employed in at least one sex-stratified sample. Again, the sample with the highest number of associations was MCS, while none were observed in BHPS. See figure 7b for main analysis results, and supplementary figures 23-28 for sensitivity analysis results.

### Mother educated

Two NPGIs and four EPGIs were significantly associated with mother educated. Associations were not observed in MCS or GPS, while the highest number of associations was observed in NCDS. See figure 7c for main analysis results, and supplementary figures 29-33 for sensitivity analysis results.

### Sensitivity analyses

Inclusion criteria made substantial differences to rGE. In MCS, the trios sub-sample exhibited almost no rGE, while further adjustment for parents’ PGIs made little difference. Including all ancestries substantially increased rGE in MCS. When bereaved participants were included, father employed became associated with SUD NPGI in NCDS men and MCS women.

Including vocational qualifications in mother’s education generally increased its rGE in BCS70. When age adjustment was omitted in GPS, family intact became associated with ADHD NPGI in women, consistent with the main analysis results for MCS and NCDS. Looking across time points in NCDS and BCS70, rGE was generally similar at multiple ages.

## Discussion

In this study, we examined whether endophenotype-based polygenic indices (EPGIs) could be used as measures of genetic risk for poor mental health, by comparing their performance to those trained on neuropsychiatric conditions (NPGIs). Compared to NPGIs, EPGIs are arguably more interpretable because they index biologically proximal phenotypes such as brain structure and function.

However, their ability to robustly associate with psychological distress, without exhibiting mediation via the rearing environment, was unknown. Using path models in four representative UK population studies, we demonstrate that neither NPGIs nor EPGIs exhibit associations with psychological distress that are both robust and un-mediated.

The MDD NPGI was the most robustly associated with psychological distress, showing associations in eight of ten sex-stratified UK sub-samples. Meta-analysis estimated that, for women in the UK, a one standard deviation increase of the MDD NPGI (P_t_ 0.5) is associated with an average increase in psychological distress of 9.1% of a standard deviation. However, its environmental mediation was estimated to range from 1.6 - 24.5% across sex-stratified samples. Furthermore, the MDD NPGI was negatively associated with family intact and father employed in a sample of young adults born between 2000 and 2002, in whom these exposures were shown to be both protective and, compared to older cohorts, less common. Hence, the only robustly-associated PGI is undermined as a “genetic” variable because it contains environmental signal.

We hypothesised that NPGIs would exhibit greater environmental mediation and rGE than EPGIs, but this was not clearly demonstrated. Focussing on the main analyses, all significant associations between PGIs and psychological distress were mediated for five NPGIs (MDD, ADHD, SUD, ASD, and PTSD) and one EPGI (GM), while all were direct for two NPGIs (OCD, cross-disorder) and one EPGI (TBV). Furthermore, PGIs for MDD, ASD, cross-disorder and GM (along with several other NPGIs and EPGIs) were modestly collinear with ancestry PCs, suggesting some shared signal with geographical location (77). Hence, environmental signal is widespread across most PGIs and is not confined to one category (neuropsychiatric vs endophenotype). The largest estimated magnitudes of mediation were, however, observed in NPGIs.

We also hypothesised that environmental mediation and rGE would differ across samples with differing sociocultural contexts. This was clearly demonstrated to be the rule rather than the exception. In the main analyses, PGIs generally showed more associations with exposures in samples where rates of the latter were lower, i.e. family intact and father employed in MCS, and mother educated in NCDS and BCS70. The impact of sample characteristics was also examined in two sensitivity analyses that were particular to MCS. These demonstrated that including participants of all ancestries, and excluding participants without parental DNA available, had a substantial impact on rGE. The latter drastically increased the exposure rates, and removed almost all of the PGIs’ associations with exposures and outcomes. Hence, genetic studies which rely on trio samples may be severely biased. It is unclear whether the additional rGE observed in the analyses on all ancestries was driven by differences in LD structure which impacted the operationalisation of PGIs. In another sensitivity analysis, the inclusion of participants who had lost a parent during youth led to associations between father employed and the SUD NPGI in two samples. Hence, rGE is highly dependent on who is included in the analytical sample.

Our final hypothesis was that the representativeness of each sample would impact the number and direction of associations observed. The MDD NPGI was associated with psychological distress in all but two of the ten sex-stratified samples: BHPS men and BCS70 men. Various forms of non-participation could underpin this lack of association. When blood samples were collected for genetic testing in BHPS, approximately two decades had elapsed since initial study recruitment.

Since this was an age-representative sample, loss-to-follow-up could have impacted the representativeness of BHPS to a greater extent than the birth cohorts, as older participants could have passed away during that time. BCS70 has a relatively high amount of missing data (50) which, if not missing at random, could have impacted representativeness. Furthermore, male sex has been associated with non-participation (78,79). Hence, these two sex-stratified samples are likely less representative than the eight others. Furthermore, a lack of representativeness could have contributed to the observed opposite sign of association between LRostMidFront EPGI and psychological distress in BHPS men compared to GPS men. Additionally, there were several unusual results in BHPS and BCS70 regardless of sex, and in men regardless of sample. For the former, ADHD NPGI was associated with women’s psychological distress in all samples except BHPS and BCS70, which in turn exhibited several unique rGE associations with EPGIs. Regarding sex differences, father employed was not associated with psychological distress in men, potentially due to collider bias as socioeconomic disadvantage is associated with non-participation (78,79).

Furthermore, despite there being no theoretical basis for observing sex differences in the rearing environment or in rGE, these were observed. This could arise due to sex-specific participation bias, which has previously been observed in genetic datasets (80). It may be possible to overcome the effects of limited representativeness by using weights, which have been shown to de-bias estimates of downstream analyses when used in GWAS (81), and which could also be applied to studies which use PGIs. This could be examined in future research.

To the best of our knowledge, this was the first study to examine whether a range of EPGIs were associated with psychological distress in four adult UK population samples. Looking across main analyses, sensitivity analyses and *post hoc* generation-stratified analyses, our results suggest that some EPGIs could serve as biologically interpretable indices of genetic risk of poor mental health, though better-powered GWAS may be required as no EPGI associations were significant in meta-analyses. The total brain volume (TBV) EPGI was protective against psychological distress in GPS women, and in middle-aged men in two samples (NCDS at age 50, and nominally in a middle-aged stratum of GPS). This pattern of association could suggest that higher genetically-predicted total brain volume is protective against poor mental health, but only in women and middle-aged men. This is a plausible finding considering that, phenotypically, total brain volume is smaller in women compared to men (82) and shows steeper decline over time in men compared to women (83).

Phenotypically, smaller total brain volume has been associated with worse mental health symptoms across the life course, i.e. in both preadolescents (84) and in a middle-aged and elderly sample (85). Hence, the TBV EPGI associations in this study are unlikely to be due to type I error. The fact that its associations with psychological distress were consistently direct (i.e. un-mediated) in the main analyses makes this a promising EPGI which warrants examination in future studies.

In the main analyses, the LRostMidFront and GM EPGIs were negatively associated with psychological distress in men and women in GPS, respectively. Both showed a replication in one other sample of the same sex in sensitivity analyses. While the direction of association for LRostMidFront EPGI was inverted in BHPS compared to GPS, this could be due to reduced representativeness of the former as discussed. Besides, the trend was concordant with GPS in the small number of millennials in BHPS, and the finding was replicated in BCS70 men at age 16.

Phenotypically, the gyrification of this region is lower in men compared to women (86,87) which could mean that its volume is more pertinent to psychological distress in men. The cortical thickness of this region has been linked to MDD incidence and duration (88), and its volume and cortical thickness have been linked to stress-related phenotypes in middle-aged men, specifically PTSD (89) and salivary cortisol (90), respectively. The association of GM EPGI in GPS women was replicated in MCS-full women. This sex-specific association is plausible given that total grey matter volume is, on average, lower in women compared to men (82). Total grey matter volume has been phenotypically associated with mental health symptoms in preadolescents (91) and with depression symptoms and status in clinical cohorts (92). Taken together, this suggests that LRostMidFront and GM EPGIs warrant further investigation as measures of genetic risk for poor mental health.

It is worth noting that the aetiology of neuropsychiatric conditions is far too complicated (93) to be satisfactorily explained by combining PGIs and measures of the rearing environment in a regression-based analysis. In the present study, explained variance in psychological distress was estimated at a maximum of just 6.4% for the combination of PGI, age and three measures of the rearing environment. We have demonstrated how, when examined at the population level, genetic factors, parental behaviours and psychological distress exhibit very weak correlations, which vary across sociocultural contexts, and which therefore do not determine one another.

### Limitations

There are several limitations in the present study. Firstly, we used fairly simple models. In the main analyses, we did not adjust for ancestry PCs, however we found that doing so in GPS made little difference. Furthermore, we focussed on passive rGE and so didn’t model evocative or active rGE. It is plausible that the participants’ own relational and occupational functioning – which likely has an impact on psychological distress – exhibits associations with PGIs such as ADHD, MDD and SUD, which were frequently associated with parents’ marital status, occupational status and education in the present study. Future research could model a greater number of covariates and mediators.

Secondly, the exposures were measured in a binary fashion, which may not be sufficiently nuanced. For example, “father employed” does not fully capture the level of socioeconomic advantage in the household, nor does it distinguish between unemployment and estrangement of the father.

Furthermore, measures were often based on a single point in time. However, a previous study has demonstrated that MDD NPGI exhibits significant rGE with a five-level measure of parental socioeconomic position in MCS, along with associations of p<0.05 with finance issues and father’s involvement in upbringing (18). In the present study we showed similar patterns of rGE in the same sample: MDD NPGI was associated with family intact and father employed. Hence, the coarse measures of the rearing environment used in the present study may be reasonable proxies for more granular and pertinent measures.

Thirdly, our outcomes were limited to cross-sectional, continuous measures of psychological distress. Further research could focus on other outcomes such as the chronicity of symptoms, or health service use.

Fourthly, while we consider EPGIs to be more interpretable than NPGIs, it is important to acknowledge their limitations. The endophenotypes measured in their respective GWAS were likely influenced by a range of factors besides genetics, including environmental exposures and stochastic variation. Direction of causality between such endophenotypes and mental health outcomes can only be established with longitudinal neuroimaging studies.

Finally, our main analyses focussed on adult participants of European ancestry. MCS was the only sample with participants of non-European ancestry, and including them had a substantial impact on rGE. However, we did not ascertain the reasons for this, which could include increased sample size and the greater number of genetic variants included in the PGIs in this sample. Furthermore, this birth cohort is still relatively young, so we could not examine the impact of including older participants of different ancestries. Finally, these analyses were specific to the UK context, so inferences cannot be made beyond the UK. Future research should examine a wide range of populations from across the world, including all ages and ancestries.

## Conclusion

In conclusion, this study demonstrates that neuropsychiatric polygenic indices contain environmental signal. The two key lines of evidence supporting this are the fact that direct genetic effects were usually smaller than total genetic effects, and the observed correlations between NPGIs and the rearing environment, which in turn vary according to sample characteristics. Hence, NPGI associations with mental health outcomes cannot be interpreted as purely “genetic”. In seeking to identify an endophenotype-based alternative to NPGIs, we demonstrate that some brain volume-based PGIs show associations with psychological distress in a minority of sex-stratified sub-samples with different age distributions. These inconsistent associations could result from true between-group heterogeneity in the biological underpinnings of psychological distress, or may be due to under-powered neuroimaging GWAS. Future work should focus on EPGIs based on larger GWAS, and should include a broader range of target populations, with greater variation in sociocultural context, age and ancestry.

## Acknowledgements

A.D. was supported by the ESRC (ES/T00200X/1, project reference number 2765580). L.S. is supported by MR/W004984/1. M.K. is supported by the University of Essex, ESRC (RES-596-28-0001) and ESRC (ES/S012486/1; ES/Y003071/1). J.M. is supported by ES/Y003071/1. The funders had no involvement in the analysis or preparation of the paper.

We would like to thank the study participants who gave their time and biological samples to support population research. We acknowledge the use of the High Performance Computing Facility (Ceres) and its associated support services at the University of Essex in this work. We are grateful for the technical advice from Tim Morris, Liam Wright, Alexander Hatoum and Grace Lutter.

## Author contributions

A.D. designed the project, conducted the analyses and drafted the manuscript. L.S. was the principal supervisor and M.K. was the secondary supervisor. All authors provided feedback on the manuscript.

## Data availability

Information on how to access the data can be found on the *Understanding Society* website https://www.understandingsociety.ac.uk/documentation/health-assessment/accessing-data/ and on the Centre for Longitudinal Studies website https://cls.ucl.ac.uk/data-access-training/genetic-data-and-biological-samples/.

## Code availability

The code is available at https://github.com/AnnaDearman/endophenotype-polygenic-indices/.

