## Supplementary materials for "Evaluating the performance of polygenic indices of neuropsychiatric conditions and brain endophenotypes in four UK population samples"

### **Supplementary methods**

#### ***Survey measures***

For full information, code can be found at https://github.com/AnnaDearman/endophenotype-polygenic-indices/blob/main/survey_data_processing.R

*Family intact*

"Family disruption" was derived first to obtain more granular information regarding family circumstances (values: "intact", "parent died", "parent moved", "other"). This was then used to assign "family intact" (1="intact", 0="parent died/parent moved/other"). Any evidence of family disruption was considered to warrant a value of 0, regardless of whether parents lived together subsequently. "Intact" was assigned towards the end of the logical tests for all datasets, to ensure other scenarios and conflicting answers were ruled out.

| | UKHLS | | NCDS | | BCS70 | | MCS | |
|---|---|---|---|---|---|---|---|---|
| Family disruption | n | % | n | % | n | % | n | % |
| Family intact | 6,868 | 79.52 | 4,970 | 79.22 | 2,629 | 73.99 | 4,341 | 59.11 |
| Parent moved | 913 | 10.57 | 394 | 6.28 | 553 | 15.56 | 2,863 | 38.98 |
| Parent died | 537 | 6.22 | 296 | 4.72 | 184 | 5.18 | 140 | 1.91 |
| Other | 319 | 3.69 | 614 | 9.79 | 187 | 5.26 | 0 | 0 |
| Missing | 640 | | 50 | | 1,654 | | 235 | |

Supplementary table 1. Family disruption rates, by reason. Values do not represent those in the final analytical samples. BCS70 = the 1970 British Cohort Study, MCS = Millennium Cohort Study, NCDS = the 1958 National Child Development Study, UKHLS = the UK Household Longitudinal Study (Understanding Society).

| | UKHLS | | NCDS | | BCS70 | | MCS | |
|---|---|---|---|---|---|---|---|---|
| Family intact | n | % | n | % | n | % | n | % |
| 1 | 6,868 | 79.52 | 4,970 | 79.22 | 2,629 | 73.99 | 4,341 | 59.11 |
| 0 | 1769 | 20.48 | 1,304 | 20.78 | 924 | 26.01 | 3,003 | 40.89 |

Supplementary table 2. Family intact rates. Values do not represent those in the final analytical samples. BCS70 = the 1970 British Cohort Study, MCS = Millennium Cohort Study, NCDS = the 1958 National Child Development Study, UKHLS = the UK Household Longitudinal Study (Understanding Society).

For Understanding Society (UKHLS), questions were asked of adult participants retrospectively. The main question was asked in relation to age 16. For the 1958 National Child Development Study (NCDS), due to the survey questions used, values of "intact" are based on lack of contrary information, so for participants with the least available information there is a higher risk of assigning incorrect values. Hence, a count of the number of productive interviews was derived first. After assigning values other than "intact", n=50 participants with 0-1 productive interviews were excluded, then "intact" was assigned to n=4,970, followed by a few values of "other". For the 1970 British Cohort Study (BCS70), there was a lot of missingness (n=1,654, 31.8%). The Millennium Cohort Study (MCS) does not have an age 16 wave, so information from up to age 14 was used.

*Father employed*

In UKHLS, the question regarding father's employment was asked in relation to age 14. While MCS has an age 14 questionnaire, NCDS and BCS70 do not, so for these cohorts the information was derived based on other time points, primarily age 15/16 but due to substantial missing data, age 11 was also used. For NCDS, the wording of questions was problematic for harmonisation as they tended to focus on "father figure" rather than the biological father. However, an attempt was made to identify whether the father figure was the biological father using variable **n104** ("Relationship person acting as father"). 99.57% of those assigned 1 had values of "natural father" for this variable, while the remaining n=20 had father's employment noted as a source of income at age 16.

For MCS, most values were based on age 14 variables regarding residency and employment situations, along with variables reflecting who answered the main and partner surveys. For many values, the mother's variable "does absent parent contribute to maintenance?" was relied upon – financial contribution does not necessarily indicate employment, but this assumption seems preferable to losing so many participants by assigning missing values (n=1,126).

| | UKHLS | | NCDS | | BCS70 | | MCS | |
|---|---|---|---|---|---|---|---|---|
| Father employed | n | % | n | % | n | % | n | % |
| 1 | 8,037 | 89.38 | 5,172 | 86.29 | 4,151 | 87.02 | 5,610 | 79.28 |
| 0 | 955 | 10.62 | 822 | 13.71 | 619 | 12.98 | 1,466 | 20.72 |
| Missing | 285 | | 350 | | 437 | | 503 | |

Supplementary table 3. Father employed rates. Values do not represent those in the final analytical samples. BCS70 = the 1970 British Cohort Study, MCS = Millennium Cohort Study, NCDS = the 1958 National Child Development Study, UKHLS = the UK Household Longitudinal Study (Understanding Society).

*Mother educated*

Given the drastic shifts in women's education across the UK since the oldest participants' parents attended school, and the variables available, "mother educated" was dichotomised according to completion of secondary school. None of the questions were asked in relation to a specific age of the mother or participant.

For UKHLS and NCDS, there was a lack of detailed information. For UKHLS, one variable was used, with a high rate of missingness (n=1,148). For NCDS, 1 was assigned where mothers stayed in education after the minimum leaving age, while not staying earned a value of 0. The "age mother left full-time education" variable was used to assign a small minority of values, however its levels were ambiguous, so participants with values of "14 to 15 years" and "15 to 16 years" and no other information were excluded, as the minimum school leaving age was likely 15 for the vast majority of mothers. A sensitivity analysis was performed using an alternative operationalisation which takes into account vocational qualifications, for datasets with sufficiently rich data (BCS70 and MCS). For BCS70, the vast majority of values were assigned based on questions asked when the child was age 10/11 regarding mother's qualifications. Most remaining values were assigned based on school leaving age, followed by educational and qualification questions asked when the child was 5.

For MCS, missingness was high (n=1,390). Variables originated from the mother's questionnaire. Questions regarding academic qualifications accounted for the majority of 1s (84.3%) while the remainder, and all of the 0s, were assigned using the "age left full time education" variable – those who were 16 or older were assigned 1 and those who were 15 or younger were assigned 0. For "mother qualifications", an additional n=110 were assigned 1s based on vocational qualifications, plus n=6 who at some point acquired new qualifications between interviews.

| | UKHLS | | NCDS | | BCS70 | | MCS | |
|---|---|---|---|---|---|---|---|---|
| Mother educated | n | % | n | % | n | % | n | % |
| 1 | 3,849 | 47.35 | 1,799 | 29.30 | 2,442 | 49.28 | 5,938 | 96.24 |
| 0 | 4,280 | 52.65 | 4,340 | 70.70 | 2,513 | 50.72 | 232 | 3.76 |
| Missing | 1,148 | | 185 | | 252 | | 1,390 | |

Supplementary table 4. Mother educated rates. Values do not represent those in the final analytical samples. BCS70 = the 1970 British Cohort Study, MCS = Millennium Cohort Study, NCDS = the 1958 National Child Development Study, UKHLS = the UK Household Longitudinal Study (Understanding Society).

*Deriving additional outcomes*

The variables in supplementary table 5 were used in sensitivity analyses. All were scaled and centered separately by sex and sample before excluding individuals with missing environment data.

| Study | Age | Scale | Number of items | Derivation |
|---|---|---|---|---|
| NCDS | 33 | malaise | 24 | The total number of values of "1" across 24 variables beginning with "n5042" were added |
| NCDS | 42 | GHQ psychological distress | 12 | Coded missing values were set to missing, then GHQ items 1, 3, 4, 7, 8 and 12 were reverse-coded, then all 12 items were added |
| NCDS | 42 | malaise | 24 | Coded missing values were set to missing, then the total number of values of "1" across 24 variables beginning with "mal" (malaise) were added |
| NCDS | 50 | malaise | 9 | **ND8MAL** was used |
| BCS70 | 16 | GHQ psychological distress | 12 | Coded missing values were set to missing, then GHQ items "c5i[1-6]" were reverse-coded, then all 12 items were added |
| BCS70 | 16 | malaise | 22 | **BD4MAL** was used |
| BCS70 | 30 | GHQ psychological distress | 12 | Coded missing values were set to missing, then GHQ items 1, 3, 4, 7, 8 and 12 were reverse-coded, then all 12 items were added |
| BCS70 | 30 | malaise | 24 | **BD6MAL** was used |
| BCS70 | 34 | Kessler psychological distress | 4 | Coded missing values were set to missing, then the total across variables "b7k[1-4]" was reverse-coded |
| BCS70 | 34 | malaise | 9 | **BD7MAL** was used |
| BCS70 | 42 | malaise | 9 | **BD9MAL** was used |
| BCS70 | 46 | malaise | 9 | **BD10MAL** was used |

Supplementary table 5. Additional mental health outcomes used for sensitivity analyses. BCS70 = the 1970 British Cohort Study, GHQ = general health questionnaire, NCDS = the 1958 National Child Development Study.

***Genetic data QC***

The UKHLS genetic data comprised 24.7 million genetic variants (including rare variants) and were supplied as 22 file sets (one per autosome), while NCDS, BCS70 and MCS each comprised between 7.5 – 8.7 million variants with minor allele frequency (MAF) >1% and were supplied as one file set each. Principal components (PCs) of ancestry and details regarding genotyped sex were supplied for UKHLS only. All study teams provided ancestry information in a categorical form, i.e. genetically-estimated continental superpopulation. Around 84% of the MCS genetic dataset is of European ancestry, while for the other datasets values are higher (93 - 100%). The main analyses were conducted in European-only (sub)samples for all four datasets, with sensitivity analyses conducted in the full, ancestrally-diverse, MCS sample.

For details of the genotyping, imputation and quality control (QC) performed by the respective study teams prior to this study, see supplementary tables 6 and 7 which summarise the information available in the readme files supplied by the University of Essex, in Prins et al (1), and in the online Centre for Longitudinal Studies (CLS) documentation (web pages accessed 6th December 2024): https://cls-genetics.github.io/docs/NCDS.html; https://cls-genetics.github.io/docs/BCS70.html; https://cls-genetics.github.io/docs/MCS.html.

Further QC was carried out on all datasets to ensure harmonisation in the present study. For full details see supplementary table 8 and for code see https://github.com/AnnaDearman/endophenotype-polygenic-indices/blob/main/genetic_data_qc.sh.

*Pre-imputation QC*

| | UKHLS | NCDS | BCS70 | MCS |
|---|---|---|---|---|
| Genotyping array | Illumina HumanCoreExome-12v1-0 | Illumina (1.2M, Human 660-Quad), Infinium (HumanHap 550K v1.1, HumanHap 550K v3), Affymetrix (v6) | Infinium Global Screening Array-24 v3.0 | Infinium Global Screening Array-24 v1.0 |
| Genotype calling | GenomeStudio (Illumina) GenCall | | GenomeStudio (v2.0, Illumina) | GenomeStudio (v2.0, Illumina) |
| Prior QC software | PLINK1.07, R | PLINK1.9, PLINK2.0, R v3.3.2 and RStudio v4.1.2 | PLINK1.9, PLINK2.0 | PLINK1.9, PLINK2.0 |
| Missing data (individuals) | >10% prefiltering, followed by >2% (59 excluded) | >2% | > 2% (136 excluded) | > 2% (677 excluded) |
| Sex check | Was performed (55 excluded) | X chromosome homozygosity compared to reported sex (females F value > 0.2, males < 0.8) | X chromosome homozygosity compared to reported sex (females F value > 0.2, males < 0.8) | X chromosome homozygosity compared to reported sex (females F value > 0.2, males < 0.8) |

| | | | | |
|---|---|---|---|---|
| | | | (15 excluded) | (86 excluded) |
| Heterozygosity | Performed at MAF≥1% and <1%<br><br>>3 SD<br><br>(148 excluded) | >3 SD from mean | >3 SD from mean<br><br>(46 excluded) | >3 SD from mean<br><br>(78 excluded) |
| Relatedness | Household study, so relatedness is permitted.<br><br>Samples excluded based on incorrect self-reported relatedness.<br><br>Duplicates and potential sample mix-ups excluded based on relatedness metric. | king-cutoff 0.0884 | king-cutoff 0.0884<br><br>(35 excluded) | Trios were sampled, so relatedness is permitted.<br><br>Samples excluded based on parent-child mix-ups. Samples were not excluded based on relatedness. Inbreeding and relatedness (e.g. cousins, twins) was noted. Mendelian error check to verify trios.<br><br>Flag was created to allow users to exclude based on families with multiple mothers and fathers identified (retains one trio per family). |
| Ancestry | Individual outliers removed after estimating principal components of ancestry alongside individuals from multi-ancestry 1000 Genomes panel.<br><br>(340 excluded) | Assessed after imputation. | GenoPred pipeline assigns everyone to a superpopulation. Outliers within groups are defined as >4 SD from mean in the PCs. | GenoPred pipeline assigns everyone to a superpopulation. Outliers within groups are defined as >4 SD from mean in the PCs. Note: MCS was over-sampled to increase representation of different ethnic groups. |
| Missing data (SNPs) | >10% prefiltering, followed by >2% | >3% | >3% | >3% |
| HWE | P < 1x10-4 | P < 1x10-6 | P < 1x10-6 | P < 1x10-6 |
| MAF filtering | Not performed as genotyping array is enriched for rare variants | <1% | <1% | <1% |
| Checked against reference panel for strand issues | HRC-1000G-check-bim.pl (version unspecified) | HRC reference panel r1.1 site list: HRC-1000G- | HRC reference panel r1.1 site list: HRC- | HRC reference panel r1.1 site list: |

| | | | | |
|---|---|---|---|---|
| | | check-bim.pl, v.4.3.0 | 1000G-check-bim.pl, v.4.3.0 | HRC-1000G-check-bim.pl, v.4.3.0 |
| Phasing | SHAPEIT (v2.r) | Eagle2 | Eagle2 | Eagle2 |
| Imputation algorithm | IMPUTE2 (v2.3.0) | Minimac4 | Minimac4 | Minimac4 |
| Imputation panel | UK10K plus 1000 Genomes phase 3 | TOPMed | TOPMed | TOPMed |

Supplementary table 6. Pre-imputation quality control (QC) performed prior to this study. BCS70 = the 1970 British Cohort Study, HWE = Hardy Weinberg Equilibrium test, HRC = Haplotype Reference Consortium, MAF = minor allele frequency, MCS = the Millennium Cohort Study, NCDS = the 1958 National Child Development Study, PCs = principal components, SD = standard deviation, SNPs = single nucleotide polymorphisms, UKHLS = the UK Household Longitudinal Study (Understanding Society).

*Post-imputation QC*

| | UKHLS | NCDS | BCS70 | MCS |
|---|---|---|---|---|
| Imputation confidence | IMPUTE info score < 0.4 | R2 INFO score < 0.8 | R2 INFO score < 0.8 | R2 INFO score < 0.8 |
| Missing data (individuals) | | >2% | >2% | >2% |
| Number of alleles | | >2 alleles (--max-allele 2) | >2 alleles (--max-allele 2) | >2 alleles (--max-allele 2) |
| Missing data (SNPs) | | >3% | >3% | >3% |
| HWE | P < 1x10-4 | P < 1x10-6 | P < 1x10-6 | |
| MAF | | <1% | <1% | |
| Relatedness | | king-cutoff 0.0884<br>(23 excluded) | | |
| Ancestry outliers | | GenoPred pipeline, >4 SD from mean in PC1<br>(72 excluded from GenoPred + PCA analyses) | | |
| Build | 37 | 38 (step not detailed) | 38 | 38 |

Supplementary table 7. Post-imputation quality control (QC) performed prior to this study. BCS70 = the 1970 British Cohort Study, HWE = Hardy Weinberg Equilibrium test, MAF = minor allele frequency, MCS = the Millennium Cohort Study, NCDS = the 1958 National Child Development Study, PC = principal component, PCA = principal component analysis, SD = standard deviation, SNPs = single nucleotide polymorphisms, UKHLS = the UK Household Longitudinal Study (Understanding Society).

The UKHLS data was converted to PLINK format using PLINK1. PLINK2 was used for all subsequent file set updates. Dummy participants were removed from UKHLS, and CLS-supplied flags were used to remove a small number of non-European individuals from BCS70, and to remove twins and families with multiple mothers and fathers from MCS. Duplicate variants were removed based on genomic coordinates, and the majority of rsIDs were assigned using the avsnp151 database, a version of dbSNP 151 reformatted by the ANNOVAR software team (2). The remaining variants were assigned rsIDs by recovering those present in the original file sets: UKHLS (build 37), and NCDS for the three birth cohorts (all build 38), after which duplicate variants were removed based on ID. UKHLS was then merged into one file set, and additionally filtered for variants with long IDs of 20+ characters, which include multiple character-separated rsIDs, and indels with long alleles in the name. Non-rsID variants were removed, having a negligible impact on NCDS and removing 2.1 - 3.0% of variants from the other datasets. Missingness checks were then performed to ensure no variant or individual had >1% missing data - this step removed over five million variants from UKHLS, but no other variants or individuals were removed from any datasets. To avoid the impact of related trios on some of the following QC steps, MCS was then split into a children file set and a parents file set. Variants were removed based on Hardy-Weinberg equilibrium testing, which had a negligible impact on UKHLS and NCDS but removed >3,300 variants from BCS70, >331,000 from MCS children and >540,000 from MCS parents. To calculate relatedness and heterozygosity, temporary file sets were created wherein variants were pruned for linkage disequilibrium (LD). Regions of high LD were removed from the pruned datasets according to scripts “highLDregions4bim_b[build number].awk” downloaded from Jonathan Coleman’s GitHub repository https://github.com/JoniColeman/gwas_scripts. PLINK was then used to estimate relatedness (using the “king” metric) and heterozygosity (F statistic). Using respective cutoffs of 0.0884 (third-degree relatives) and 0.2, I removed 643 individuals from UKHLS, none from NCDS, 3 from BCS70, 18 MCS children and 59 MCS parents. Of these 723 individuals, 669 were removed based on relatedness. Variants with MAF <1% were removed. A separate UKHLS dataset was created which was filtered with a more relaxed MAF threshold (0.5%), retaining 519,927 variants (dataset hereafter referred to as UKHLS-rare). The final variant-filtering step restricted each dataset (aside from UKHLS-rare) to the variants which were present in all datasets, resulting in 3,673,101 variants. Separate European-only file sets were then generated for MCS children and parents using the flag supplied by CLS (the version including non-Europeans will hereafter be referred to as MCS-full).

*QC performed in the present study*

| | UKHLS | NCDS | BCS70 | MCS |
|---|---|---|---|---|
| Convert to PLINK format (using PLINK1)<br>**--gen**<br>**--oxford-single-chr**<br>**--make-bed** | Step performed for UKHLS only | NA | NA | NA |
| Initial numbers of variants and individuals | Variants:<br>24,727,148<br>Individuals:<br>9,961 | Variants:<br>7,545,708<br>Individuals:<br>6,324 | Variants:<br>8,625,760<br>Individuals:<br>5,598 | Variants:<br>8,720,874<br>Individuals:<br>20,247 |
| Manually edit placeholder sample IDs for dummy samples in UKHLS .fam files | Step performed for UKHLS only | NA | NA | NA |
| Remove dummy samples<br>**--remove**<br>*.participants* | Individuals: 9,921 (-40) | NA | NA | NA |

| | | | | |
|---|---|---|---|---|
| Restrict BCS70 to European ancestry<br>**--remove**<br>*.EUR* | NA | NA | Individuals: 5,210 (-388) | (Performed later for MCS) |
| Remove individuals identified by pre-supplied flags “king_related_families” and “singleton_twin”<br>**--remove**<br>*.trios_notwins* | NA | NA | NA | Individuals: 19,729 (-518) |
| Deduplicate based on coordinates (first convert variant ID column to coordinate string, then remove duplicates based on this ID)<br>**--set-all-var-ids @:#[b37]** or **--set-all-var-ids @:#[b38]** as appropriate<br>*.allcoord*<br>**--rm-dup exclude-all**<br>*.dedupcoord* | Variants:<br>24,697,826<br>(-29,322) | Variants:<br>7,543,600<br>(-2,108) | Variants:<br>8,618,026<br>(-7,734) | Variants:<br>8,713,777<br>(-7,097) |
| Assign rsIDs using ANNOVAR. For each dataset, make ANNOVAR query file based on .bim, query avsnp151 database using hg19 for UKHLS and hg38 for others - the resulting file from ANNOVAR has suffix “_dropped” which is then deduplicated, then if the returned ID is the same as the queried ID the variant is removed. The resulting file (.avsnp151) is then used to update variant IDs in PLINK<br>**--update-name**<br>*.avsnp151*<br>Remove duplicate variants after dbSNP ID update<br>**--rm-dup exclude-all**<br>*.deduprsids1* | | | | |
| Top up variant IDs with those from the older versions of UKHLS and NCDS. First, overwrite the original data files with as many from avsnp151 as possible<br>**--recover-var-ids partial**<br>*.avsnp151*<br>Then apply the variant IDs from these updated original files (containing mostly rsIDs from avsnp151 but also some originally-supplied rsIDs) to the latest data files<br>**--recover-var-ids partial** for UKHLS and NCDS<br>**--recover-var-ids partial strict-bim-order** for BCS70 and MCS<br>*.avsnp151origids* | | | | |
| Deduplicate based on original variant IDs<br>**--rm-dup exclude-all**<br>*.deduprsids2* | Variants:<br>24,697,228<br>(-598) | Variants:<br>7,543,592<br>(-8) | Variants:<br>8,618,014<br>(-12) | Variants:<br>8,713,765<br>(-12) |

| | | | | |
|---|---|---|---|---|
| Merge chromosomes into one file and remove any duplicates from across chromosomes<br>**--pmerge-list --merge-mode nm-match**<br>*.deduprsids2pre*<br>**--rm-dup exclude-all**<br>*.deduprsids2* | Variants:<br>24,695,228<br>(no change) | NA | NA | NA |
| Variants removed if they have long IDs (20 or more characters) i.e. character-separated multiple rsIDs or indels with alleles in the name<br>**--exclude**<br>*.nolongids* | Variants:<br>24,585,368<br>(-111,860) | Variants:<br>7,543,592<br>(-0) | Variants:<br>8,618,014<br>(-0) | Variants:<br>8,713,765<br>(-0) |
| All non-rsID variants removed<br>***--exclude***<br>*.rsids* | Variants:<br>24,065,402<br>(-519,966) | Variants:<br>7,543,398<br>(-194) | Variants:<br>8,369,166<br>(-248,848) | Variants:<br>8,450,560<br>(-263,205) |
| Remove variants with missingness above 1%<br>**--geno 0.01**<br>*.lowmissvar* | Variants:<br>18,675,766<br>(-5,389,636) | Variants:<br>7,543,398<br>(no change) | Variants:<br>8,369,166<br>(no change) | Variants:<br>8,450,560<br>(no change) |
| Remove individuals with missingness above 1%<br>**--mind 0.01**<br>*.lowmiss* | Individuals:<br>9,921<br>(no change) | Individuals:<br>6,324<br>(no change) | Individuals:<br>5,210<br>(no change) | Individuals:<br>19,729<br>(no change) |
| Split MCS into children and parents<br>**--keep**<br>**--remove** | NA | NA | NA | Children: 7,597<br>Parents: 12,132 |
| Remove variants which do not pass Hardy Weinberg Equilibrium test<br>**--hwe 0.00001**<br>*.lowmiss.hwe* | Variants:<br>18,675,587<br>(-179) | Variants:<br>7,543,337<br>(-61) | Variants:<br>8,365,839<br>(-3,327) | Children: Variants:<br>8,119,302<br>(-331,258)<br>Parents: Variants:<br>7,910,113<br>(-540,447) |
| Create a copy of each dataset with variants pruned for LD (with a view to calculating relatedness and heterozygosity)<br>**--indep-pairwise 1500 150 0.2** | | | | |

| | | | | |
|---|---|---|---|---|
| *.LD_one*<br>**--extract *LD_one.prune.in**<br>*.LD_two* | | | | |
| Exclude variants with high LD using highLDregions4bim_b37.awk or highLDregions4bim_38.awk<br>**--exclude**<br>*.LD_three* | | | | |
| Identify a list of individuals to exclude due to high relatedness<br>**--king-cutoff 0.0884**<br>*.unrelated.king.cutoff.out.id* | | | | |
| Measure heterozygosity<br>**--het**<br>*.het*<br>Make file of individuals with high heterozygosity<br>*.cohort.highhet* | | | | |
| Make file to remove highly related individuals and those with high heterozygosity, then remove them<br>*.toexclude*<br>**--remove**<br><cohort name> | Variants:<br>18,675,587<br>(no change)<br>Individuals:<br>9,278 (-643, none based on heterozygosity) | Variants:<br>7,543,337<br>(no change)<br>Individuals:<br>6,324 (no change) | Variants:<br>8,365,839<br>(no change)<br>Individuals:<br>5,207 (-3, two based on heterozygosity) | Children: Variants:<br>8,119,302<br>(no change)<br>Individuals:<br>7,579<br>(-18, fifteen based on heterozygosity)<br>Parents: Variants:<br>7,910,113<br>(no change)<br>Individuals:<br>12,073<br>(-59, 37 based on heterozygosity) |
| Remove rare variants<br>**--maf 0.01**<br>*.maf0_01* | Variants:<br>4,542,703<br>(-14,132,884) | Variants:<br>7,543,296<br>(-41) | Variants:<br>8,343,208<br>(-22,631) | Children: Variants:<br>8,092,056<br>(-27,246)<br>Parents: Variants:<br>7,889,764<br>(-20,349) |
| Remove rare variants using a different cutoff for sensitivity analysis | Variants:<br>5,062,630<br>(-13,612,957) | NA | NA | NA |

| | | | | |
|---|---|---|---|---|
| **--maf 0.005**<br>*.maf0_005* | | | | |
| Make file to keep overlapping variants across all files, then restrict to these<br>**--extract**<br>*.harmonised* | maf 0.01: Variants:<br>3,673,101<br>(-869,602) | Variants:<br>3,673,101<br>(-3,870,195) | Variants:<br>3,673,101<br>(-4,670,107) | Children: Variants:<br>3,673,101<br>(-4,418,955)<br>Parents: Variants:<br>3,673,101<br>(-4,237,012) |
| Restrict MCS to European ancestry<br>**--remove**<br>*.european* | NA | NA | NA | Children:<br>6,476<br>(-1,103)<br>Parents:<br>10,484<br>(-1,589) |

Supplementary table 8. Quality control (QC) performed in the current study. Bold text indicates PLINK commands. PLINK2 used except where otherwise specified. Italics represents suffix for new file set. BCS70 = the 1970 British Cohort Study, LD = linkage disequilibrium, MCS = the Millennium Cohort Study, NA = not applicable, NCDS = the 1958 National Child Development Study, UKHLS = Understanding Society a.k.a. the UK Household Longitudinal Study.

*NPGI selection*

Neuropsychiatric polygenic indices (NPGIs) were derived based on studies published on the Psychiatric Genomics Consortium (PGC) website https://pgc.unc.edu/for-researchers/download-results/ [accessed 7th June 2024]. From the 14 sub-headings, ten neuropsychiatric conditions were chosen (anxiety disorders (AD), attention deficit hyperactivity disorder (ADHD), autism spectrum disorder (ASD), bipolar disorder (BD), eating disorders (ED), major depressive disorder (MDD), obsessive-compulsive disorder (OCD), post-traumatic stress disorder (PTSD), schizophrenia (SCZ), substance use disorder (SUD)) and the “cross-disorder” trait, omitting Alzheimer’s Disease, Other and Suicide attempt. There was only one file to choose from for ADHD (3), ASD (4), cross-disorder (5), ED (6) and OCD (7), however the others had multiple options. For AD, the case-control (as opposed to factor score) phenotype was chosen (8), for BD the “all cases” (i.e. bipolar I and bipolar II) (9) file was chosen, for MDD the version excluding 23andMe was chosen (as the inclusive version is restricted to the top 10,000 variants) (10), and the European versions were chosen for PTSD (11), SCZ (12) and SUD (13).

| Title | Author, year | Neuropsychiatric condition |
|---|---|---|
| Meta-analysis of genome-wide association studies of anxiety disorders | Otowa et al, 2016 | Anxiety disorders (AD) |
| Genome-wide analyses of ADHD identify 27 risk loci, refine the genetic architecture and implicate several cognitive domains | Demontis et al, 2023 | Attention deficit hyperactivity disorder (ADHD) |
| Identification of common genetic risk variants for autism spectrum disorder | Grove et al, 2019 | Autism spectrum disorder (ASD) |
| Genome-wide association study of more than 40,000 bipolar disorder cases provides new insights into the underlying biology | Mullins et al, 2021 | Bipolar disorder (BD) |
| Genome-wide association study identifies eight risk loci and implicates metabo-psychiatric origins for anorexia nervosa | Watson et al, 2019 | Eating disorders (ED) |
| Genome-wide meta-analysis of depression identifies 102 independent variants and highlights the importance of the prefrontal brain regions | Howard et al, 2019 | Major depressive disorder (MDD) |
| Revealing the complex genetic architecture of obsessive-compulsive disorder using meta-analysis | Arnold et al, 2018 | Obsessive-compulsive disorder (OCD) |
| International meta-analysis of PTSD genome-wide association studies identifies sex- and ancestry-specific genetic risk loci | Nievergelt et al, 2019 | Post-traumatic stress disorder (PTSD) |
| Mapping genomic loci implicates genes and synaptic biology in schizophrenia | Trubetskoy et al, 2022 | Schizophrenia (SCZ) |
| Multivariate genome-wide association meta-analysis of over 1 million subjects identifies loci underlying multiple substance use disorders | Hatoum et al, 2023 | Substance use disorder (SUD) |
| Genomic Relationships, Novel Loci, and Pleiotropic Mechanisms across Eight Psychiatric Disorders | Lee et al, 2019 | Cross-disorder |

Supplementary table 9. Genome-wide association studies (GWAS) whose summary statistics were used for deriving polygenic indices of neuropsychiatric conditions.

*Endophenotype PGI selection*

The “All associations v1.0” file was downloaded from the NHGRI-EBI GWAS catalog (57) on 19th December 2024. This file contains one row per SNP-trait association (p <= 0.00001) across 36,676 traits and 6,207 studies (earliest publication 2005), resulting in 692,444 rows. The table was deduplicated to leave one row per study-trait pair (n=47,173). All studies without the word “brain” in the title were removed, leaving 82 studies and 2,793 traits. Based on the “DISEASE.TRAIT” column, traits which were unambiguously measured in plasma or cerebrospinal fluid were removed, leaving 2,421 traits, some of which were listed without a specific tissue of origin. Study titles (and where necessary, corresponding traits) were then screened and out-of-scope studies were removed, specifically where a) the topic is pain sensitivity, ageing, frailty, multiple sclerosis, Alzheimer’s disease, Parkinson’s disease, malignancy, brain injury, stroke, intelligence or education, or b) the traits were out-of-scope, i.e. measured in bodily fluids, or behavioural (depression, opioid use, disorder pairs e.g. "Peptic ulcer disease or schizophrenia (pleiotropy)", sleep behaviour, motor coordination, handedness, executive function, restricted and repetitive behaviours, risk-taking behaviour), leaving 49 studies and 2,298 traits. The 49 papers were then downloaded and the full texts were screened. Four were out-of-scope either because the trait was task-based or focussed on brain health with no mention of neuropsychiatric conditions. A further seven brain health papers were excluded from consideration because they only briefly mention one or more neuropsychiatric conditions. Sixteen studies were excluded because they did not attempt to examine genomic overlap between endophenotypes and relevant behavioural outcomes. Ten studies attempted such analyses but found no significant genomic overlaps, while four others found significant results but the summary statistics are un-signed (i.e. lacking effect sizes) so cannot be used to generate PGIs. One study demonstrated genomic overlap between a measure of brain skew and ASD but the summary statistics were not readily available. The remaining seven studies fulfilled the criteria by demonstrating genomic overlap between a relevant behavioural outcome and a non-behavioural brain endophenotype using signed GWAS summary statistics. See table 1 for details, see supplementary figure 1 for flow chart, and for code see https://github.com/AnnaDearman/endophenotype-polygenic-indices/blob/main/choosing%20endophenotypes.R.

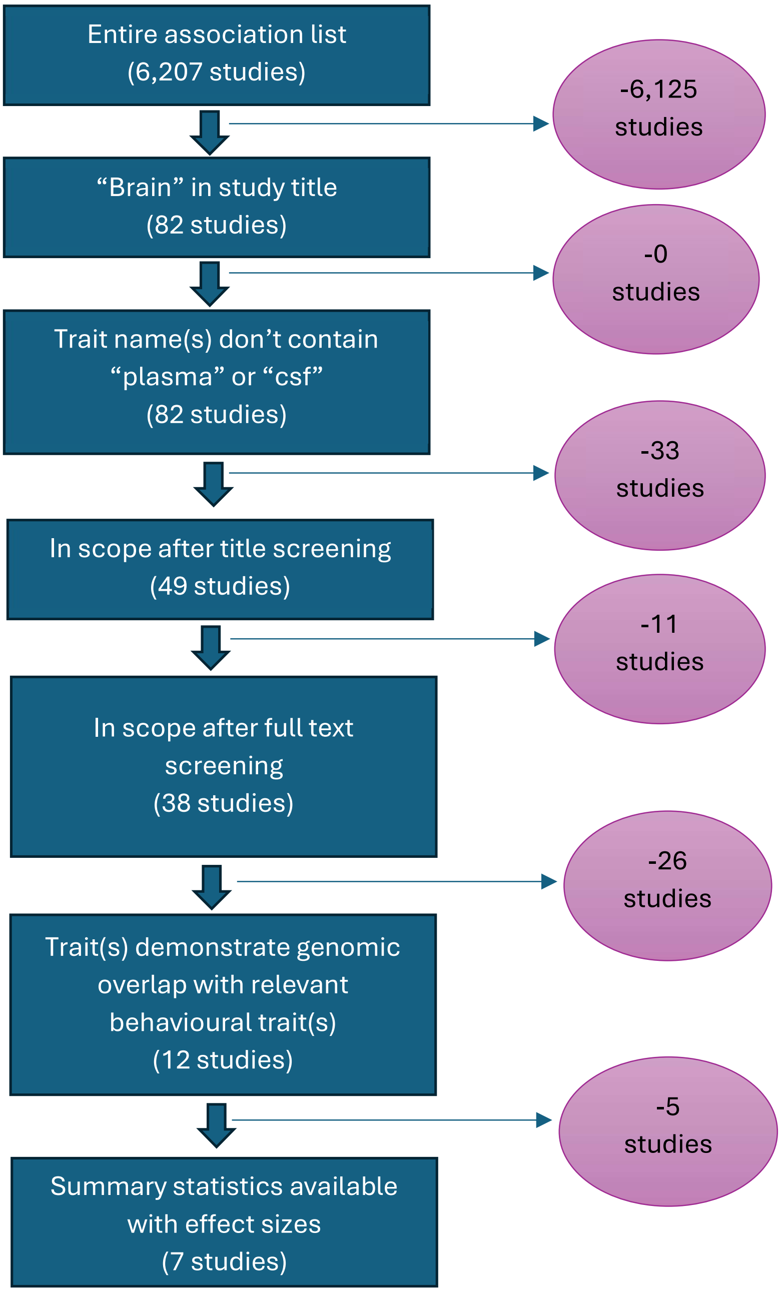


Supplementary figure 1. Flow chart of steps taken to identify genome-wide association studies with suitable summary statistics for endophenotype polygenic indices. “Entire association list” refers to the “All associations v1.0” file downloaded from GWAS catalog on 19th December 2024.

Two study teams provided access to summary statistics after we filled in the corresponding form at https://enigma.ini.usc.edu/research/download-enigma-gwas-results/ (14,15) (due to the sensitivity of the phenotypes in the present study, only the “unrestricted” version was granted for Satizabal et al – this version omits four cohorts), one study author shared the summary statistics download link by e-mail (16), and the remaining four were accessed by following the corresponding links available at the respective GitHub repositories: https://www.med.unc.edu/bigs2/data/gwas-summary-statistics/ (17–19) and https://github.com/norment/open-science/tree/main/2021_Elvsashagen_NatComms_ThalamusGenetics (20).

*Generating PGIs*

*Preparation of summary statistics*

To ensure that PRSice software matched variants based solely on rsIDs, the chromosome and base pair location columns were deleted. Where necessary, long file headers (but not column headers) were trimmed off, effect sizes converted to log-odds-ratios, column headers edited to SNP, A1, A2, P and BETA for consistency, and A1 and A2 columns were switched where necessary based on information in the corresponding README. Fifty rows with missing chromosome information in the SUD summary statistics were removed. For white matter hyperintensities, beta was calculated from the Z score provided.

*PRSice parameters*

Where necessary, a file was passed to the PRSice *--target* parameter which lists the variants to be included for consideration (these files were generated during failed test runs, and contain the suffix ".valid"). The *--binary-target* parameter was set to "T" (i.e. "true"). Default parameters were used for *--clump-kb* (250kb), *--clump-p* (1.0) and *--clump-r2* (0.10). For more information, see code at [https://github.com/AnnaDearman/endophenotype-polygenic-indices/blob/main/polygenic_indices.sh](https://github.com/AnnaDearman/endophenotype-polygenic-indices/blob/main/polygenic_indices.sh).

*Cohort differences*

328 PGI files were generated: 41 phenotypes (11 diagnoses plus 30 endophenotypes) for eight datasets - six with a harmonised set of 3,673,101 variants and similar LD structure (UKHLS, NCDS, BCS70, MCS children, MCS parents), two datasets with harmonised variants but a more diverse LD structure (MCS-full children, MCS-full parents) and one dataset with additional rare variants (UKHLS-rare) = 41 * 8 = 328.

PRSice chose variants to include in each PGI calculation based on the LD structure in the dataset, meaning that cohort differences in LD could result in different SNPs (and different numbers of SNPs) being used, despite filtering for variants present in all datasets. To demonstrate this, the PRSice output files which list the number of variants used (suffix ".prsice") were checked. For each PGI phenotype, across all cohorts, the minimum and maximum numbers of variants used per $P_t$ were identified, and from this a difference was calculated. These minimums, maximums, and differences were then averaged across all phenotypes. This was done once for the European-only datasets, and again after including the ancestrally diverse MCS-full datasets (for self-reported ethnicity in this sample, see supplementary table 26). UKHLS-rare was excluded from these calculations. Across European-only datasets, the average difference was less than 1% of the average minimum, ranging from 0.76 (for $P_t$ 0.0001) to 884.07 (for $P_t$ 1.0). When including the ancestrally diverse MCS-full datasets, average differences are much larger, reaching 3.5% of the average minimum for $P_t$ 1.0 (an average difference of 3,657.46 SNPs). See supplementary table 22 for more info. Hence, differences in PGI construction were greater for the MCS-full samples.

| | Based on European-only samples | | | Including MCS-full samples | | |
|---|---|---|---|---|---|---|
| $P_t$ | Average min n SNPs | Average max n SNPs | Average between-sample difference in n SNPs | Average min n SNPs | Average max n SNPs | Average between-sample difference in n SNPs |
| 0.0001 | 193.68 | 194.44 | 0.76 | 193.85 | 195.73 | 1.88 |
| 0.01 | 4,175.51 | 4,187.05 | 11.54 | 4,177.61 | 4,217.98 | 40.37 |
| 0.05 | 14,044.98 | 14,097.05 | 52.07 | 14,054.71 | 14,256.93 | 202.22 |
| 0.1 | 23,544.71 | 23,648.22 | 103.51 | 23,560.98 | 23,977.56 | 416.59 |
| 0.5 | 71,353.49 | 71,897.42 | 543.93 | 71,411.76 | 73,601.44 | 2,189.68 |
| 1.0 | 98,598.90 | 99,482.98 | 884.07 | 98,706.49 | 102,363.95 | 3,657.46 |

Supplementary table 10. Average numbers of variants included in polygenic index calculations, across phenotypes. Min, max and difference were calculated per phenotype, per $P_t$, across samples. Average min, max and difference across phenotypes (per $P_t$) were then calculated and are shown here. For each $P_t$, the average max n SNPs is higher when the ancestrally diverse MCS-full is included, as is the average between-sample difference in n SNPs. MCS = the Millennium Cohort Study, $P_t$ = p value threshold, SNPs = single nucleotide polymorphisms.

### Sensitivity analyses

Sensitivity analyses were carried out to examine the impacts of inclusion criteria, operationalisation (of polygenic indices (PGIs), mother's education, and symptom scale used), covariate adjustment, model specification and age at symptom measurement. See below and supplementary table 11.

#### *Inclusion criteria*

1. Bereaved participants were included in all samples.
2. Participants of all ancestries were included in MCS.
3. Participants without parental genotype data were excluded - see "Covariate adjustment" point 3.

#### *Operationalisation*

1. PGIs derived in the UKHLS general population sample (GPS) after using MAF filtering of 0.5% were examined .
2. Mother's education was operationalised to include vocational qualifications in BCS70 and MCS.
3. For some waves, NCDS and BCS70 used two different symptom scales contemporaneously. For more detail see "Age at measurement" point 1.

#### *Age at measurement*

1. Four and eight additional analyses were run for NCDS and BCS70, respectively, using mental health symptoms measured at different ages, some of which used alternative scales.

#### *Covariate adjustment*

1. Ten ancestry principal components (PCs) were adjusted for in GPS, for paths all paths with a PGI as an explanatory variable.
2. Covariates age and age$^2$ were omitted from the model in GPS.

3. Parental PGIs were adjusted for in MCS. To examine the impact of subsetting (as this entailed a nontrivial reduction in sample size) these analyses were repeated in the sub-sample without adjustment for parental PGIs (see "Inclusion criteria" point 3).

*Model specification*

1. In the main analysis, path models do not model pairwise correlations between the three exposures. Analyses were repeated including these correlation terms.

| Dataset | MAF filtering | Ancestry | Age at symptom measurement | Symptom scale | Covariate adjustment | Mother's education | Exposure pair correlation terms |
|---|---|---|---|---|---|---|---|
| GPS, **bereaved included** | 1% | European | 16 – 99 (mean 54) | GHQ-12 | age | Secondary | No |
| BHPS, **bereaved included** | 1% | European | 16 – 98 (mean 55) | GHQ-12 | age | Secondary | No |
| NCDS, **bereaved included** | 1% | European | 23 | Malaise-24 | none | Secondary | No |
| BCS70, **bereaved included** | 1% | European | 26 | Malaise-24 | none | Secondary | No |
| MCS, **bereaved included** | 1% | European | 17 | Kessler-6 | none | Secondary | No |
| MCS **– full** | 1% | **All** | 17 | Kessler-6 | none | Secondary | No |
| MCS **– parent PGI sub-sample** | 1% | European | 17 | Kessler-6 | none | Secondary | No |
| GPS **– rare** | **0.5%** | European | 16 – 99 (mean 54) | GHQ-12 | age | Secondary | No |
| BCS70 | 1% | European | 26 | Malaise-24 | none | **Secondary and vocational** | No |
| MCS | 1% | European | 17 | Kessler-6 | none | **Secondary and vocational** | No |
| NCDS | 1% | European | **33, 42, 50** | **Various** | none | Secondary | No |
| BCS70 | 1% | European | **16, 30, 34, 42, 46** | **Various** | none | Secondary | No |
| GPS | 1% | European | 16 – 99 (mean 54) | GHQ-12 | age, **ancestry PCs x10** | Secondary | No |
| GPS | 1% | European | 16 – 99 (mean 54) | GHQ-12 | **none** | Secondary | No |

| | | | | | | | |
|---|---|---|---|---|---|---|---|
| MCS **– parent PGI sub-sample** | 1% | European | 17 | Kessler-6 | **mother's PGI, father's PGI** | Secondary | No |
| GPS | 1% | European | 16 – 99 (mean 54) | GHQ-12 | age | Secondary | **Yes** |
| BHPS | 1% | European | 16 – 98 (mean 55) | GHQ-12 | age | Secondary | **Yes** |
| NCDS | 1% | European | 23 | Malaise-24 | none | Secondary | **Yes** |
| BCS70 | 1% | European | 26 | Malaise-24 | none | Secondary | **Yes** |
| MCS | 1% | European | 17 | Kessler-6 | none | Secondary | **Yes** |

Supplementary table 11. Details of sensitivity analyses. Bold is used to indicate aspects which deviate from the main analyses. BCS70 = the 1970 British Cohort Study, BHPS = British Household Panel Survey, GHQ = general health questionnaire, GPS = general population sample of Understanding Society, MCS = Millennium Cohort Study, NCDS = the 1958 National Child Development Study, PCs = principal components, PGI = polygenic index.

### Supplementary results

#### ***Descriptive statistics***

| | GPS | | BHPS | | NCDS | | BCS70 | | MCS | |
|---|---|---|---|---|---|---|---|---|---|---|
| | Men | Women | Men | Women | Men | Women | Men | Women | Men | Women |
| n (%) | 2,336 (42.9) | 3,115 (57.1) | 723 (44.0) | 920 (56.0) | 2,522 (49.3) | 2,596 (50.7) | 1,093 (43.4) | 1,428 (56.6) | 2,053 (48.4) | 2,187 (51.6) |
| Year of birth (age at biospecimen collection for MCS): mean (SD) range | 1955 (16.0) 1917-1994 | 1957 (15.7) 1915-1994 | 1953 (14.4) 1913-1986 | 1954 (14.8) 1914-1987 | 1958 | 1958 | 1970 | 1970 | 13.7 (0.5) 13-15 | 13.8 (0.5) 13-15 |
| Family intact (%) | 82.4 | 80.9 | 84.0 | 84.0 | 80.9 | 78.0 | 77.8 | 78.9 | 63.4 | 62.8 |
| Father employed (%) | 90.6 | 88.8 | 92.9 | 91.5 | 88.1 | 85.8 | 87.2 | 85.7 | 80.4 | 80.8 |
| Mother educated (%) | 46.7 | 49.0 | 45.8 | 41.1 | 29.7 | 29.1 | 54.2 | 52.6 | 97.3 | 97.3 |

Supplementary table 12. Descriptive statistics for the alternative samples used in sensitivity analyses which include individuals whose parent(s) died during youth. When the sample is expanded to include bereaved participants, family intact necessarily decreases - by around five percentage points for most datasets but by less than one for MCS. Father employed decreases by around four percentage points, except for MCS where it decreases by one. BCS70 = the 1970 British Cohort Study, BHPS = British Household Panel Survey, GPS = general population sample of Understanding Society, MCS = Millennium Cohort Study, NCDS = the 1958 National Child Development Study, SD = standard deviation.

| | MCS - full | | MCS - parent PGIs | |
|---|---|---|---|---|
| | Men | Women | Men | Women |
| n (%) | 2,340 (48.5) | 2,488 (51.5) | 1,072 (48.9) | 1,118 (51.1) |
| Age at biospecimen collection: mean (SD) range | 13.7 (0.5) 13-15 | 13.8 (0.5) 13-15 | 13.7 (0.5) 13-15 | 13.8 (0.5) 13-15 |
| Family intact (%) | 64.9 | 63.9 | 96.1 | 96.9 |
| Father employed (%) | 80.4 | 80.9 | 92.6 | 93.0 |
| Mother educated (%) | 96.5 | 96.5 | 98.4 | 98.8 |
| White (%) | 1,964 (83.9) | 2,099 (84.4) | | |
| Pakistani and Bangladeshi (%) | 118 (5.0) | 125 (5.0) | | |
| Indian (%) | 57 (2.4) | 60 (2.4) | | |
| Black or Black British (%) | 49 (2.1) | 49 (2.0) | | |
| Mixed (%) | 42 (1.8) | 59 (2.4) | | |
| Other (%) | 30 (1.3) | 20 (0.8) | | |
| Missing (%) | 80 (3.4) | 76 (3.1) | | |

Supplementary table 13. Descriptive statistics for the alternative MCS samples used in sensitivity analyses. MCS-full was not filtered for ancestry. MCS-parent PGIs indicates the sub-sample (of the European dataset) with both parents' PGIs available. Rates of the three exposures are similar for MCS-full and its European-only counterpart, in contrast to the MCS-parent PGIs sub-sample, for which rates of all three are higher. MCS – parent PGIs sub-sample is around 52% the size of the main sample, which in turn is around 87% the size of MCS-full. MCS = Millennium Cohort Study, PGIs = polygenic indices, SD = standard deviation.

| | BCS70 | | MCS | |
|---|---|---|---|---|
| | Men | Women | Men | Women |
| Mother qualifications (%) | 65.0 | 63.3 | 99.0 | 99.0 |

Supplementary table 14. Rates of maternal education when operationalised by including vocational qualifications ("mother qualifications"). BCS70 = the 1970 British Cohort Study, MCS = Millennium Cohort Study.

| | NCDS, 33 (malaise) | | NCDS, 42 (psych. distress) | | NCDS, 42 (malaise) | | NCDS, 50 (malaise) | | BCS70, 16 (psych. distress) | | BCS70, 16 (malaise) | | BCS70, 30 (psych. distress) | | BCS70, 30 (malaise) | | BCS70, 34 (psych. distress) | | BCS70, 34 (malaise) | | BCS70, 42 (malaise) | | BCS70, 46 (malaise) | |
|---|---|---|---|---|---|---|---|---|---|---|---|---|---|---|---|---|---|---|---|---|---|---|---|---|
| | M | W | M | W | M | W | M | W | M | W | M | W | M | W | M | W | M | W | M | W | M | W | M | W |
| n (%) | 2,141 (48.8) | 2,243 (51.2) | 2,305 (49.2) | 2,383 (50.8) | 2,302 (49.2) | 2,380 (50.8) | 2,091 (48.8) | 2,196 (51.2) | 488 (36.9) | 834 (63.1) | 546 (37.4) | 914 (62.6) | 959 (42.9) | 1,275 (57.1) | 960 (42.9) | 1,277 (57.1) | 925 (42.8) | 1,235 (57.2) | 925 (42.8) | 1,235 (57.2) | 905 (42.7) | 1,215 (57.3) | 985 (43.2) | 1,297 (56.8) |
| Family intact (%) | 85.4 | 82.5 | 85.2 | 82.3 | 85.1 | 82.3 | 84.9 | 82.5 | 84.8 | 85.1 | 82.9 | 83.5 | 82.8 | 83.5 | 83.5 | 83.6 | 83.5 | 83.6 | 83.5 | 83.6 | 82.7 | 83.9 | 81.8 | 83.4 |
| Father employed (%) | 91.6 | 89.5 | 91.3 | 89.5 | 91.3 | 89.5 | 91.5 | 90 | 89.9 | 89.3 | 91.2 | 89.7 | 91.3 | 89.7 | 91.7 | 89.7 | 91.7 | 89.7 | 91.7 | 89.7 | 91.4 | 89.9 | 91.4 | 89.6 |
| Mother educated (%) | 30.6 | 29.3 | 30.1 | 29.3 | 30.1 | 29.4 | 30.7 | 30.4 | 61.4 | 54.6 | 54.1 | 53.3 | 54.1 | 53.4 | 55 | 53.6 | 55 | 53.6 | 55 | 53.6 | 54.1 | 53.7 | 54.7 | 53.8 |

Supplementary table 15. Descriptive statistics for the alternative samples used in sensitivity analyses which examine associations with mental health measured at different ages. Sample sizes vary for the NCDS and BCS70 samples which include mental health measures at different ages. Multiple symptom scales were used at some waves, with differential missingness except for BCS70 at age 34. Sample sizes are low for the age 16 analyses in BCS70 especially for men, who also seem to have higher rates of maternal education compared to other time points, or compared to women. BCS70 = the 1970 British Cohort Study, M = men, NCDS = the 1958 National Child Development Study, W = women.

***PGI correlations***

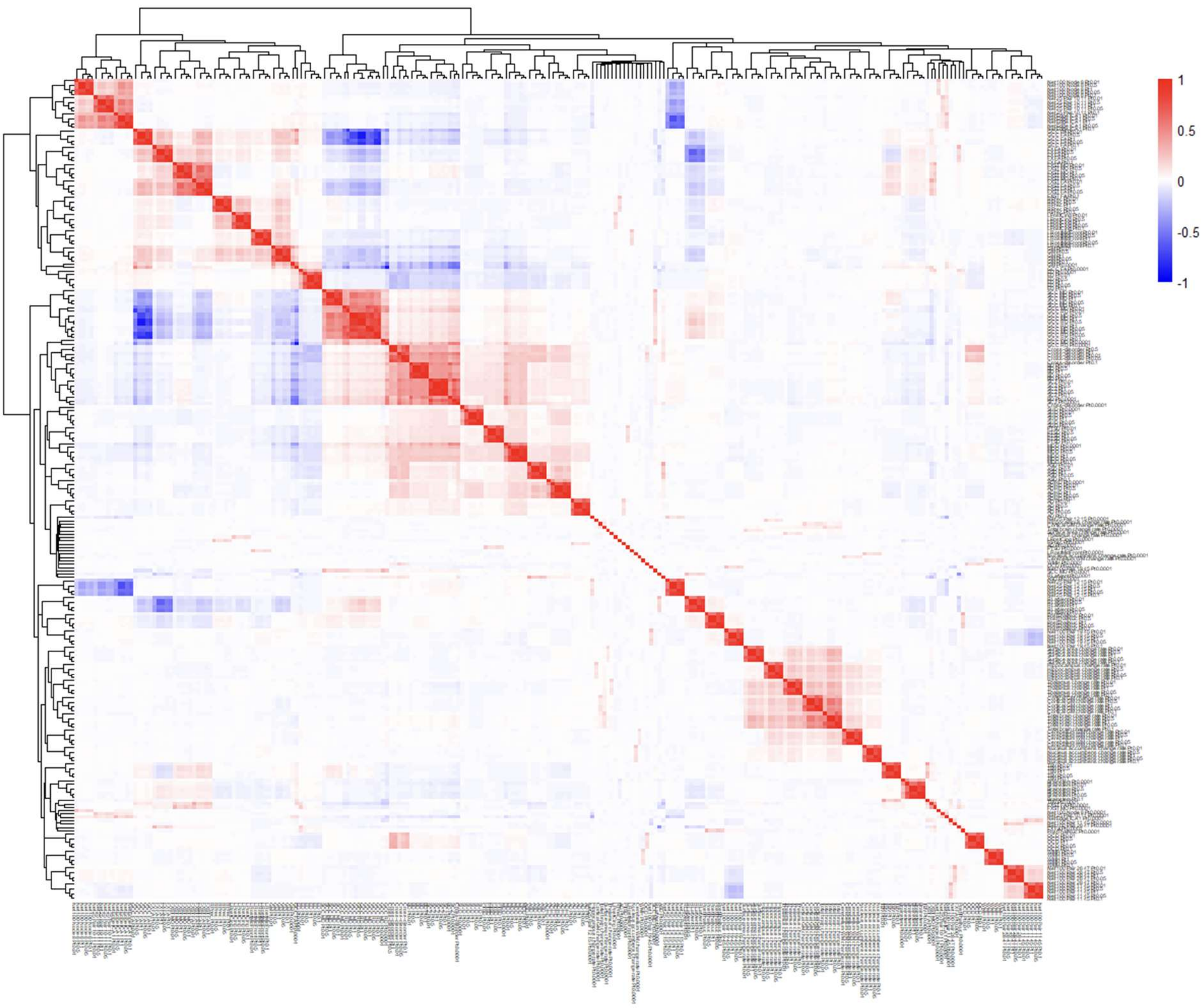

Supplementary figure 2. Full correlation heatmap of all polygenic indices (PGIs). All samples (except UKHLS-rare and MCS-full) and all $P_t$s included. 5x5 squares of high correlation tend to indicate single traits, with each row and column indicating a different $P_t$ from 0.01 to 1.0 (PGIs with $P_t$ 0.0001 tend to be less collinear with their trait counterparts). These 5x5 squares tend to cluster with those of similar traits, forming larger squares e.g. 15x15 for three resting-state brain activity traits in the top left corner.

### ***Exposure associations with psychological distress***

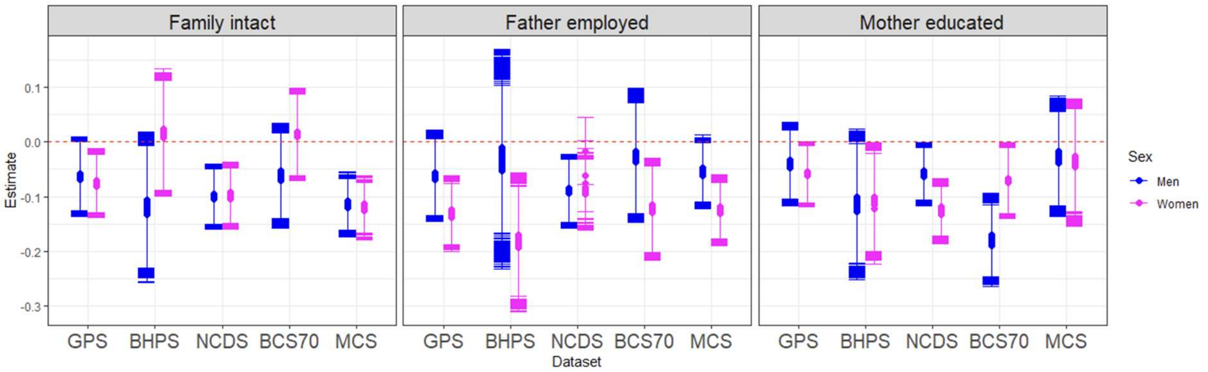


Supplementary figure 3. Exposure associations with psychological distress as estimated in multiple path models, each with a different polygenic index, hence the slight variability in superimposed estimates. Error bars represent 95% confidence intervals. Negative coefficients indicate that the exposure is protective. Compared to linear regression models, additional associations are observed for family intact (GPS women), father employed (NCDS men) and mother educated (BCS70 women). BCS70 = the 1970 British Cohort Study, BHPS = British Household Panel Survey, GPS = general population sample of Understanding Society, MCS = Millennium Cohort Study, NCDS = the 1958 National Child Development Study.

| Dataset | Sex | Family intact | Father employed | Mother educated |
|---|---|---|---|---|
| GPS | Women | **0.009 (0.006-0.019)** | **0.000 (0.000-0.000)** | 0.036 (0.030-0.053) |
| | Men | 0.069 (0.050-0.093) | 0.104 (0.068-0.157) | 0.238 (0.182-0.345) |
| BHPS | Women | 0.812 (0.684-0.883) | **0.002 (0.001-0.004)** | **0.029 (0.019-0.046)** |
| | Men | 0.066 (0.040-0.095) | 0.731 (0.572-0.905) | 0.070 (0.045-0.112) |
| NCDS | Women | **0.000 (0.000-0.001)** | 0.008 (0.004-0.595) | **0.000 (0.000-0.000)** |
| | Men | **0.000 (0.000-0.001)** | **0.006 (0.003-0.008)** | **0.026 (0.018-0.047)** |
| BCS70 | Women | 0.760 (0.641-0.828) | **0.005 (0.003-0.009)** | **0.034 (0.023-0.044)** |
| | Men | 0.148 (0.110-0.242) | 0.638 (0.511-0.777) | **0.000 (0.000-0.000)** |
| MCS | Women | **0.000 (0.000-0.000)** | **0.000 (0.000-0.000)** | 0.517 (0.397-0.628) |
| | Men | **0.000 (0.000-0.000)** | 0.063 (0.048-0.121) | 0.544 (0.424-0.750) |

Supplementary table 16. P value mean (range) across main analysis path models, for exposures' associations with psychological distress. Bold indicates significant associations (max $p < 0.05$). Compared to linear regression models, additional associations are observed for family intact (GPS women), father employed (NCDS men) and mother educated (BCS70 women). BCS70 = the 1970 British Cohort Study, BHPS = British Household Panel Survey, GPS = general population sample of Understanding Society, MCS = Millennium Cohort Study, NCDS = the 1958 National Child Development Study.

| Dataset | Sex | Family intact | Father employed | Mother educated |
|---|---|---|---|---|
| GPS | Women | **0.014 (0.010-0.027)** | **0.000 (0.000-0.000)** | **0.016 (0.013-0.024)** |
| | Men | 0.132 (0.112-0.165) | 0.298 (0.223-0.364) | 0.207 (0.163-0.299) |
| BHPS | Women | 0.721 (0.637-0.779) | **0.013 (0.010-0.018)** | 0.051 (0.033-0.079) |
| | Men | 0.105 (0.072-0.131) | 0.608 (0.499-0.702) | 0.084 (0.053-0.130) |
| NCDS | Women | **0.000 (0.000-0.001)** | **0.003 (0.002-0.009)** | **0.000 (0.000-0.000)** |
| | Men | **0.002 (0.001-0.003)** | **0.018 (0.013-0.025)** | **0.008 (0.006-0.016)** |
| BCS70 | Women | 0.631 (0.541-0.749) | **0.001 (0.000-0.002)** | **0.033 (0.022-0.047)** |
| | Men | 0.192 (0.156-0.277) | 0.483 (0.356-0.592) | **0.000 (0.000-0.000)** |
| MCS | Women | **0.000 (0.000-0.000)** | **0.000 (0.000-0.000)** | 0.526 (0.403-0.634) |
| | Men | **0.000 (0.000-0.000)** | 0.098 (0.073-0.184) | 0.447 (0.341-0.645) |

Supplementary table 17. Analyses including bereaved participants. P value mean (range) across path models, for exposures' associations with psychological distress. Bold indicates significant associations (max p < 0.05). Compared to main analysis path models, additional associations are observed for father employed (NCDS women) and mother educated (GPS women) while the association for mother educated is lost in BHPS women. BCS70 = the 1970 British Cohort Study, BHPS = British Household Panel Survey, GPS = general population sample of Understanding Society, MCS = Millennium Cohort Study, NCDS = the 1958 National Child Development Study.

| Sensitivity analysis | Sex | Family intact | Father employed | Mother educated |
|---|---|---|---|---|
| MCS – full (all ancestries) | Women | **0.000 (0.000-0.000)** | **0.000 (0.000-0.000)** | 0.851 (0.759-0.997) |
| | Men | **0.000 (0.000-0.000)** | 0.128 (0.086-0.195) | 0.421 (0.306-0.534) |
| MCS – parent PGI sub-sample | Women | 0.311 (0.236-0.412) | **0.001 (0.000-0.002)** | 0.129 (0.032-0.171) |
| | Men | 0.090 (0.038-0.124) | 0.676 (0.540-0.798) | 0.834 (0.639-0.956) |
| MCS – parent PGIs adjusted | Women | 0.314 (0.234-0.418) | **0.001 (0.000-0.001)** | 0.129 (0.039-0.173) |
| | Men | 0.094 (0.062-0.152) | 0.673 (0.538-0.846) | 0.832 (0.693-0.952) |

Supplementary table 18. MCS-specific sensitivity analyses examining the impact of inclusion criteria, and adjustment for parental PGIs. P value mean (range) across path models, for exposures' associations with psychological distress. Bold indicates significant associations (max p < 0.05). Compared to main analysis path models, using the parent PGI sub-sample removed the association with family intact. MCS = Millennium Cohort Study, PGI = polygenic index.

| Sensitivity analysis | Sex | Family intact | Father employed | Mother educated |
|---|---|---|---|---|
| Rarer variants included in PGIs | Women | **0.009 (0.006-0.019)** | **0.000 (0.000-0.000)** | 0.036 (0.028-0.051) |
| | Men | 0.068 (0.048-0.088) | 0.103 (0.069-0.156) | 0.237 (0.182-0.339) |
| Adjusted for ancestry PCs | Women | **0.010 (0.007-0.020)** | **0.000 (0.000-0.000)** | 0.045 (0.039-0.063) |
| | Men | 0.048 (0.034-0.074) | 0.092 (0.058-0.153) | 0.255 (0.196-0.357) |
| No age adjustment | Women | **0.009 (0.006-0.021)** | **0.000 (0.000-0.000)** | 0.376 (0.332-0.441) |
| | Men | **0.030 (0.024-0.050)** | 0.120 (0.089-0.186) | 0.627 (0.558-0.718) |

Supplementary table 19. GPS-specific sensitivity analyses. P value mean (range) across path models, for exposures' associations with psychological distress. Bold indicates significant associations (max p < 0.05). Compared to main analysis path models, the only difference observed is that lack of age adjustment induces an association for family intact in GPS men. GPS = general population sample of Understanding Society, PCs = principal components, PGIs = polygenic indices.

| Dataset | Sex | Family intact | Father employed | Mother educated |
|---|---|---|---|---|
| BCS70 | Women | 0.760 (0.641-0.828) | **0.005 (0.003-0.009)** | **0.001 (0.001-0.003)** |
| | Men | 0.148 (0.110-0.242) | 0.638 (0.511-0.777) | **0.000 (0.000-0.000)** |
| MCS | Women | **0.000 (0.000-0.000)** | **0.000 (0.000-0.000)** | 0.144 (0.076-0.193) |
| | Men | **0.000 (0.000-0.000)** | 0.063 (0.048-0.121) | 0.841 (0.687-0.985) |

Supplementary table 20. Use of alternative measure of mother's education which includes vocational qualifications. P value mean (range) across path models, for exposures' associations with psychological distress. Bold indicates significant associations (max p < 0.05). BCS70 = the 1970 British Cohort Study, MCS = Millennium Cohort Study.

| Dataset, age (outcome) | Sex | Family intact | Father employed | Mother educated |
|---|---|---|---|---|
| NCDS, 33 (malaise) | Women | 0.152 (0.127-0.260) | 0.070 (0.048-0.137) | **0.000 (0.000-0.000)** |
| NCDS, 33 (malaise) | Men | **0.002 (0.002-0.003)** | 0.052 (0.032-0.073) | **0.003 (0.002-0.007)** |
| NCDS, 42 (psychological distress) | Women | 0.050 (0.042-0.080) | 0.388 (0.348-0.553) | 0.968 (0.784-1.000) |
| NCDS, 42 (psychological distress) | Men | **0.021 (0.016-0.027)** | 0.568 (0.500-0.629) | 0.047 (0.027-0.060) |
| NCDS, 42 (malaise) | Women | 0.115 (0.095-0.176) | 0.080 (0.063-0.144) | **0.000 (0.000-0.000)** |
| NCDS, 42 (malaise) | Men | **0.000 (0.000-0.000)** | 0.733 (0.350-0.837) | **0.012 (0.009-0.023)** |
| NCDS, 50 (malaise) | Women | 0.061 (0.044-0.085) | 0.101 (0.075-0.154) | 0.063 (0.048-0.111) |
| NCDS, 50 (malaise) | Men | **0.000 (0.000-0.000)** | 0.105 (0.072-0.143) | 0.070 (0.055-0.102) |
| BCS70, 16 (psychological distress) | Women | 0.471 (0.390-0.573) | **0.010 (0.005-0.019)** | 0.322 (0.280-0.395) |
| BCS70, 16 (psychological distress) | Men | **0.003 (0.001-0.007)** | 0.198 (0.135-0.265) | 0.445 (0.343-0.730) |
| BCS70, 16 (malaise) | Women | 0.851 (0.746-0.978) | **0.004 (0.002-0.007)** | 0.547 (0.486-0.639) |
| BCS70, 16 (malaise) | Men | 0.726 (0.643-1.000) | 0.688 (0.585-0.933) | 0.467 (0.393-0.573) |
| BCS70, 30 (psychological distress) | Women | 0.667 (0.602-0.754) | 0.354 (0.300-0.430) | 0.648 (0.560-0.702) |
| BCS70, 30 (psychological distress) | Men | 0.216 (0.171-0.306) | 0.745 (0.659-0.853) | 0.365 (0.311-0.491) |
| BCS70, 30 (malaise) | Women | 0.349 (0.284-0.404) | 0.338 (0.274-0.401) | 0.435 (0.358-0.483) |
| BCS70, 30 (malaise) | Men | 0.028 (0.019-0.056) | 0.380 (0.319-0.475) | **0.000 (0.000-0.000)** |
| BCS70, 34 (psychological distress) | Women | 0.779 (0.666-0.837) | 0.065 (0.042-0.101) | 0.360 (0.299-0.419) |
| BCS70, 34 (psychological distress) | Men | 0.204 (0.157-0.306) | 0.152 (0.108-0.215) | 0.455 (0.372-0.710) |
| BCS70, 34 (malaise) | Women | 0.703 (0.618-0.815) | **0.019 (0.013-0.033)** | 0.373 (0.261-0.443) |
| BCS70, 34 (malaise) | Men | 0.072 (0.049-0.122) | 0.835 (0.633-0.955) | 0.370 (0.321-0.521) |
| BCS70, 42 (malaise) | Women | 0.924 (0.823-0.993) | 0.839 (0.746-0.978) | 0.153 (0.109-0.179) |
| BCS70, 42 (malaise) | Men | 0.731 (0.650-0.848) | 0.422 (0.340-0.498) | 0.123 (0.102-0.178) |
| BCS70, 46 (malaise) | Women | 0.131 (0.090-0.170) | 0.213 (0.163-0.359) | 0.091 (0.060-0.134) |
| BCS70, 46 (malaise) | Men | 0.057 (0.038-0.096) | 0.143 (0.101-0.190) | 0.834 (0.721-1.000) |

Supplementary table 21. Mental health measured at different ages. P value mean (range) across path models, for exposures' associations with psychological distress. Bold indicates significant associations (max p < 0.05). When measuring mental health in NCDS at ages 33, 42 and 50, family intact remained protective in men but became non-significant in women. Father employed became non-significant in both, while mother educated was protective in both sexes at ages 33 and 42 (malaise only) but not 50. For ages 16, 30, 34, 42 and 46 in BCS70, family intact remained largely non-significant, except in men at age 16 (psychological distress but not malaise) and age 30 (malaise but not psychological distress), father employed remained protective in women at ages 16 (both outcomes) and 34 (malaise only), while mother educated became non-significant, except in men at age 30 (malaise only). BCS70 = the 1970 British Cohort Study, NCDS = the 1958 National Child Development Study.

| Dataset | Sex | Family intact | Father employed | Mother educated |
|---|---|---|---|---|
| GPS | Women | 0.925 (0.853-0.994) | **0.007 (0.003-0.010)** | 0.257 (0.229-0.344) |
| | Men | 0.434 (0.368-0.529) | 0.570 (0.478-0.664) | 0.388 (0.319-0.489) |
| BHPS | Women | 0.089 (0.057-0.108) | **0.001 (0.001-0.002)** | 0.048 (0.030-0.065) |
| | Men | 0.080 (0.047-0.105) | 0.771 (0.600-0.964) | 0.117 (0.078-0.164) |
| NCDS | Women | 0.155 (0.001-0.186) | 0.888 (0.018-1.000) | **0.000 (0.000-0.000)** |
| | Men | 0.126 (0.098-0.196) | 0.844 (0.684-0.904) | 0.043 (0.033-0.079) |
| BCS70 | Women | 0.616 (0.503-0.682) | **0.015 (0.011-0.025)** | 0.271 (0.218-0.302) |
| | Men | 0.167 (0.123-0.281) | 0.559 (0.469-0.643) | **0.000 (0.000-0.000)** |
| MCS | Women | 0.151 (0.124-0.204) | 0.078 (0.067-0.119) | 0.767 (0.665-0.927) |
| | Men | **0.005 (0.004-0.010)** | 0.480 (0.408-0.563) | 0.907 (0.778-1.000) |

Supplementary table 22. Pairwise correlation terms between exposures were modelled in path models. P value mean (range) across path models, for exposures' associations with psychological distress. Bold indicates significant associations (max $p < 0.05$). Specifying pairwise correlation terms between exposures had a drastic effect on the associations between exposures and psychological distress in path models: family intact lost four out of five associations, father employed lost two out of five, and mother educated lost three out of five. BCS70 = the 1970 British Cohort Study, BHPS = British Household Panel Survey, GPS = general population sample of Understanding Society, MCS = Millennium Cohort Study, NCDS = the 1958 National Child Development Study, UKHLS = the UK Household Longitudinal Study (Understanding Society).

***PGI associations with psychological distress***

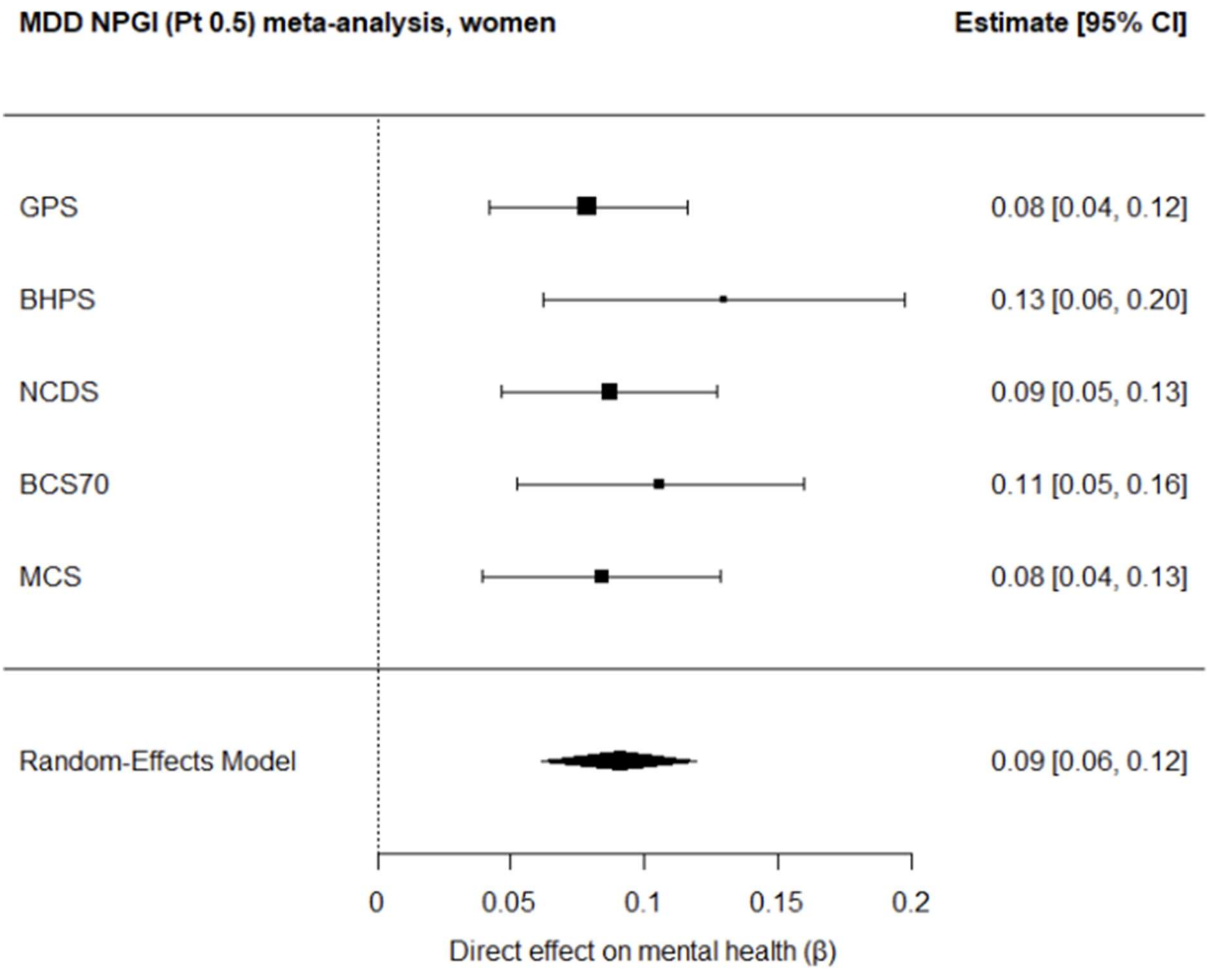


Supplementary figure 4. Meta-analysis plot for depression PGI ($P_t$ 0.5) in women. BCS70 = the 1970 British Cohort Study, BHPS = British Household Panel Survey, CI = confidence interval, GPS = general population sample of Understanding Society, MCS = Millennium Cohort Study, MDD = major depressive disorder, NCDS = the 1958 National Child Development Study, NPGI = neuropsychiatric polygenic index, $P_t$ = p value threshold.

| | Men | | | Women | | |
|---|---|---|---|---|---|---|
| PGI | Beta | P | Hetero geneity P | Beta | P | Hetero geneity P |
| NPGIs | | | | | | |
| AD Pt 0.0001 | 0.0188 | 0.1550 | 0.8294 | 0.0049 | 0.6592 | 0.7892 |
| AD Pt 0.01 | 0.0234 | 0.3741 | **0.0119** | 0.0177 | 0.1596 | 0.7509 |
| AD Pt 0.05 | 0.0266 | 0.4177 | **0.0005** | 0.0248 | 0.0764 | 0.9695 |
| AD Pt 0.1 | 0.0329 | 0.2617 | **0.0037** | 0.0244 | 0.0765 | 0.9587 |
| AD Pt 0.5 | 0.0297 | 0.2449 | **0.0117** | 0.0189 | 0.1389 | 0.5652 |
| AD Pt 1 | 0.0253 | 0.2773 | **0.0221** | 0.0191 | 0.1362 | 0.4926 |
| ADHD Pt 0.0001 | 0.0272 | 0.1094 | 0.3299 | 0.0413 | **0.0153** | 0.7574 |
| ADHD Pt 0.01 | 0.0504 | **0.0274** | 0.1968 | 0.0622 | **0.0042** | 0.8320 |
| ADHD Pt 0.05 | 0.0468 | **0.0143** | 0.9046 | 0.0646 | **0.0036** | 0.8563 |
| ADHD Pt 0.1 | 0.0390 | **0.0262** | 0.9295 | 0.0630 | **0.0041** | 0.9575 |
| ADHD Pt 0.5 | 0.0468 | **0.0147** | 0.7786 | 0.0531 | **0.0074** | 0.9258 |
| ADHD Pt 1 | 0.0483 | **0.0133** | 0.7255 | 0.0533 | **0.0073** | 0.8486 |
| ASD Pt 0.0001 | 0.0180 | 0.1814 | 0.4477 | 0.0195 | 0.1815 | 0.2790 |
| ASD Pt 0.01 | 0.0201 | 0.2377 | 0.2587 | 0.0106 | 0.3575 | 0.5139 |
| ASD Pt 0.05 | 0.0294 | 0.1619 | 0.0860 | 0.0183 | 0.2343 | 0.1831 |
| ASD Pt 0.1 | 0.0300 | 0.1383 | 0.1347 | 0.0205 | 0.3065 | **0.0303** |
| ASD Pt 0.5 | 0.0232 | 0.1863 | 0.2586 | 0.0203 | 0.2853 | **0.0474** |
| ASD Pt 1 | 0.0250 | 0.1586 | 0.2736 | 0.0192 | 0.3380 | **0.0281** |
| BD Pt 0.0001 | 0.0153 | 0.2443 | 0.5889 | 0.0067 | 0.5609 | 0.3159 |
| BD Pt 0.01 | 0.0207 | 0.1462 | 0.2587 | 0.0163 | 0.1986 | 0.8387 |
| BD Pt 0.05 | 0.0281 | 0.3117 | **0.0082** | 0.0231 | 0.0950 | 0.9053 |
| BD Pt 0.1 | 0.0341 | 0.1622 | **0.0332** | 0.0239 | 0.0859 | 0.8732 |
| BD Pt 0.5 | 0.0408 | **0.0225** | 0.2718 | 0.0255 | 0.0739 | 0.8597 |
| BD Pt 1 | 0.0398 | **0.0243** | 0.1811 | 0.0238 | 0.0882 | 0.8803 |
| Cross-disorder Pt 0.0001 | 0.0336 | **0.0413** | 0.7943 | 0.0048 | 0.6714 | 0.6541 |
| Cross-disorder Pt 0.01 | 0.0459 | **0.0147** | 0.6598 | 0.0214 | 0.1086 | 0.4548 |
| Cross-disorder Pt 0.05 | 0.0534 | **0.0402** | 0.0656 | 0.0333 | **0.0321** | 0.6319 |
| Cross-disorder Pt 0.1 | 0.0495 | **0.0119** | 0.1616 | 0.0363 | **0.0240** | 0.8031 |
| Cross-disorder Pt 0.5 | 0.0467 | **0.0149** | 0.3121 | 0.0411 | **0.0160** | 0.4586 |
| Cross-disorder Pt 1 | 0.0482 | **0.0137** | 0.2437 | 0.0437 | **0.0133** | 0.4924 |
| ED Pt 0.0001 | -0.0179 | 0.1814 | 0.5622 | 0.0020 | 0.8580 | 0.5812 |
| ED Pt 0.01 | -0.0302 | 0.0540 | 0.6964 | -0.0051 | 0.6416 | 0.2848 |
| ED Pt 0.05 | -0.0387 | **0.0254** | 0.5888 | -0.0069 | 0.5432 | 0.5021 |
| ED Pt 0.1 | -0.0412 | **0.0208** | 0.5813 | -0.0028 | 0.7980 | 0.3063 |
| ED Pt 0.5 | -0.0297 | 0.0762 | 0.2964 | 0.0037 | 0.7947 | 0.1030 |
| ED Pt 1 | -0.0280 | 0.0769 | 0.3380 | 0.0044 | 0.7299 | 0.1203 |
| MDD Pt 0.0001 | 0.0668 | **0.0038** | 0.4463 | 0.0434 | **0.0243** | 0.2452 |
| MDD Pt 0.01 | 0.0802 | **0.0021** | 0.8060 | 0.0665 | **0.0031** | 0.8258 |
| MDD Pt 0.05 | 0.0836 | **0.0017** | 0.9558 | 0.0816 | **0.0014** | 0.8467 |
| MDD Pt 0.1 | 0.0893 | **0.0014** | 0.9704 | 0.0843 | **0.0013** | 0.8776 |

| | | | | | | |
|---|---|---|---|---|---|---|
| MDD Pt 0.5 | 0.0828 | **0.0019** | 0.9981 | 0.0907 | **0.0010** | 0.7174 |
| MDD Pt 1 | 0.0827 | **0.0019** | 0.9971 | 0.0894 | **0.0010** | 0.6791 |
| OCD Pt 0.0001 | 0.0235 | 0.1015 | 0.8421 | 0.0117 | 0.3176 | 0.9751 |
| OCD Pt 0.01 | 0.0245 | 0.2684 | **0.0339** | 0.0073 | 0.5202 | 0.9722 |
| OCD Pt 0.05 | 0.0233 | 0.2994 | **0.0200** | 0.0029 | 0.7985 | 0.8917 |
| OCD Pt 0.1 | 0.0200 | 0.4256 | **0.0047** | 0.0043 | 0.7015 | 0.9116 |
| OCD Pt 0.5 | 0.0253 | 0.1713 | 0.1329 | -0.0029 | 0.7989 | 0.7866 |
| OCD Pt 1 | 0.0231 | 0.2178 | 0.1065 | -0.0025 | 0.8269 | 0.7521 |
| PTSD Pt 0.0001 | -0.0144 | 0.3585 | 0.2447 | -0.0202 | 0.1225 | 0.5986 |
| PTSD Pt 0.01 | 0.0279 | 0.0673 | 0.7012 | 0.0238 | 0.0828 | 0.4232 |
| PTSD Pt 0.05 | 0.0258 | 0.0839 | 0.7740 | 0.0336 | 0.0634 | 0.2378 |
| PTSD Pt 0.1 | 0.0320 | **0.0455** | 0.9618 | 0.0284 | 0.1497 | 0.0644 |
| PTSD Pt 0.5 | 0.0400 | **0.0238** | 0.9975 | 0.0388 | 0.0669 | 0.0877 |
| PTSD Pt 1 | 0.0401 | **0.0240** | 0.9951 | 0.0382 | 0.0688 | 0.0917 |
| SCZ Pt 0.0001 | 0.0338 | **0.0402** | 0.8433 | -0.0002 | 0.9833 | 0.3358 |
| SCZ Pt 0.01 | 0.0377 | **0.0273** | 0.6255 | 0.0132 | 0.2715 | 0.4244 |
| SCZ Pt 0.05 | 0.0268 | 0.0732 | 0.7557 | 0.0138 | 0.3252 | 0.1995 |
| SCZ Pt 0.1 | 0.0313 | **0.0490** | 0.8539 | 0.0157 | 0.2036 | 0.5920 |
| SCZ Pt 0.5 | 0.0370 | **0.0296** | 0.6713 | 0.0182 | 0.1522 | 0.3687 |
| SCZ Pt 1 | 0.0364 | **0.0309** | 0.6142 | 0.0184 | 0.1475 | 0.3897 |
| SUD Pt 0.0001 | -0.0018 | 0.8789 | 0.2800 | 0.0067 | 0.5544 | 0.7493 |
| SUD Pt 0.01 | 0.0145 | 0.5971 | **0.0053** | 0.0111 | 0.3491 | 0.8091 |
| SUD Pt 0.05 | 0.0235 | 0.1789 | 0.1203 | 0.0141 | 0.2488 | 0.9436 |
| SUD Pt 0.1 | 0.0240 | 0.1042 | 0.2681 | 0.0088 | 0.4465 | 0.8746 |
| SUD Pt 0.5 | 0.0288 | 0.0628 | 0.1284 | 0.0087 | 0.4457 | 0.9379 |
| SUD Pt 1 | 0.0284 | 0.1007 | 0.1117 | 0.0086 | 0.4566 | 0.9689 |
| EPGIs – brain volumes | | | | | | |
| TBV Pt 0.0001 | -0.0079 | 0.5190 | 0.7509 | -0.0167 | 0.3730 | **0.0453** |
| TBV Pt 0.01 | -0.0318 | **0.0462** | 0.5462 | -0.0273 | 0.1723 | **0.0380** |
| TBV Pt 0.05 | -0.0395 | **0.0259** | 0.4116 | -0.0275 | 0.1680 | **0.0460** |
| TBV Pt 0.1 | -0.0378 | **0.0271** | 0.5388 | -0.0302 | 0.1010 | 0.1245 |
| TBV Pt 0.5 | -0.0350 | **0.0348** | 0.7526 | -0.0246 | 0.1342 | 0.2161 |
| TBV Pt 1 | -0.0331 | **0.0410** | 0.8201 | -0.0229 | 0.1437 | 0.2680 |
| Total GM Pt 0.0001 | -0.0135 | 0.3903 | 0.2459 | -0.0082 | 0.4794 | 0.3606 |
| Total GM Pt 0.01 | 0.0017 | 0.9273 | 0.0691 | -0.0240 | 0.1577 | 0.1768 |
| Total GM Pt 0.05 | -0.0063 | 0.6882 | 0.1797 | -0.0178 | 0.1911 | 0.3670 |
| Total GM Pt 0.1 | -0.0046 | 0.8014 | 0.0738 | -0.0226 | 0.1343 | 0.2808 |
| Total GM Pt 0.5 | -0.0031 | 0.8817 | **0.0299** | -0.0262 | 0.1391 | 0.1246 |
| Total GM Pt 1 | -0.0032 | 0.8734 | **0.0388** | -0.0247 | 0.1309 | 0.1862 |
| LPostCing Pt 0.0001 | -0.0066 | 0.5903 | 0.7657 | -0.0099 | 0.4618 | 0.2274 |
| LPostCing Pt 0.01 | 0.0082 | 0.5072 | 0.8271 | -0.0090 | 0.4593 | 0.4177 |
| LPostCing Pt 0.05 | 0.0062 | 0.6050 | 0.8596 | -0.0175 | 0.1643 | 0.3393 |
| LPostCing Pt 0.1 | 0.0071 | 0.5545 | 0.5679 | -0.0184 | 0.1471 | 0.4169 |
| LPostCing Pt 0.5 | 0.0004 | 0.9754 | 0.9401 | -0.0136 | 0.2547 | 0.8057 |

| | | | | | | |
|---|---|---|---|---|---|---|
| LPostCing Pt 1 | 0.0036 | 0.7527 | 0.8520 | -0.0121 | 0.3067 | 0.7744 |
| LRostMidFront Pt 0.0001 | -0.0222 | 0.1211 | 0.6320 | 0.0028 | 0.8012 | 0.8986 |
| LRostMidFront Pt 0.01 | -0.0034 | 0.8823 | **0.0172** | 0.0134 | 0.2711 | 0.8765 |
| LRostMidFront Pt 0.05 | -0.0043 | 0.8783 | **0.0022** | 0.0024 | 0.8240 | 0.8406 |
| LRostMidFront Pt 0.1 | -0.0024 | 0.9354 | **0.0027** | 0.0018 | 0.8852 | 0.3288 |
| LRostMidFront Pt 0.5 | 0.0123 | 0.7436 | **0.0000** | -0.0006 | 0.9616 | 0.4658 |
| LRostMidFront Pt 1 | 0.0119 | 0.7417 | **0.0001** | -0.0011 | 0.9198 | 0.5244 |
| RLatVent Pt 0.0001 | -0.0035 | 0.7715 | 0.8898 | 0.0145 | 0.2390 | 0.9025 |
| RLatVent Pt 0.01 | -0.0069 | 0.6081 | 0.3784 | 0.0036 | 0.7477 | 0.9418 |
| RLatVent Pt 0.05 | -0.0072 | 0.5523 | 0.7282 | 0.0054 | 0.6314 | 0.6817 |
| RLatVent Pt 0.1 | 0.0043 | 0.7169 | 0.4666 | 0.0057 | 0.6190 | 0.9463 |
| RLatVent Pt 0.5 | 0.0093 | 0.4435 | 0.5593 | 0.0034 | 0.7623 | 0.8857 |
| RLatVent Pt 1 | 0.0090 | 0.4864 | 0.4709 | 0.0028 | 0.8052 | 0.9183 |
| RPrec Pt 0.0001 | -0.0163 | 0.2227 | 0.6798 | 0.0067 | 0.6494 | 0.1609 |
| RPrec Pt 0.01 | 0.0036 | 0.7621 | 0.4688 | 0.0014 | 0.9023 | 0.5149 |
| RPrec Pt 0.05 | 0.0067 | 0.6574 | 0.1651 | 0.0025 | 0.8233 | 0.8576 |
| RPrec Pt 0.1 | 0.0079 | 0.6248 | 0.1281 | -0.0048 | 0.6674 | 0.8410 |
| RPrec Pt 0.5 | 0.0041 | 0.7306 | 0.3448 | -0.0069 | 0.5504 | 0.4322 |
| RPrec Pt 1 | 0.0022 | 0.8488 | 0.3960 | -0.0053 | 0.6345 | 0.4642 |
| Brainstem Pt 0.0001 | 0.0202 | 0.2994 | 0.0671 | -0.0194 | 0.1396 | 0.8508 |
| Brainstem Pt 0.01 | -0.0013 | 0.9131 | 0.8781 | -0.0130 | 0.2830 | 0.9724 |
| Brainstem Pt 0.05 | 0.0016 | 0.9323 | 0.0942 | -0.0141 | 0.2489 | 0.9758 |
| Brainstem Pt 0.1 | 0.0034 | 0.8564 | 0.0852 | -0.0092 | 0.4233 | 0.8990 |
| Brainstem Pt 0.5 | 0.0097 | 0.5524 | 0.1833 | -0.0068 | 0.5480 | 0.8725 |
| Brainstem Pt 1 | 0.0084 | 0.5635 | 0.3208 | -0.0083 | 0.4723 | 0.8452 |
| PostThalNuc Pt 0.0001 | 0.0332 | 0.1387 | **0.0490** | 0.0097 | 0.4233 | 0.3946 |
| PostThalNuc Pt 0.01 | 0.0172 | 0.3626 | 0.0784 | 0.0197 | 0.1423 | 0.3753 |
| PostThalNuc Pt 0.05 | 0.0109 | 0.6001 | **0.0303** | 0.0172 | 0.1682 | 0.9915 |
| PostThalNuc Pt 0.1 | 0.0042 | 0.7757 | 0.2309 | 0.0137 | 0.2510 | 0.9752 |
| PostThalNuc Pt 0.5 | 0.0020 | 0.8660 | 0.5036 | 0.0141 | 0.2409 | 0.9661 |
| PostThalNuc Pt 1 | 0.0003 | 0.9800 | 0.4908 | 0.0162 | 0.1872 | 0.8581 |
| EPGIs – white matter microstructure | | | | | | |
| FXST FA Pt 0.0001 | -0.0207 | 0.1322 | 0.6682 | -0.0014 | 0.8982 | 0.6711 |
| FXST FA Pt 0.01 | -0.0191 | 0.1575 | 0.8463 | -0.0047 | 0.6802 | 0.8605 |
| FXST FA Pt 0.05 | -0.0131 | 0.3717 | 0.2361 | -0.0114 | 0.3416 | 0.4086 |
| FXST FA Pt 0.1 | -0.0115 | 0.5671 | **0.0251** | -0.0067 | 0.5604 | 0.7249 |
| FXST FA Pt 0.5 | -0.0113 | 0.5505 | **0.0407** | -0.0096 | 0.4090 | 0.8477 |
| FXST FA Pt 1 | -0.0126 | 0.5257 | **0.0288** | -0.0070 | 0.5417 | 0.6768 |
| GCC FA Pt 0.0001 | -0.0096 | 0.4476 | 0.6493 | 0.0109 | 0.3719 | 0.3718 |
| GCC FA Pt 0.01 | -0.0158 | 0.2291 | 0.5303 | -0.0226 | 0.1565 | 0.1635 |
| GCC FA Pt 0.05 | -0.0177 | 0.1821 | 0.8891 | -0.0168 | 0.2581 | 0.1697 |
| GCC FA Pt 0.1 | -0.0151 | 0.2402 | 0.8060 | -0.0167 | 0.3302 | 0.0978 |
| GCC FA Pt 0.5 | -0.0138 | 0.2728 | 0.9417 | -0.0206 | 0.1364 | 0.2943 |
| GCC FA Pt 1 | -0.0151 | 0.2377 | 0.9583 | -0.0172 | 0.2135 | 0.2532 |
| FX FA Pt 0.0001 | 0.0210 | 0.1529 | 0.3891 | -0.0007 | 0.9523 | 0.7443 |

| | | | | | | |
|---|---|---|---|---|---|---|
| FX FA Pt 0.01 | 0.0074 | 0.5405 | 0.9199 | -0.0064 | 0.5742 | 0.6732 |
| FX FA Pt 0.05 | 0.0171 | 0.1947 | 0.6289 | -0.0011 | 0.9220 | 0.5835 |
| FX FA Pt 0.1 | 0.0187 | 0.1556 | 0.7219 | 0.0002 | 0.9867 | 0.7483 |
| FX FA Pt 0.5 | 0.0153 | 0.2275 | 0.9485 | -0.0015 | 0.8902 | 0.4553 |
| FX FA Pt 1 | 0.0139 | 0.2654 | 0.9697 | -0.0037 | 0.7421 | 0.4589 |
| GCC MD Pt 0.0001 | 0.0186 | 0.1651 | 0.4560 | 0.0053 | 0.7565 | 0.0651 |
| GCC MD Pt 0.01 | 0.0187 | 0.1636 | 0.6977 | 0.0269 | 0.0622 | 0.7318 |
| GCC MD Pt 0.05 | 0.0178 | 0.1790 | 0.9295 | 0.0147 | 0.2315 | 0.6964 |
| GCC MD Pt 0.1 | 0.0163 | 0.2160 | 0.9170 | 0.0212 | 0.1106 | 0.6653 |
| GCC MD Pt 0.5 | 0.0230 | 0.1051 | 0.8029 | 0.0224 | 0.0969 | 0.4964 |
| GCC MD Pt 1 | 0.0220 | 0.1168 | 0.8186 | 0.0236 | 0.0858 | 0.4704 |
| SCC MD Pt 0.0001 | 0.0178 | 0.1875 | 0.5735 | -0.0191 | 0.1435 | 0.7954 |
| SCC MD Pt 0.01 | 0.0245 | 0.2250 | 0.0677 | 0.0007 | 0.9479 | 0.4374 |
| SCC MD Pt 0.05 | 0.0118 | 0.5807 | **0.0388** | 0.0111 | 0.3387 | 0.6593 |
| SCC MD Pt 0.1 | 0.0152 | 0.2514 | 0.2060 | 0.0146 | 0.2308 | 0.4691 |
| SCC MD Pt 0.5 | 0.0198 | 0.3153 | 0.0645 | 0.0135 | 0.2571 | 0.8324 |
| SCC MD Pt 1 | 0.0195 | 0.3257 | 0.0621 | 0.0144 | 0.2334 | 0.8269 |
| FXST MO Pt 0.0001 | -0.0135 | 0.2789 | 0.8501 | -0.0150 | 0.2199 | 0.5999 |
| FXST MO Pt 0.01 | -0.0088 | 0.4706 | 0.3919 | 0.0028 | 0.8497 | 0.1386 |
| FXST MO Pt 0.05 | -0.0067 | 0.6370 | 0.1939 | -0.0124 | 0.3016 | 0.4578 |
| FXST MO Pt 0.1 | -0.0149 | 0.3239 | 0.2417 | -0.0057 | 0.6095 | 0.4574 |
| FXST MO Pt 0.5 | -0.0084 | 0.4870 | 0.5715 | -0.0036 | 0.7499 | 0.5358 |
| FXST MO Pt 1 | -0.0084 | 0.5027 | 0.4841 | -0.0025 | 0.8247 | 0.5527 |
| GCC RD Pt 0.0001 | 0.0120 | 0.3625 | 0.6568 | -0.0135 | 0.4879 | **0.0304** |
| GCC RD Pt 0.01 | 0.0211 | 0.1341 | 0.4136 | 0.0260 | 0.0670 | 0.3564 |
| GCC RD Pt 0.05 | 0.0198 | 0.1412 | 0.9681 | 0.0218 | 0.1641 | 0.1704 |
| GCC RD Pt 0.1 | 0.0119 | 0.3365 | 0.9781 | 0.0158 | 0.2767 | 0.1687 |
| GCC RD Pt 0.5 | 0.0158 | 0.2238 | 0.9786 | 0.0159 | 0.3076 | 0.1406 |
| GCC RD Pt 1 | 0.0139 | 0.2739 | 0.9684 | 0.0203 | 0.1544 | 0.2141 |
| EPGIs – white matter lesions | | | | | | |
| WMH Pt 0.0001 | 0.0149 | 0.2467 | 0.4675 | -0.0071 | 0.5363 | 0.9683 |
| WMH Pt 0.01 | 0.0011 | 0.9292 | 0.9107 | -0.0122 | 0.3955 | 0.1847 |
| WMH Pt 0.05 | 0.0039 | 0.7754 | 0.2202 | -0.0019 | 0.8665 | 0.5272 |
| WMH Pt 0.1 | 0.0004 | 0.9717 | 0.1816 | -0.0031 | 0.7822 | 0.7500 |
| WMH Pt 0.5 | 0.0004 | 0.9705 | 0.2577 | 0.0033 | 0.7714 | 0.9168 |
| WMH Pt 1 | 0.0000 | 0.9974 | 0.1962 | 0.0043 | 0.7051 | 0.8775 |
| EPGIs – resting-state brain activity | | | | | | |
| Net Edge ICA1 Pt 0.0001 | 0.0138 | 0.2898 | 0.5904 | -0.0046 | 0.6777 | 0.7305 |
| Net Edge ICA1 Pt 0.01 | -0.0232 | 0.1272 | 0.2319 | -0.0010 | 0.9285 | 0.7176 |
| Net Edge ICA1 Pt 0.05 | -0.0171 | 0.1969 | 0.7807 | -0.0065 | 0.5707 | 0.8919 |
| Net Edge ICA1 Pt 0.1 | -0.0106 | 0.3946 | 0.9991 | -0.0130 | 0.2919 | 0.9848 |
| Net Edge ICA1 Pt 0.5 | -0.0054 | 0.6443 | 0.9881 | -0.0110 | 0.3587 | 0.9156 |
| Net Edge ICA1 Pt 1 | -0.0038 | 0.7454 | 0.9781 | -0.0103 | 0.3858 | 0.8938 |
| Net 100 Node 8 Pt 0.0001 | 0.0003 | 0.9770 | 0.6134 | -0.0014 | 0.8946 | 0.6069 |
| Net 100 Node 8 Pt 0.01 | 0.0031 | 0.7909 | 0.8421 | -0.0109 | 0.4621 | 0.1725 |

| | | | | | | |
|---|---|---|---|---|---|---|
| Net 100 Node 8 Pt 0.05 | 0.0081 | 0.5049 | 0.5909 | -0.0160 | 0.2010 | 0.9773 |
| Net 100 Node 8 Pt 0.1 | 0.0048 | 0.7465 | 0.2691 | -0.0163 | 0.1939 | 0.6237 |
| Net 100 Node 8 Pt 0.5 | 0.0055 | 0.7065 | 0.2776 | -0.0221 | 0.0990 | 0.7343 |
| Net 100 Node 8 Pt 1 | 0.0066 | 0.6535 | 0.2645 | -0.0214 | 0.1066 | 0.7647 |
| Net 100 Pair 11 45 Pt 0.0001 | 0.0119 | 0.4750 | 0.1059 | -0.0200 | 0.1987 | 0.2045 |
| Net 100 Pair 11 45 Pt 0.01 | 0.0160 | 0.2301 | 0.3004 | -0.0151 | 0.2661 | 0.2903 |
| Net 100 Pair 11 45 Pt 0.05 | 0.0193 | 0.3468 | 0.0563 | -0.0226 | 0.1790 | 0.1294 |
| Net 100 Pair 11 45 Pt 0.1 | 0.0213 | 0.1419 | 0.2491 | -0.0225 | 0.1880 | 0.1471 |
| Net 100 Pair 11 45 Pt 0.5 | 0.0177 | 0.1982 | 0.3887 | -0.0292 | 0.1093 | 0.1382 |
| Net 100 Pair 11 45 Pt 1 | 0.0184 | 0.1865 | 0.3486 | -0.0312 | 0.1040 | 0.1103 |
| Net 100 Pair 19 45 Pt 0.0001 | -0.0046 | 0.7023 | 0.9201 | -0.0139 | 0.4826 | **0.0248** |
| Net 100 Pair 19 45 Pt 0.01 | -0.0104 | 0.4363 | 0.2910 | 0.0041 | 0.7047 | 0.8697 |
| Net 100 Pair 19 45 Pt 0.05 | -0.0032 | 0.8156 | 0.2584 | 0.0017 | 0.9236 | 0.0520 |
| Net 100 Pair 19 45 Pt 0.1 | -0.0017 | 0.8898 | 0.2574 | 0.0074 | 0.6845 | **0.0490** |
| Net 100 Pair 19 45 Pt 0.5 | -0.0111 | 0.3903 | 0.3329 | 0.0150 | 0.4319 | **0.0390** |
| Net 100 Pair 19 45 Pt 1 | -0.0117 | 0.3685 | 0.3477 | 0.0176 | 0.3738 | **0.0320** |
| Net 100 Pair 28 47 Pt 0.0001 | 0.0094 | 0.4330 | 0.6394 | -0.0160 | 0.2967 | 0.1440 |
| Net 100 Pair 28 47 Pt 0.01 | -0.0053 | 0.6553 | 0.5617 | -0.0127 | 0.2928 | 0.7792 |
| Net 100 Pair 28 47 Pt 0.05 | -0.0130 | 0.2964 | 0.7123 | -0.0138 | 0.2976 | 0.3678 |
| Net 100 Pair 28 47 Pt 0.1 | -0.0092 | 0.4518 | 0.5013 | -0.0199 | 0.2472 | 0.1362 |
| Net 100 Pair 28 47 Pt 0.5 | -0.0043 | 0.7148 | 0.3427 | -0.0211 | 0.2534 | 0.0834 |
| Net 100 Pair 28 47 Pt 1 | -0.0077 | 0.5237 | 0.3734 | -0.0213 | 0.2136 | 0.1374 |
| Net 25 Pair 10 11 Pt 0.0001 | -0.0219 | 0.1700 | 0.2394 | -0.0004 | 0.9701 | 0.9782 |
| Net 25 Pair 10 11 Pt 0.01 | 0.0001 | 0.9936 | 0.1087 | -0.0118 | 0.3080 | 0.9474 |
| Net 25 Pair 10 11 Pt 0.05 | 0.0000 | 0.9970 | 0.3163 | -0.0196 | 0.1238 | 0.6063 |
| Net 25 Pair 10 11 Pt 0.1 | 0.0027 | 0.8462 | 0.2477 | -0.0097 | 0.3947 | 0.6313 |
| Net 25 Pair 10 11 Pt 0.5 | 0.0001 | 0.9927 | 0.1612 | -0.0004 | 0.9684 | 0.5455 |
| Net 25 Pair 10 11 Pt 1 | 0.0026 | 0.8519 | 0.2873 | -0.0004 | 0.9736 | 0.5134 |
| Net 25 Pair 12 15 Pt 0.0001 | -0.0144 | 0.2729 | 0.7175 | -0.0061 | 0.6031 | 0.4218 |
| Net 25 Pair 12 15 Pt 0.01 | -0.0029 | 0.8016 | 0.6087 | 0.0064 | 0.5781 | 0.5867 |
| Net 25 Pair 12 15 Pt 0.05 | -0.0007 | 0.9538 | 0.7439 | -0.0016 | 0.8821 | 0.7396 |
| Net 25 Pair 12 15 Pt 0.1 | -0.0033 | 0.7781 | 0.8290 | -0.0091 | 0.4273 | 0.7992 |
| Net 25 Pair 12 15 Pt 0.5 | -0.0013 | 0.9084 | 0.9748 | -0.0068 | 0.5436 | 0.8895 |
| Net 25 Pair 12 15 Pt 1 | -0.0013 | 0.9127 | 0.9605 | -0.0070 | 0.5372 | 0.8808 |
| EPGIs – longitudinal brain structure | | | | | | |
| Cerebellum WM change rate Pt 0.0001 | -0.0084 | 0.4922 | 0.7648 | 0.0122 | 0.2991 | 0.4497 |
| Cerebellum WM change rate Pt 0.01 | -0.0006 | 0.9692 | 0.1300 | -0.0108 | 0.3642 | 0.8054 |
| Cerebellum WM change rate Pt 0.05 | -0.0056 | 0.6425 | 0.5354 | -0.0171 | 0.3136 | 0.1003 |
| Cerebellum WM change rate Pt 0.1 | -0.0002 | 0.9911 | 0.1926 | -0.0276 | 0.0567 | 0.6804 |
| Cerebellum WM change rate Pt 0.5 | 0.0010 | 0.9439 | 0.2818 | -0.0363 | **0.0243** | 0.9267 |
| Cerebellum WM change rate Pt 1 | 0.0025 | 0.8523 | 0.3543 | -0.0361 | **0.0247** | 0.8798 |
| Surface area change rate Pt 0.0001 | -0.0123 | 0.3341 | 0.6105 | -0.0175 | 0.1697 | 0.9251 |
| Surface area change rate Pt 0.01 | -0.0157 | 0.2679 | 0.4429 | -0.0140 | 0.3385 | 0.1631 |
| Surface area change rate Pt 0.05 | -0.0181 | 0.4104 | **0.0218** | -0.0113 | 0.3411 | 0.7734 |
| Surface area change rate Pt 0.1 | -0.0159 | 0.3741 | 0.1345 | -0.0042 | 0.7092 | 0.5526 |

| | | | | | | |
|---|---|---|---|---|---|---|
| Surface area change rate Pt 0.5 | -0.0192 | 0.2586 | 0.2181 | -0.0028 | 0.8035 | 0.3381 |
| Surface area change rate Pt 1 | -0.0184 | 0.2454 | 0.3047 | -0.0041 | 0.7496 | 0.2169 |
| Cortical GM change rate Pt 0.0001 | -0.0309 | 0.0886 | 0.1136 | -0.0033 | 0.7639 | 0.9175 |
| Cortical GM change rate Pt 0.01 | -0.0044 | 0.7212 | 0.5500 | -0.0095 | 0.4166 | 0.5834 |
| Cortical GM change rate Pt 0.05 | -0.0151 | 0.2442 | 0.7380 | 0.0065 | 0.5723 | 0.4796 |
| Cortical GM change rate Pt 0.1 | -0.0126 | 0.4028 | 0.2602 | 0.0056 | 0.6173 | 0.9584 |
| Cortical GM change rate Pt 0.5 | -0.0177 | 0.2493 | 0.2987 | 0.0032 | 0.7757 | 0.6387 |
| Cortical GM change rate Pt 1 | -0.0164 | 0.2611 | 0.3506 | 0.0034 | 0.7575 | 0.6663 |
| Thalamus change rate Pt 0.0001 | 0.0140 | 0.3463 | 0.3943 | -0.0121 | 0.5018 | **0.0419** |
| Thalamus change rate Pt 0.01 | 0.0060 | 0.6865 | 0.2609 | -0.0125 | 0.3057 | 0.5089 |
| Thalamus change rate Pt 0.05 | 0.0083 | 0.5057 | 0.4895 | -0.0152 | 0.3369 | 0.1660 |
| Thalamus change rate Pt 0.1 | 0.0085 | 0.5447 | 0.3410 | -0.0201 | 0.2195 | 0.1832 |
| Thalamus change rate Pt 0.5 | 0.0002 | 0.9889 | 0.9027 | -0.0186 | 0.1514 | 0.6033 |
| Thalamus change rate Pt 1 | 0.0014 | 0.9044 | 0.9562 | -0.0182 | 0.1587 | 0.5518 |
| Hippocampus change rate Pt 0.0001 | 0.0123 | 0.3422 | 0.8634 | -0.0004 | 0.9784 | 0.2888 |
| Hippocampus change rate Pt 0.01 | -0.0053 | 0.6560 | 0.8527 | -0.0105 | 0.4706 | 0.1693 |
| Hippocampus change rate Pt 0.05 | 0.0044 | 0.7144 | 0.9357 | -0.0102 | 0.3902 | 0.4391 |
| Hippocampus change rate Pt 0.1 | -0.0089 | 0.4706 | 0.7615 | -0.0128 | 0.2930 | 0.6125 |
| Hippocampus change rate Pt 0.5 | -0.0124 | 0.3230 | 0.9943 | -0.0146 | 0.2304 | 0.6872 |
| Hippocampus change rate Pt 1 | -0.0135 | 0.2876 | 0.9802 | -0.0138 | 0.2559 | 0.6358 |
| Total brain change rate Pt 0.0001 | -0.0021 | 0.8590 | 0.8813 | 0.0075 | 0.5185 | 0.4969 |
| Total brain change rate Pt 0.01 | -0.0033 | 0.8340 | 0.1696 | 0.0067 | 0.6069 | 0.2513 |
| Total brain change rate Pt 0.05 | -0.0095 | 0.4748 | 0.3840 | -0.0047 | 0.6705 | 0.4983 |
| Total brain change rate Pt 0.1 | -0.0162 | 0.2493 | 0.3771 | -0.0021 | 0.8624 | 0.3944 |
| Total brain change rate Pt 0.5 | -0.0158 | 0.2325 | 0.5937 | -0.0042 | 0.7046 | 0.7324 |
| Total brain change rate Pt 1 | -0.0153 | 0.2438 | 0.6518 | -0.0029 | 0.7901 | 0.7154 |
| Nucleus accumbens change rate Pt 0.0001 | 0.0165 | 0.5270 | **0.0078** | -0.0112 | 0.3418 | 0.5054 |
| Nucleus accumbens change rate Pt 0.01 | -0.0096 | 0.5243 | 0.2625 | -0.0066 | 0.6649 | 0.1574 |
| Nucleus accumbens change rate Pt 0.05 | -0.0030 | 0.8016 | 0.7304 | 0.0011 | 0.9466 | 0.0795 |
| Nucleus accumbens change rate Pt 0.1 | -0.0039 | 0.7437 | 0.7204 | -0.0036 | 0.8314 | 0.0685 |
| Nucleus accumbens change rate Pt 0.5 | -0.0009 | 0.9410 | 0.4793 | -0.0073 | 0.6368 | 0.1319 |
| Nucleus accumbens change rate Pt 1 | 0.0010 | 0.9348 | 0.5518 | -0.0068 | 0.6569 | 0.1461 |

Supplementary table 23. Full meta-analysis results. Bold indicates a p value below 0.05. Polygenic indices nominally associated with mental health symptoms in men are ADHD, BD, cross-disorder, ED, MDD, PTSD, SCZ, and TBV, albeit with some heterogeneity for the latter. Polygenic indices nominally associated with mental health symptoms in women are ADHD, cross-disorder, MDD and cerebellum white matter change rate. AD = anxiety disorders, ADHD = attention deficit hyperactivity disorder, ASD = autism spectrum disorder, BD = bipolar disorder, ED = eating disorder, FA = fractional anisotropy, FX = fornix, FXST = fornix/stria terminalis, GCC = genu of corpus callosum, GM = grey matter, ICA = independent components analysis, LPostCing = left posterior cingulate, LRostMidFront = left rostral middle frontal, MD = mean diffusivity, MDD = major depressive disorder, MO = mode of anisotropy, OCD = obsessive-compulsive disorder, PostThalNuc = posterior thalamic nucleus, Pt = p value threshold, PTSD = post-traumatic stress disorder, RD = radial diffusivity, RLatVent = right lateral ventricle, RPrec = right precuneus, SCC = splenium of corpus callosum, SCZ = schizophrenia, SUD = substance use disorders, TBV = total brain volume, WM = white matter, WMH = white matter hyperintensities.

| | | GPS | | | | BHPS | | | |
|---|---|---|---|---|---|---|---|---|---|
| | | Men | | Women | | Men | | Women | |
| EPGI | Stratum | β | p | β | p | β | p | β | p |
| LRost Mid Front $P_t$ 1.0 | Greatest | 0.0569 | 0.647 | -0.115 | 0.288 | 0.232 | 0.620 | 0.0819 | 0.632 |
| | Silent | -0.0849 | 0.0147 | -0.0229 | 0.520 | 0.0569 | 0.407 | 0.0732 | 0.223 |
| | Baby boomer | -0.0499 | 0.163 | -0.0148 | 0.612 | 0.147 | 0.0375 | -0.0670 | 0.209 |
| | X | -0.0938 | 0.0198 | -0.0468 | 0.189 | 0.215 | 0.0130 | 0.0299 | 0.667 |
| | Millennial | -0.0652 | 0.456 | 0.0805 | 0.273 | -0.261 | 0.473 | -0.0283 | 0.921 |
| GM $P_t$ 0.01 | Greatest | -0.0442 | 0.757 | 0.119 | 0.257 | -0.502 | 0.418 | -0.0474 | 0.829 |
| | Silent | -0.0520 | 0.140 | -0.0636 | 0.0794 | 0.0903 | 0.156 | -0.0665 | 0.253 |
| | Baby boomer | -0.0626 | 0.0675 | -0.0705 | 0.0163 | -0.0253 | 0.687 | -0.0480 | 0.398 |
| | X | -0.0287 | 0.476 | -0.0582 | 0.0927 | 0.159 | 0.0669 | 0.0844 | 0.237 |
| | Millennial | -0.0615 | 0.478 | -0.119 | 0.107 | -0.494 | 0.274 | 0.0304 | 0.943 |
| TBV $P_t$ 0.01 | Greatest | 0.0520 | 0.686 | -0.131 | 0.254 | 1.66 | 0.0185 | 0.189 | 0.176 |
| | Silent | -0.0312 | 0.387 | -0.0686 | 0.0552 | 0.00403 | 0.945 | -0.0078 | 0.898 |
| | Baby boomer | -0.0862 | 0.0103 | -0.0631 | 0.0265 | 0.00533 | 0.935 | -0.0065 | 0.905 |
| | X | 0.0218 | 0.605 | -0.0555 | 0.129 | -0.0416 | 0.624 | 0.0490 | 0.478 |
| | Millennial | 0.0592 | 0.475 | -0.163 | 0.0407 | 0.468 | 0.105 | 0.129 | 0.523 |

Supplementary table 24. Polygenic index (PGI) associations with psychological distress in age- and sex-stratified sub-samples of Understanding Society (UKHLS). Coefficients for PGI terms in linear regressions of psychological distress adjusted for family intact, father employed, mother educated, age and $age^2$ (covariates individually removed if they lack variation in small sub-samples). BHPS = British Household Panel Survey, EPGI = endophenotype polygenic index, GM = total grey matter volume, GPS = General Population Sample, LRostMidFront = left rostral middle frontal region volume, $P_t$ = p value threshold, TBV = total brain volume.

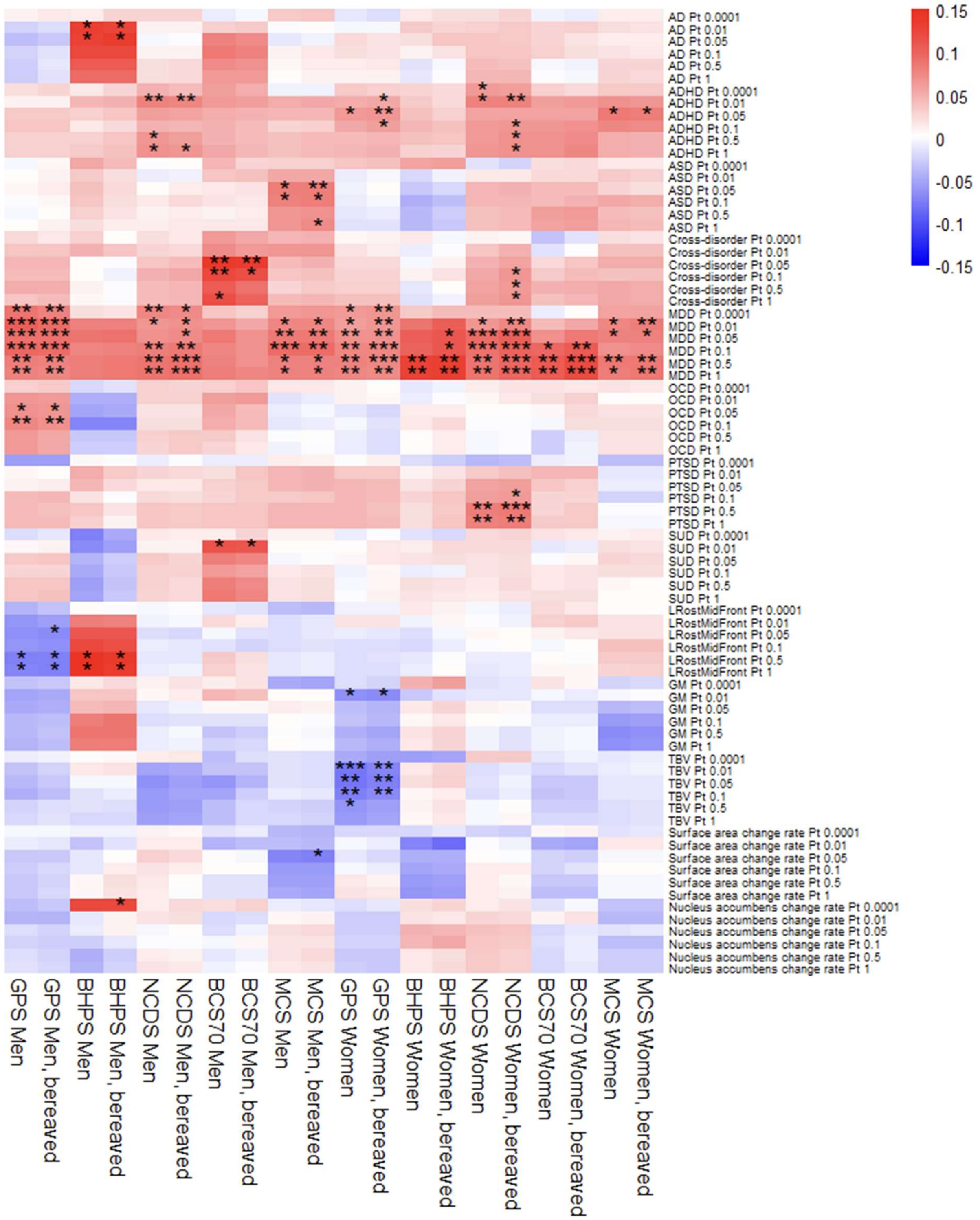


Supplementary figure 5. Associations between polygenic indices (PGIs) and psychological distress in sensitivity analyses which include participants whose parent(s) passed away during youth. Main analysis results are also presented. Cross-disorder PGI becomes associated in NCDS women, surface area change rate PGI becomes associated in MCS men, and nucleus accumbens change rate PGI becomes associated in BHPS men. * = p < 0.05/41, ** = p < 0.01/41, *** = p < 0.001/41. AD = anxiety disorders, ADHD = attention deficit hyperactivity disorder, ASD = autism spectrum disorder, BCS70 = the 1970 British Cohort Study, BHPS = British Household Panel Survey, GM = total grey matter volume, GPS = general population sample of Understanding Society, LRostMidFront = left rostral middle frontal region volume, MCS = Millennium Cohort Study, MDD = major depressive disorder, NCDS = the 1958 National Child Development Study, OCD = obsessive-compulsive disorder, Pt = p value threshold, PTSD = post-traumatic stress disorder, SUD = substance use disorders, TBV = total brain volume.

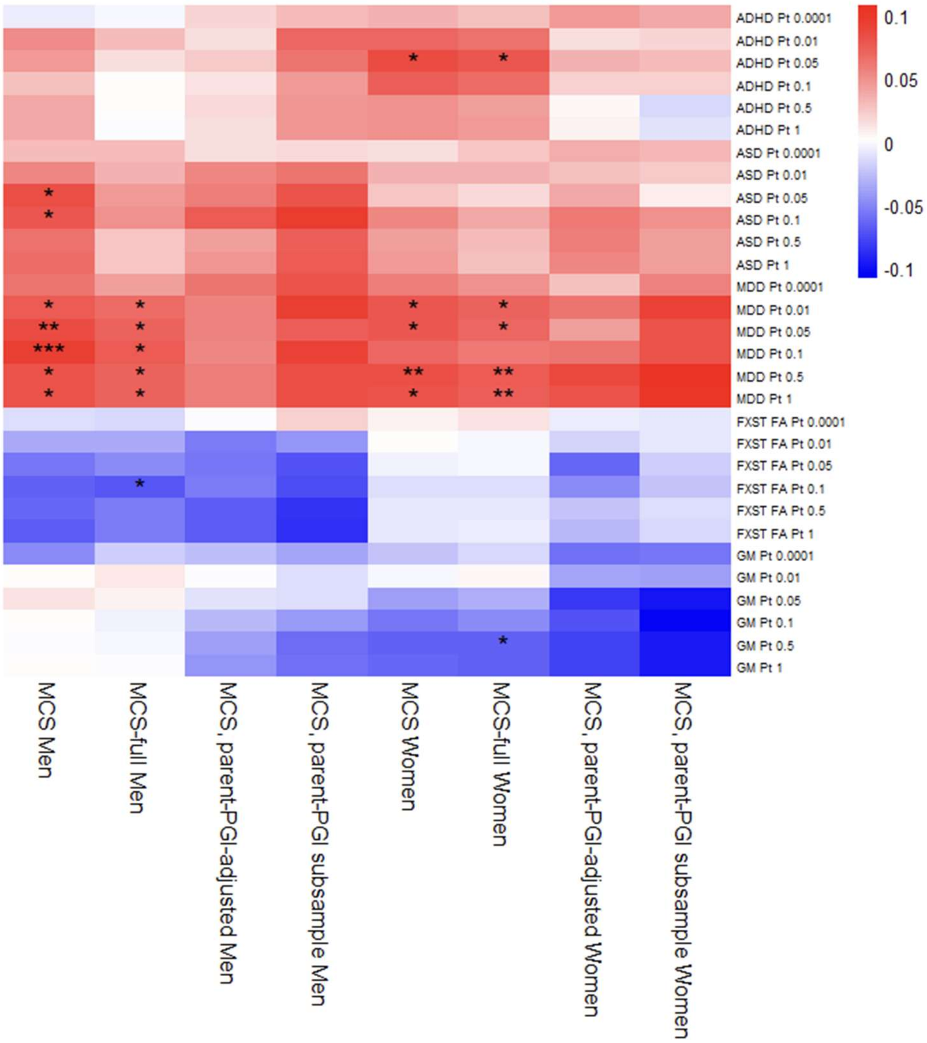


Supplementary figure 6. Associations between polygenic indices (PGIs) and psychological distress in sensitivity analyses specific to MCS. Main analysis results are also presented. The association with GM PGI in GPS women (main analysis) was replicated in MCS-full women. The association with ASD PGI in MCS men was lost in MCS-full men, but an association was gained with FXST FA PGI in this sample. * = p < 0.05/41, ** = p < 0.01/41, *** = p < 0.001/41. ADHD = attention deficit hyperactivity disorder, ASD = autism spectrum disorder, FA = fractional anisotropy, FXST = fornix/stria terminalis, GM = total grey matter volume, MCS = Millennium Cohort Study, MDD = major depressive disorder, PGI = polygenic index, Pt = p value threshold.

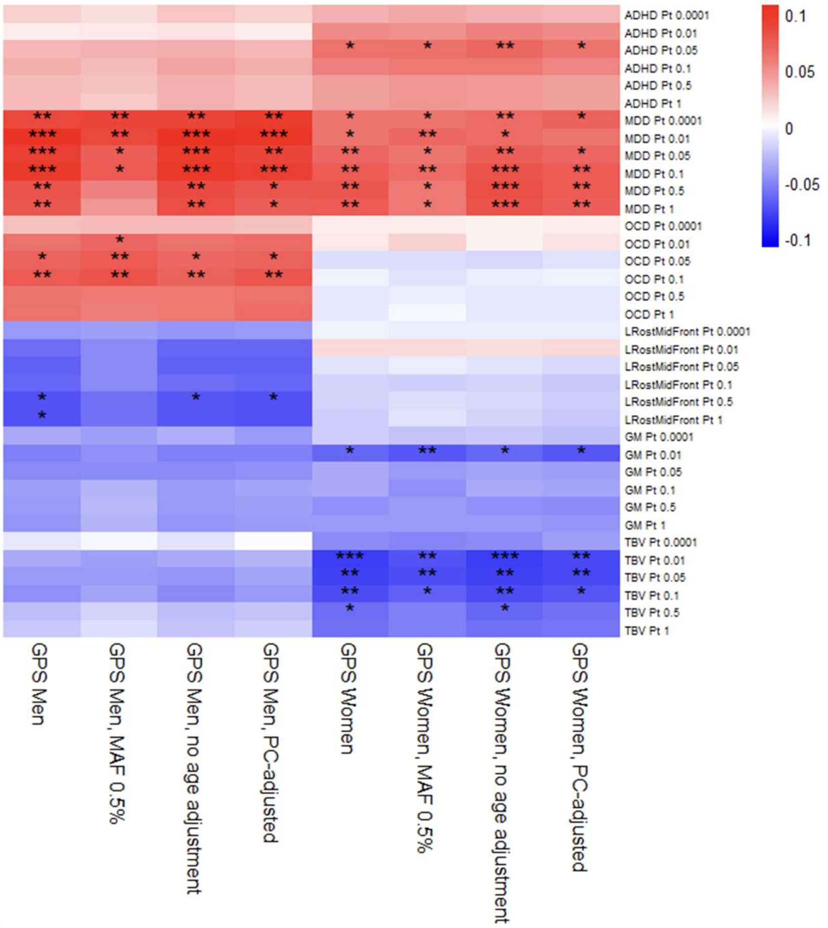


Supplementary figure 7. Associations between polygenic indices (PGIs) and psychological distress in sensitivity analyses specific to GPS. Main analysis results are also presented. Inclusion of rarer alleles removed the association for LRostMidFront in men. Adjusting for PCs, and not adjusting for age, make little difference. * = p < 0.05/41, ** = p < 0.01/41, *** = p < 0.001/41. ADHD = attention deficit hyperactivity disorder, GM = total grey matter volume, GPS = general population sample of Understanding Society, LRostMidFront = left rostral middle frontal region volume, MAF = minor allele frequency (filtering), MDD = major depressive disorder, OCD = obsessive-compulsive disorder, PC = principal components (of ancestry), Pt = p value threshold, TBV = total brain volume.

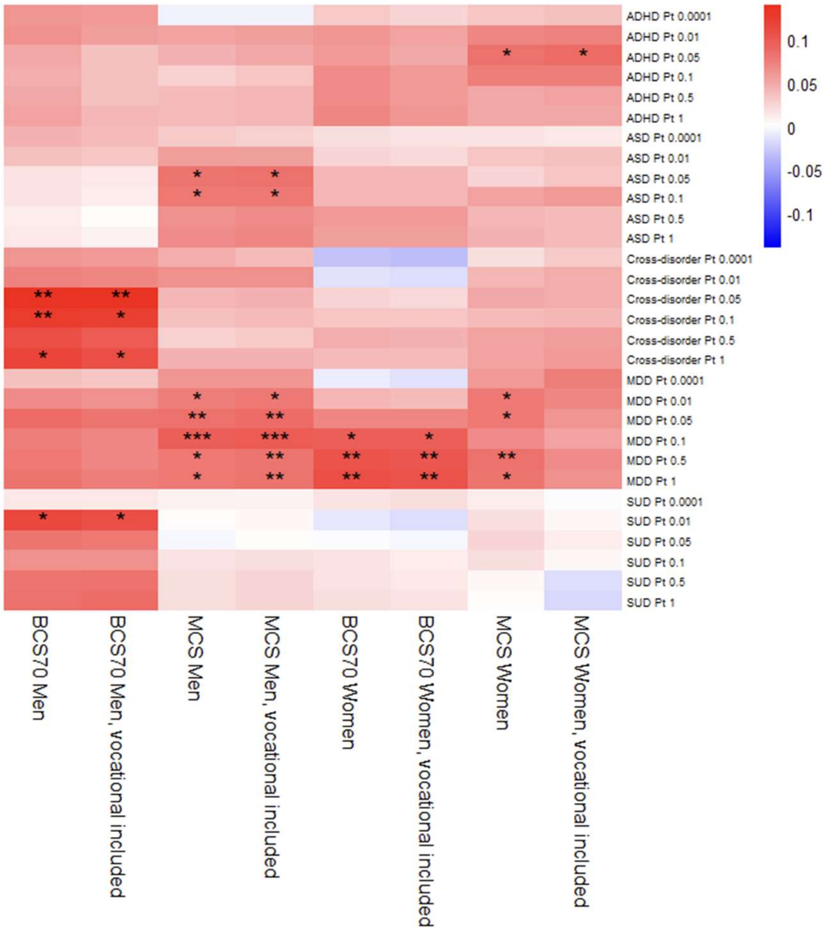


Supplementary figure 8. Associations between polygenic indices (PGIs) and psychological distress in sensitivity analyses which use an alternative measure of mother's education (including vocational qualifications). Main analysis results are also presented. The association between MDD PGI and mental health symptoms is lost in MCS women. * = p < 0.05/41, ** = p < 0.01/41, *** = p < 0.001/41. ADHD = attention deficit hyperactivity disorder, ASD = autism spectrum disorder, BCS70 = the 1970 British Cohort Study, MCS = Millennium Cohort Study, MDD = major depressive disorder, Pt = p value threshold, SUD = substance use disorders.

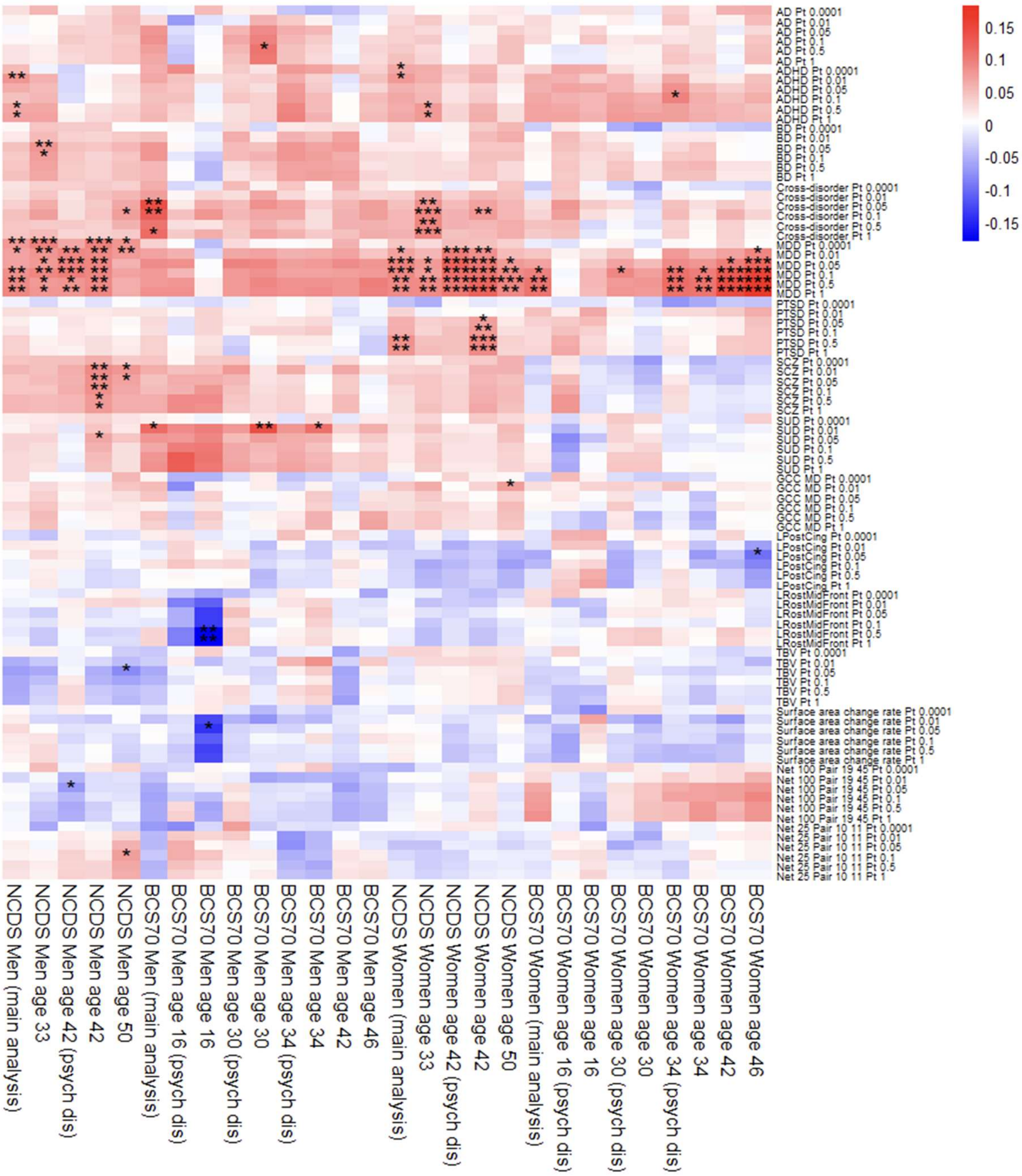


Supplementary figure 9. Associations between polygenic indices (PGIs) and psychological distress in sensitivity analyses using symptom measures at different ages and using different scales. Main analysis results are also presented. MDD PGI associations were generally robust across ages, but ADHD PGI associations in NCDS did not persist for men, and only persisted until the next wave for women. For men, SCZ PGI became associated at ages 42 and 50 in NCDS, and the SUD PGI-malaise association in BCS70 persisted until age 34. Less robust associations appeared in NCDS men for two neuropsychiatric PGIs (BD and cross-disorder), and three endophenotype PGIs (TBV, Net 100 Pair 19 45 and Net25 Pair 10 11), and in BCS70 men for LRostMidFront PGI, surface area change rate PGI, and AD PGI. For women, cross-disorder PGI became associated with malaise at ages 33 and 42 in NCDS, and the PTSD PGI-malaise association in NCDS was repeated at age 42. Less robust associations appeared in NCDS women for GCC MD PGI, and in BCS70 women for LPostCing PGI. In BCS70 men, there were no PGI associations with psychological distress, or with malaise at ages 42 and 46. * = p < 0.05/41, ** = p < 0.01/41, *** = p < 0.001/41. AD = anxiety disorders, ADHD = attention deficit hyperactivity disorder, BCS70 = the 1970 British Cohort Study, BD = bipolar disorder, GCC = genu of corpus callosum, LPostCing = left posterior cingulate volume, LRostMidFront = left rostral middle frontal region volume, MD = mean diffusivity, MDD = major depressive disorder, NCDS = the 1958 National Child Development Study, psych dis = psychological distress, Pt = p value threshold, PTSD = post-traumatic stress disorder, SCZ = schizophrenia, SUD = substance use disorders, TBV = total brain volume.

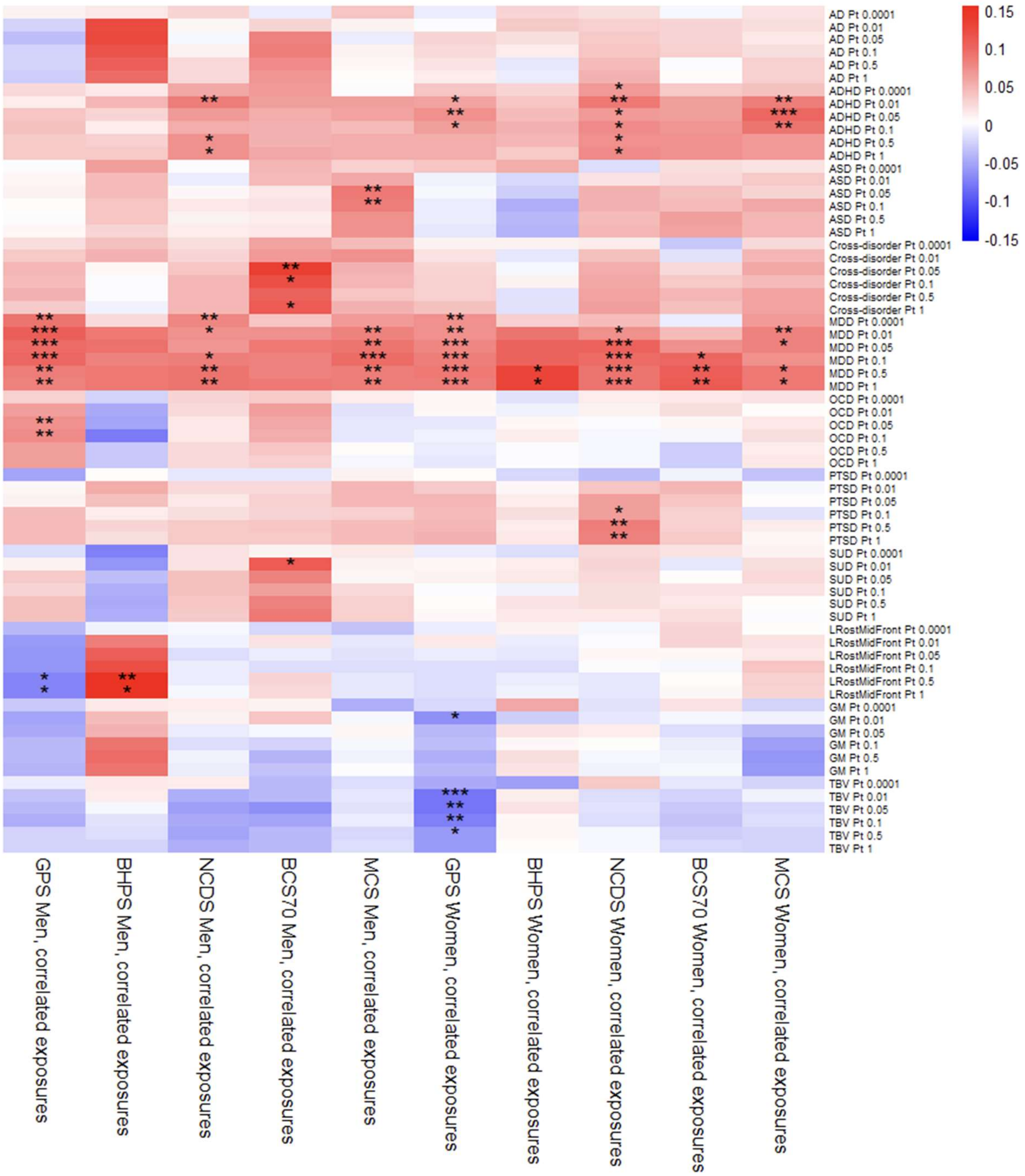


Supplementary figure 10. Associations between polygenic indices (PGIs) and psychological distress in sensitivity analyses wherein pairwise correlation terms between exposures were modelled. The association with AD PGI is lost in BHPS men. * = p < 0.05/41, ** = p < 0.01/41, *** = p < 0.001/41. AD = anxiety disorders, ADHD = attention deficit hyperactivity disorder, ASD = autism spectrum disorder, BCS70 = the 1970 British Cohort Study, BHPS = British Household Panel Survey, GM = total grey matter volume, GPS = general population sample of Understanding Society, LRostMidFront = left rostral middle frontal region volume, MCS = Millennium Cohort Study, MDD = major depressive disorder, NCDS = the 1958 National Child Development Study, OCD = obsessive-compulsive disorder, Pt = p value threshold, PTSD = post-traumatic stress disorder, SUD = substance use disorders, TBV = total brain volume.

| | Men | | | | | Women | | | | |
|---|---|---|---|---|---|---|---|---|---|---|
| PGI | GPS | BHPS | NCDS | BCS70 | MCS | GPS | BHPS | NCDS | BCS70 | MCS |
| AD 0.0001 | | | | | | | | | | |
| AD 0.01 | | **-1.068** | | | | | | | | |
| AD 0.05 | | **4.857** | | 9.399 | | | | 18.884 | | |
| AD 0.1 | | -6.492 | | 12.442 | | | | 18.742 | | |
| AD 0.5 | | -14.539 | | 5.397 | | | | 19.963 | | |
| AD 1.0 | | -17.120 | | 6.100 | | | | 18.732 | | |
| ADHD 0.0001 | | | 20.351 | 27.373 | | 21.423 | | **19.794** | | 30.384 |
| ADHD 0.01 | | | **10.726** | 32.224 | 31.431 | 15.111 | | **26.087** | 6.760 | 28.754 |
| ADHD 0.05 | 26.676 | | 12.679 | 38.644 | 36.505 | **12.083** | | 35.006 | 15.667 | **23.427** |
| ADHD 0.1 | 20.916 | | 12.894 | 39.901 | 49.651 | 16.844 | | 31.850 | 14.738 | 25.241 |
| ADHD 0.5 | | | **11.300** | 36.409 | 38.668 | 20.559 | | 32.885 | 19.448 | 34.185 |
| ADHD 1.0 | | | **9.956** | 34.212 | 39.046 | 19.597 | | 29.860 | 18.723 | 34.283 |
| ASD 0.0001 | | | | | | 0.566 | | | | |
| ASD 0.01 | | | | | 12.738 | | | | | |
| ASD 0.05 | | | | | **10.905** | | | 5.171 | | |
| ASD 0.1 | | | | | **12.069** | | | 9.702 | | -1.180 |
| ASD 0.5 | | | | | 12.438 | | | 9.790 | 10.383 | 6.153 |
| ASD 1.0 | | | | | 10.911 | | | 8.697 | 10.628 | 4.902 |
| Cross-disorder 0.0001 | | | | -6.493 | 10.089 | | | | | |
| Cross-disorder 0.01 | | | | -13.200 | 14.434 | | | 16.545 | | 21.002 |
| Cross-disorder 0.05 | -6.967 | | | **-5.951** | 26.639 | | | 8.095 | | 18.517 |
| Cross-disorder 0.1 | -5.465 | | -5.329 | **-9.618** | 30.032 | | | 4.352 | | 26.633 |
| Cross-disorder 0.5 | -10.235 | | 8.528 | -4.334 | | | | 9.475 | | 18.948 |
| Cross-disorder 1.0 | | | 10.261 | **-6.450** | 17.154 | 5.432 | | 0.722 | | 16.765 |
| MDD 0.0001 | **1.644** | | **7.202** | | 14.531 | **17.565** | | 12.531 | | 17.362 |
| MDD 0.01 | **3.286** | 20.614 | **12.025** | 20.710 | **19.905** | **22.884** | 15.979 | **16.531** | | **18.095** |
| MDD 0.05 | **7.616** | 24.324 | 11.700 | 12.720 | **15.538** | **13.869** | 17.292 | **16.951** | 16.378 | **19.776** |
| MDD 0.1 | **6.568** | 24.490 | **7.139** | 14.344 | **13.969** | **12.367** | 17.578 | **11.375** | **10.706** | 24.524 |
| MDD 0.5 | **7.593** | 22.616 | **9.251** | 13.809 | **16.538** | **12.112** | **16.507** | **14.738** | **8.014** | **21.318** |
| MDD 1.0 | **8.235** | 23.803 | **8.558** | 12.588 | **16.880** | **12.194** | **16.877** | **15.101** | **7.959** | **22.339** |
| OCD 0.0001 | | | | | | | | | | |
| OCD 0.01 | -12.096 | | | | | | | | | |
| OCD 0.05 | **-5.382** | | | | | | | | | |
| OCD 0.1 | **-5.788** | | | | | | | | | |
| OCD 0.5 | -8.777 | | | | | | | | | |
| OCD 1.0 | -9.464 | | | | | | | | | |
| PTSD 0.0001 | -4.794 | | | | | | | | | |
| PTSD 0.01 | | | | | 13.098 | | | 23.613 | 16.958 | |
| PTSD 0.05 | | | | | 15.696 | -3.125 | | 11.368 | | |
| PTSD 0.1 | 3.000 | | | | | -5.793 | | 16.700 | | |

| | | | | | | | | | | |
|---|---|---|---|---|---|---|---|---|---|---|
| PTSD 0.5 | -0.941 | | | | 13.658 | 2.336 | | **7.116** | | |
| PTSD 1.0 | -2.156 | | | | 13.624 | 2.541 | | **6.907** | | |
| SUD 0.0001 | | | | | | | | | | |
| SUD 0.01 | | | | **7.362** | | | | | | |
| SUD 0.05 | | | 33.952 | 16.435 | | | | 51.593 | | |
| SUD 0.1 | | | 35.433 | 20.078 | | | | | | |
| SUD 0.5 | 11.855 | | 39.179 | 14.602 | | | | | | |
| SUD 1.0 | 10.760 | | 41.739 | 13.976 | | | | | | |
| LRostMidFront 0.0001 | | | | | | | | | | |
| LRostMidFront 0.01 | -6.714 | 6.460 | | | | | | | | |
| LRostMidFront 0.05 | -8.231 | 1.451 | | | | | | | | |
| LRostMidFront 0.1 | -5.011 | -5.669 | | | | | | | | |
| LRostMidFront 0.5 | **-1.687** | **1.874** | | | | | | | | |
| LRostMidFront 1.0 | **-3.040** | **1.416** | | | | | | | | |
| GM 0.0001 | | | | | 3.539 | | | | | |
| GM 0.01 | -2.338 | | | | | **3.644** | | | | |
| GM 0.05 | -3.797 | | | | | 17.846 | | | | 21.399 |
| GM 0.1 | | 7.261 | | | | 17.366 | | | | 12.449 |
| GM 0.5 | | -0.546 | | | | 0.167 | | | | 0.798 |
| GM 1.0 | -13.760 | -1.970 | | | | -0.027 | | | | -1.173 |
| TBV 0.0001 | | | | | | 15.433 | | | | |
| TBV 0.01 | | | -27.689 | | | **-12.680** | | | | |
| TBV 0.05 | 4.263 | | -33.496 | -8.629 | | **-15.541** | | | | |
| TBV 0.1 | 5.475 | | -32.199 | | | **-11.197** | | | | |
| TBV 0.5 | | | -29.071 | | | **-10.216** | | | | |
| TBV 1.0 | | | -30.336 | | | -11.661 | | | | |

Supplementary table 25. Estimates of “per cent mediated”, i.e. the extent to which polygenic index (PGI) associations with psychological distress are mediated by the rearing environment. Significant associations shown in bold. Values of 0 and below represent no environmental mediation while positive values indicate some environmental mediation. Values were not calculated when both the linear regression (for estimating total genetic effects) and the path model (for estimating direct genetic effects) had p values greater than 0.05. AD = anxiety disorders, ADHD = attention deficit hyperactivity disorder, ASD = autism spectrum disorder, BCS70 = the 1970 British Cohort Study, BHPS = British Household Panel Survey, GM = total grey matter volume, GPS = general population sample of Understanding Society, LRostMidFront = left rostral middle frontal region volume, MCS = Millennium Cohort Study, MDD = major depressive disorder, NCDS = the 1958 National Child Development Study, OCD = obsessive-compulsive disorder, PGI = polygenic index, PTSD = post-traumatic stress disorder, SUD = substance use disorders, TBV = total brain volume.

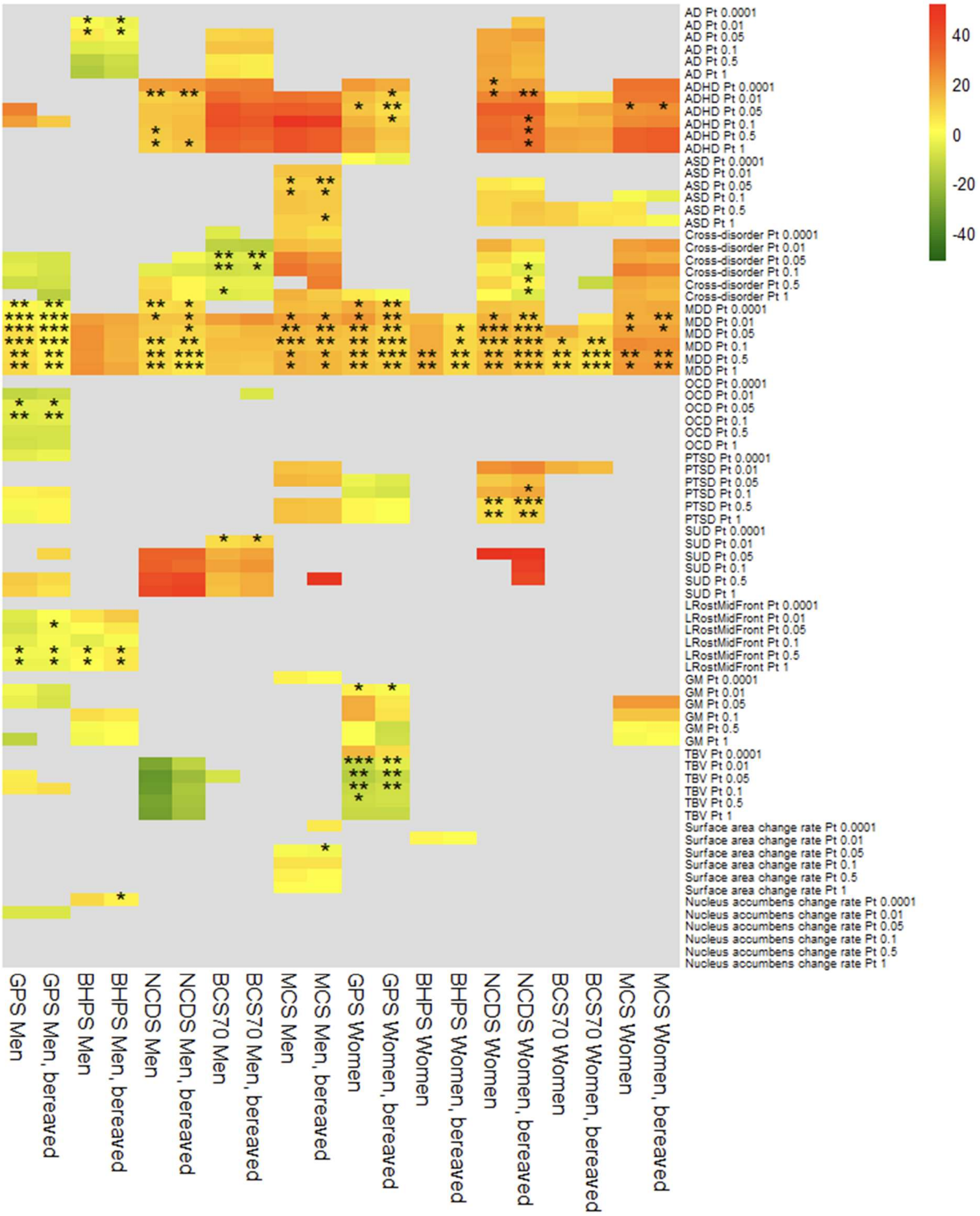


Supplementary figure 11. Estimates of environmental mediation of associations between polygenic indices (PGIs) and psychological distress. Sensitivity analysis results for analyses which include participants whose parent(s) passed away during youth. Main analysis results are also shown. * = p < 0.05/41, ** = p < 0.01/41, *** = p < 0.001/41. AD = anxiety disorders, ADHD = attention deficit hyperactivity disorder, ASD = autism spectrum disorder, BCS70 = the 1970 British Cohort Study, BHPS = British Household Panel Survey, GM = total grey matter volume, GPS = general population sample of Understanding Society, LRostMidFront = left rostral middle frontal region volume, MCS = Millennium Cohort Study, MDD = major depressive disorder, NCDS = the 1958 National Child Development Study, OCD = obsessive-compulsive disorder, Pt = p value threshold, PTSD = post-traumatic stress disorder, SUD = substance use disorders, TBV = total brain volume.

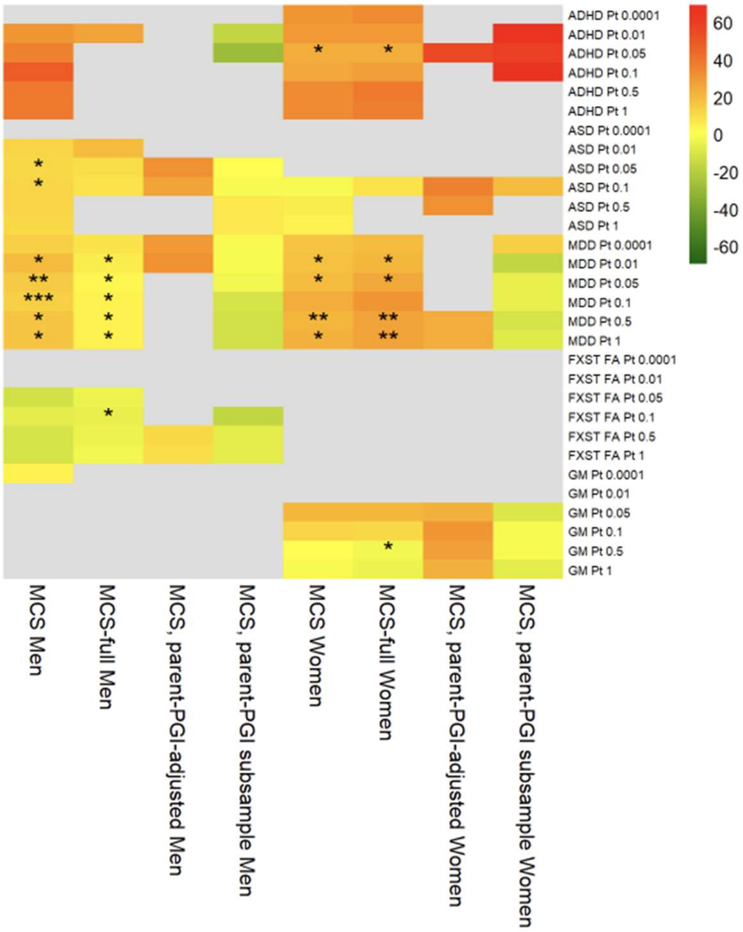


Supplementary figure 12. Estimates of environmental mediation of associations between polygenic indices (PGIs) and psychological distress. Results for MCS-specific sensitivity analyses, and MCS main analysis, are shown. * = $p < 0.05/41$, ** = $p < 0.01/41$, *** = $p < 0.001/41$. ADHD = attention deficit hyperactivity disorder, ASD = autism spectrum disorder, FA = fractional anisotropy, FXST = fornix/stria terminalis, GM = total grey matter volume, MCS = Millennium Cohort Study, MDD = major depressive disorder, PGI = polygenic index, Pt = p value threshold.

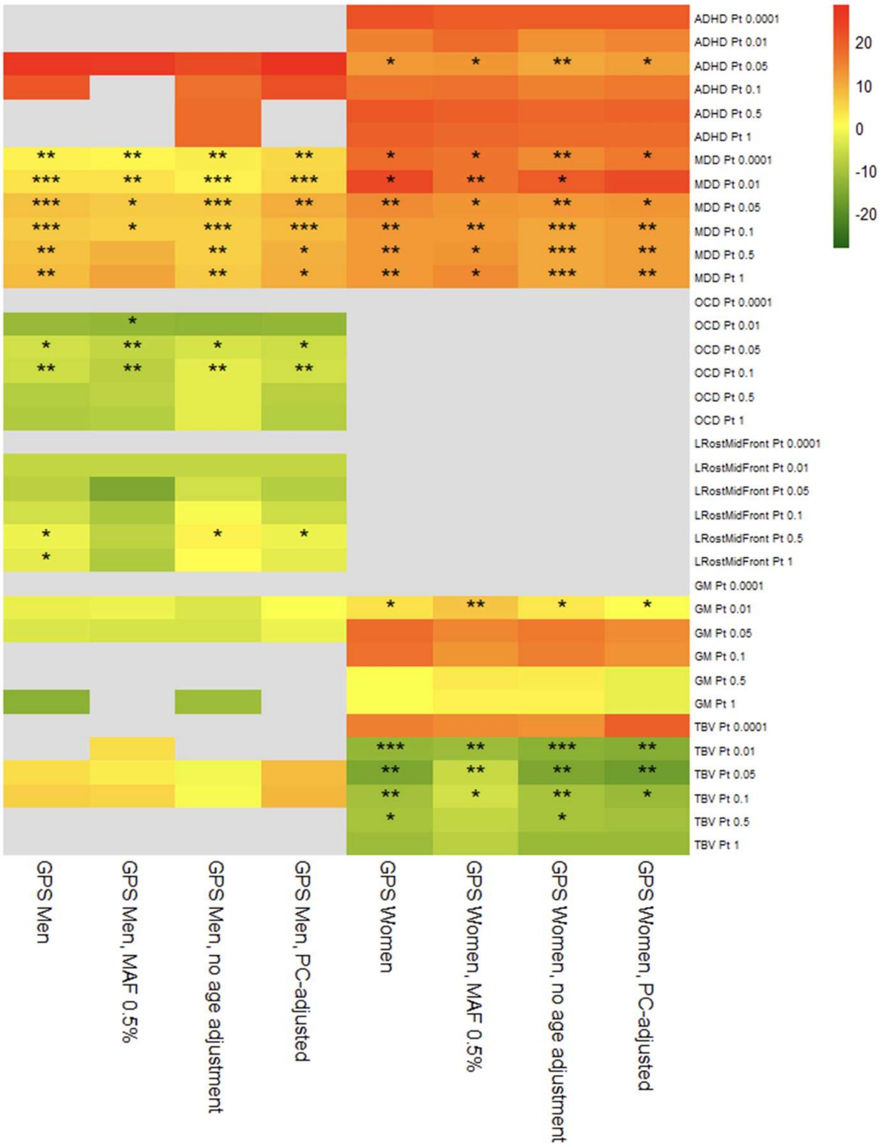


Supplementary figure 13. Estimates of environmental mediation of associations between polygenic indices (PGIs) and psychological distress. Results for GPS-specific sensitivity analyses, and GPS main analysis, are shown. * = p < 0.05/41, ** = p < 0.01/41, *** = p < 0.001/41. ADHD = attention deficit hyperactivity disorder, GM = total grey matter volume, GPS = general population sample of Understanding Society, LRostMidFront = left rostral middle frontal region volume, MAF = minor allele frequency (filtering), MDD = major depressive disorder, OCD = obsessive-compulsive disorder, PC = principal components (of ancestry), Pt = p value threshold, TBV = total brain volume.

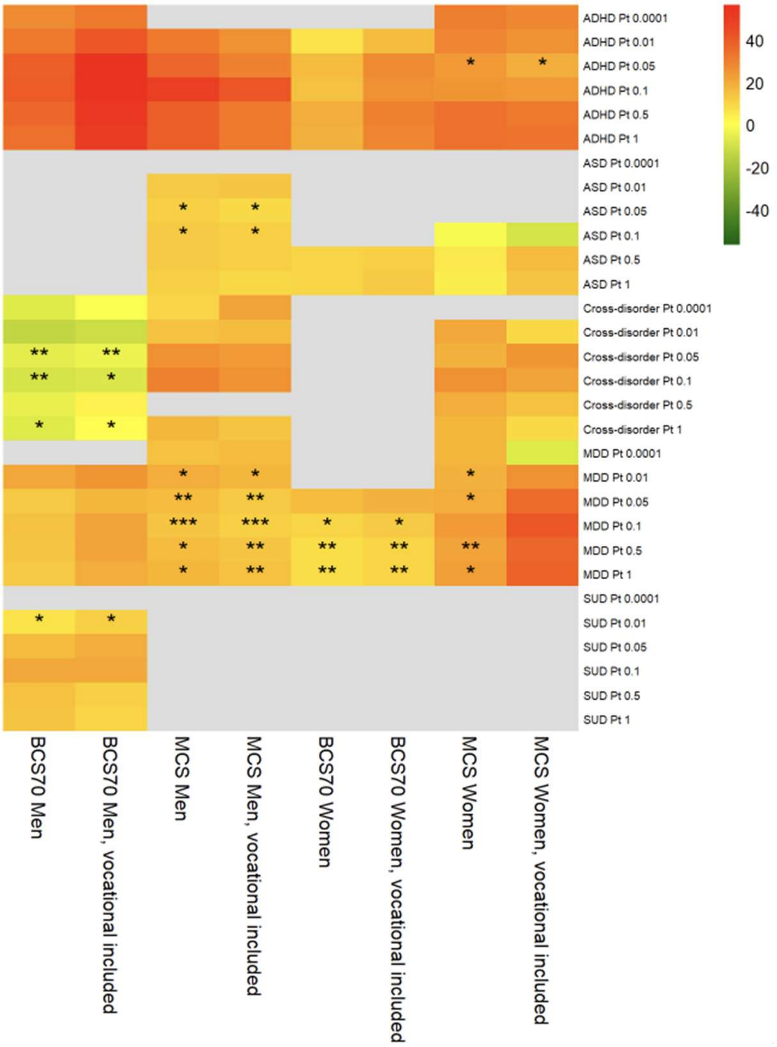


Supplementary figure 14. Estimates of environmental mediation of associations between polygenic indices (PGIs) and psychological distress. Sensitivity analysis results are shown for BCS70 and MCS using an alternative measure of mother's education which includes vocational qualifications. Main analysis results are also shown. * = p < 0.05/41, ** = p < 0.01/41, *** = p < 0.001/41. ADHD = attention deficit hyperactivity disorder, ASD = autism spectrum disorder, BCS70 = the 1970 British Cohort Study, MCS = Millennium Cohort Study, MDD = major depressive disorder, Pt = p value threshold, SUD = substance use disorders.

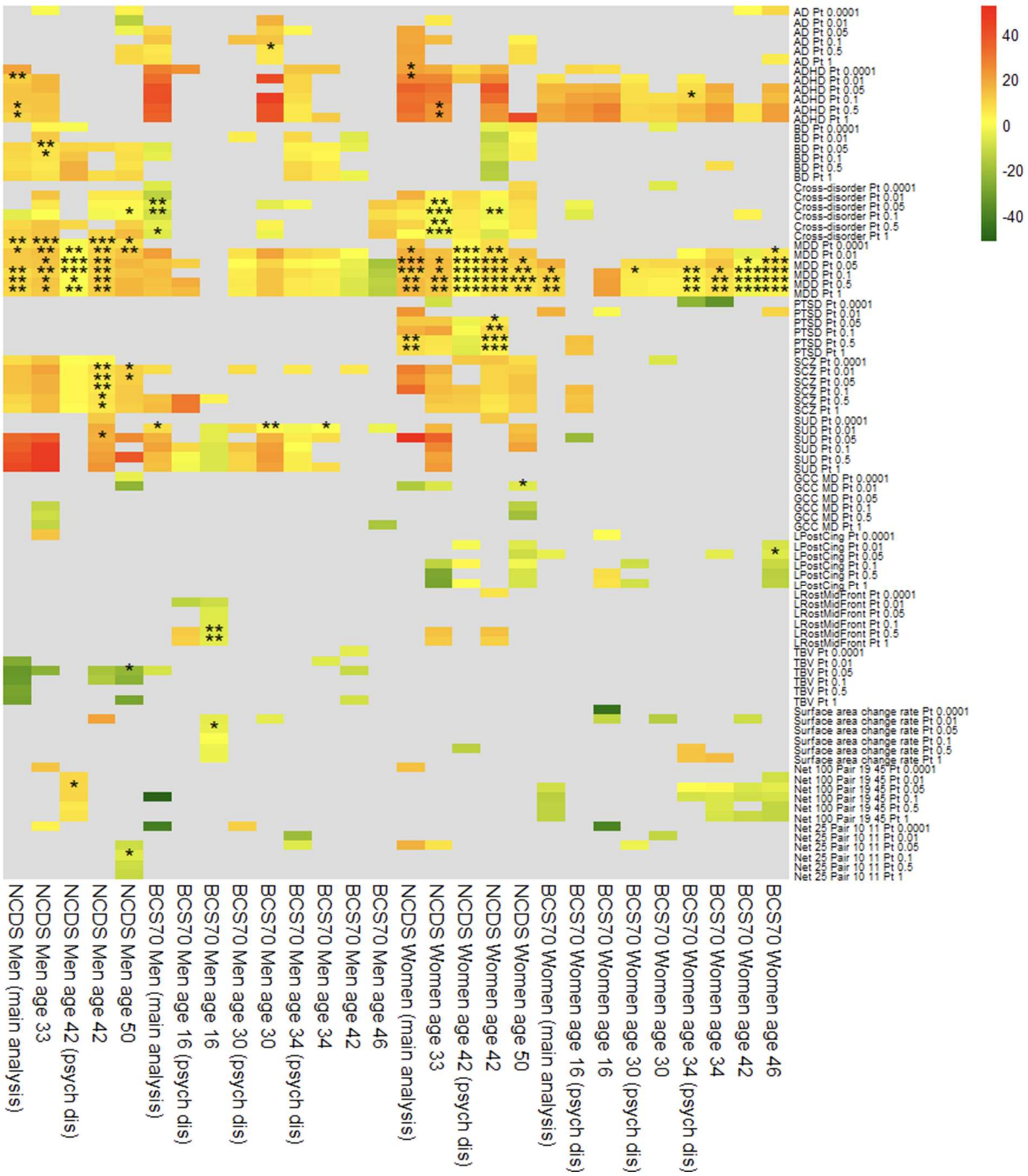


Supplementary figure 15. Estimates of environmental mediation of associations between polygenic indices (PGIs) and psychological distress. Sensitivity analysis results for NCDS and BCS70 using mental health outcomes at different ages. Mediation estimates were mostly attenuated with increasing age - for example, SUD PGI's association with malaise became less mediated in BCS70 men (main analysis age 26: 7.4%, age 30: 2.1%, age 34: 0.0%, $P_t$ 0.01), and MDD PGI's associations generally became less mediated in women (main analysis age 26: 16.4%, age 34: 12.8%, age 46: 6.0%, $P_t$ 0.05). Mediation was generally lower for psychological distress compared to contemporaneous measures of malaise. * = p < 0.05/41, ** = p < 0.01/41, *** = p < 0.001/41. AD = anxiety disorders, ADHD = attention deficit hyperactivity disorder, BCS70 = the 1970 British Cohort Study, BD = bipolar disorder, GCC = genu of corpus callosum, LPostCing = left posterior cingulate, LRostMidFront = left rostral middle frontal, MD = mean diffusivity, MDD = major depressive disorder, NCDS = the 1958 National Child Development Study, psych dis = psychological distress, PTSD = post-traumatic stress disorder, SCZ = schizophrenia, SUD = substance use disorders, TBV = total brain volume.

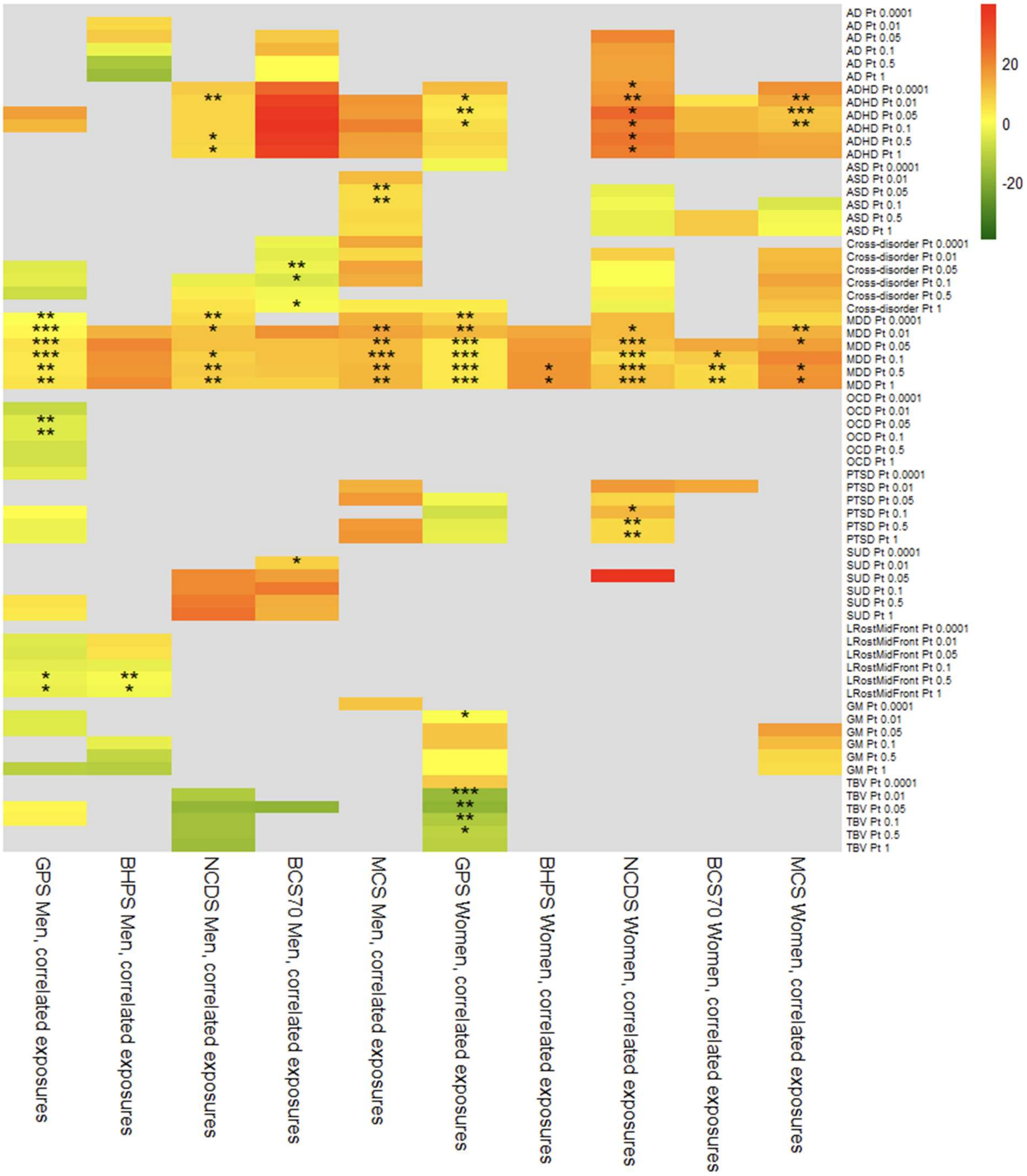


Supplementary figure 16. Estimates of environmental mediation of associations between polygenic indices (PGIs) and psychological distress. Sensitivity analysis results for path models with pairwise correlation terms for the three exposures. Mediation estimates were mostly attenuated by modelling pairwise correlation terms between the exposures. * = p < 0.05/41, ** = p < 0.01/41, *** = p < 0.001/41. AD = anxiety disorders, ADHD = attention deficit hyperactivity disorder, ASD = autism spectrum disorder, BCS70 = the 1970 British Cohort Study, BHPS = British Household Panel Survey, GM = total grey matter volume, GPS = general population sample of Understanding Society, LRostMidFront = left rostral middle frontal, MCS = Millennium Cohort Study, MDD = major depressive disorder, NCDS = the 1958 National Child Development Study, OCD = obsessive-compulsive disorder, PTSD = post-traumatic stress disorder, SUD = substance use disorders, TBV = total brain volume.

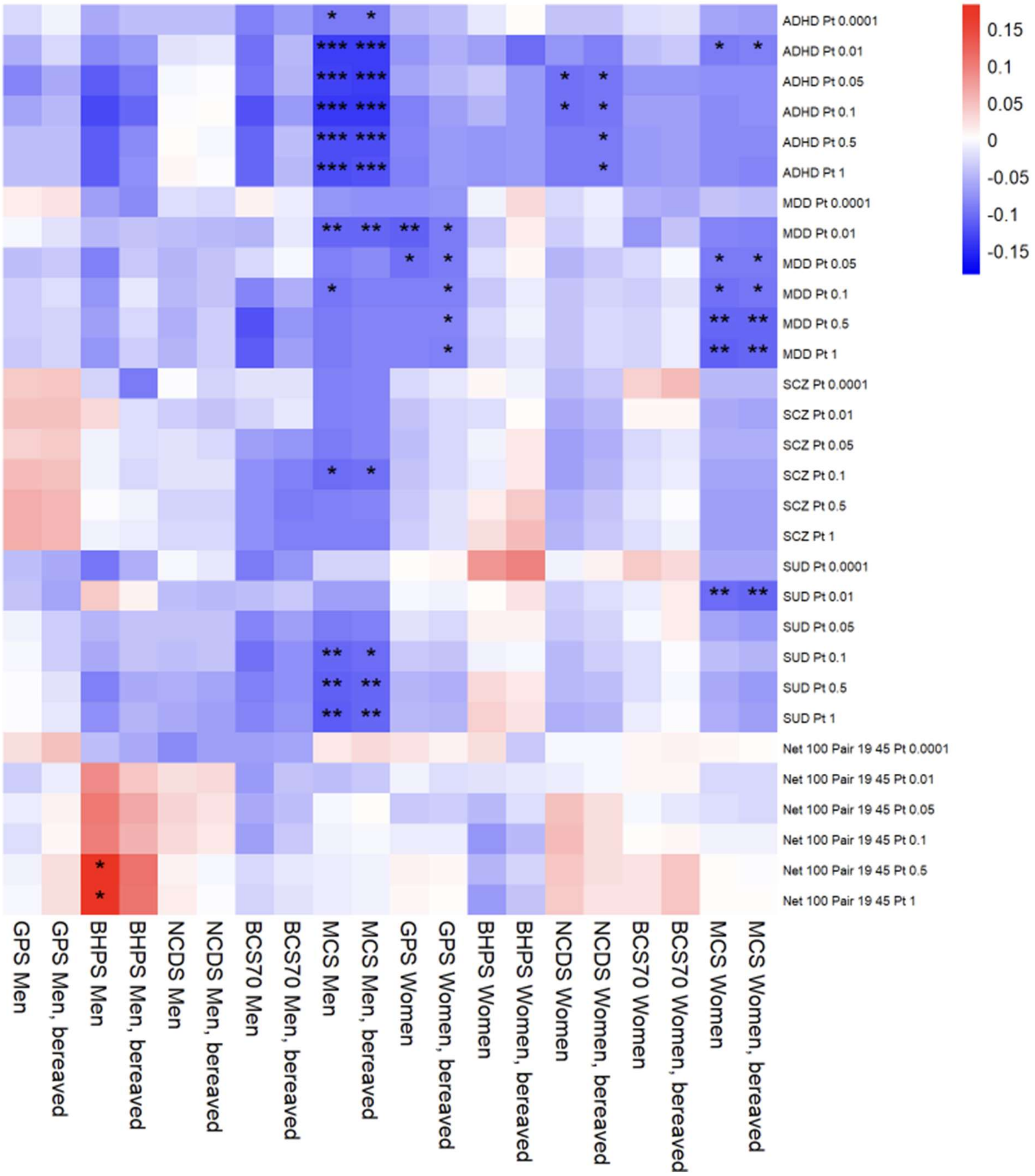


Supplementary figure 17. Associations between PGIs and family intact. Sensitivity analysis results for analyses which include participants whose parent(s) passed away during youth. Main analysis results are also shown. * = p < 0.05/41, ** = p < 0.01/41, *** = p < 0.001/41. ADHD = attention deficit hyperactivity disorder, BCS70 = the 1970 British Cohort Study, BHPS = British Household Panel Survey, GPS = general population sample of Understanding Society, MCS = Millennium Cohort Study, MDD = major depressive disorder, NCDS = the 1958 National Child Development Study, Pt = p value threshold, SCZ = schizophrenia, SUD = substance use disorders.

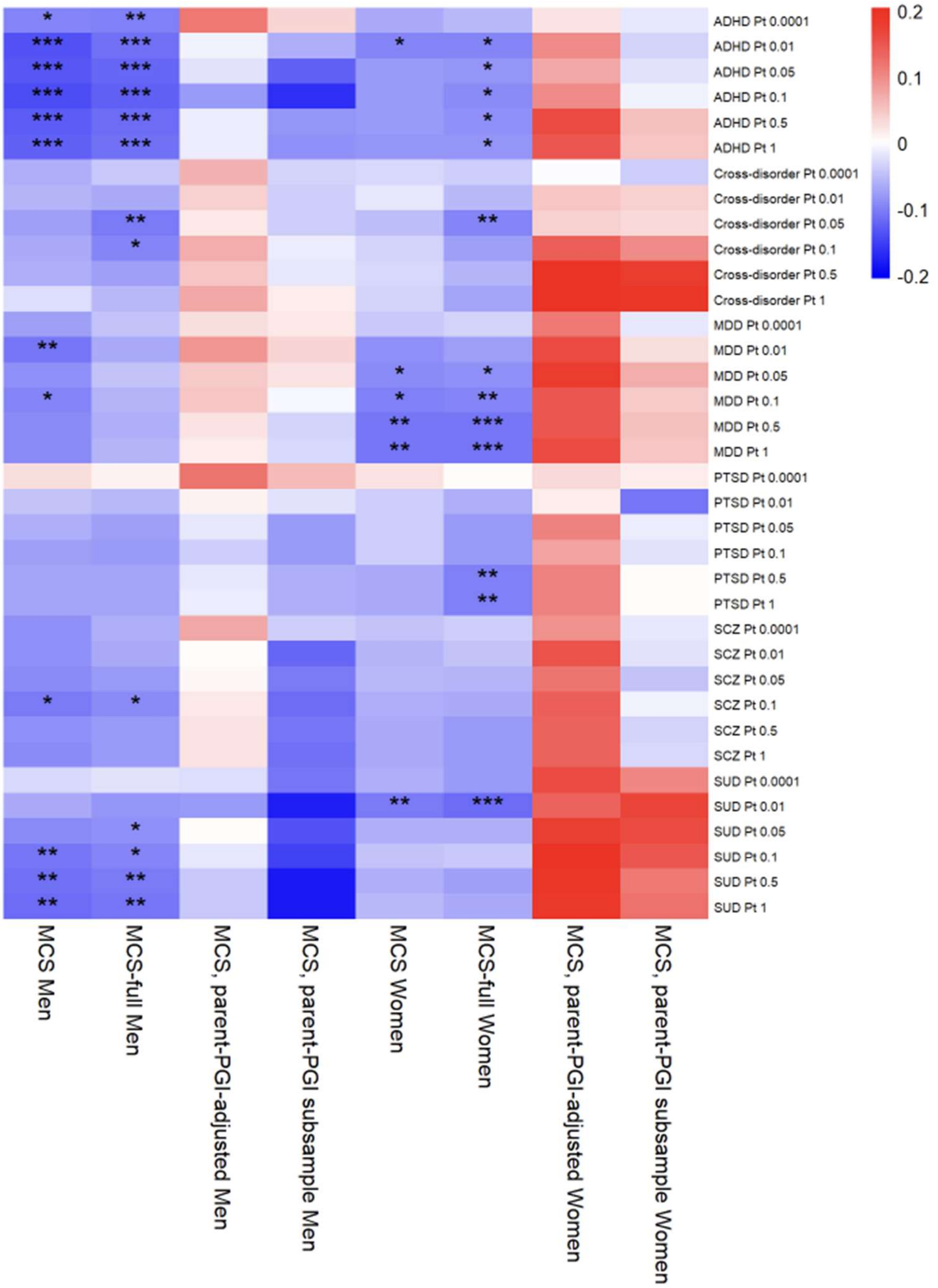


Supplementary figure 18. Associations between polygenic indices (PGIs) and family intact. Results for MCS-specific sensitivity analyses, and MCS main analysis, are shown. Including all ancestries leads to greater gene-environment correlation (rGE) in women, while restricting to participants with parents' PGIs removes all rGE. * = p < 0.05/41, ** = p < 0.01/41, *** = p < 0.001/41. ADHD = attention deficit hyperactivity disorder, MCS = Millennium Cohort Study, MDD = major depressive disorder, PGI = polygenic index, Pt = p value threshold, PTSD = post-traumatic stress disorder, SCZ = schizophrenia, SUD = substance use disorder.

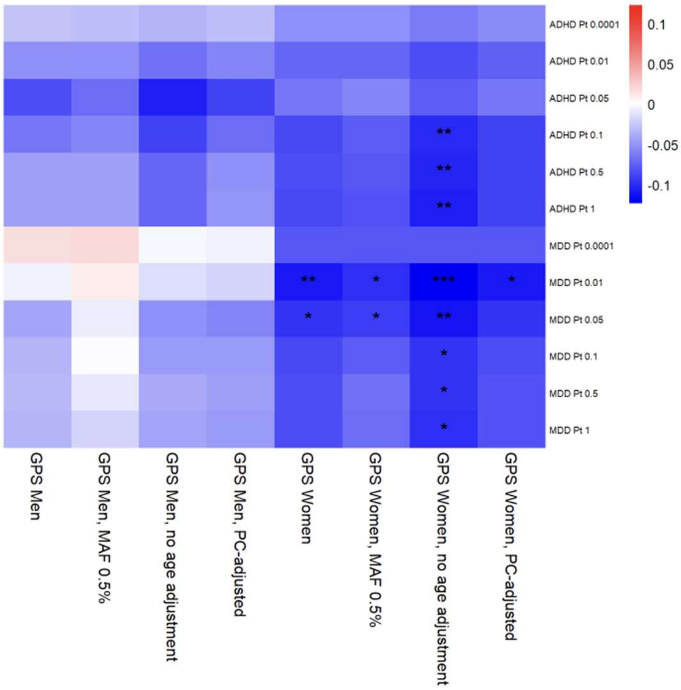


Supplementary figure 19. Associations between polygenic indices (PGIs) and family intact. Results for GPS-specific sensitivity analyses, and GPS main analysis, are shown. * = p < 0.05/41, ** = p < 0.01/41, *** = p < 0.001/41. ADHD = attention deficit hyperactivity disorder, GPS = general population sample of Understanding Society, MAF = minor allele frequency (filtering), MDD = major depressive disorder, PC = principal components (of ancestry), Pt = p value threshold.

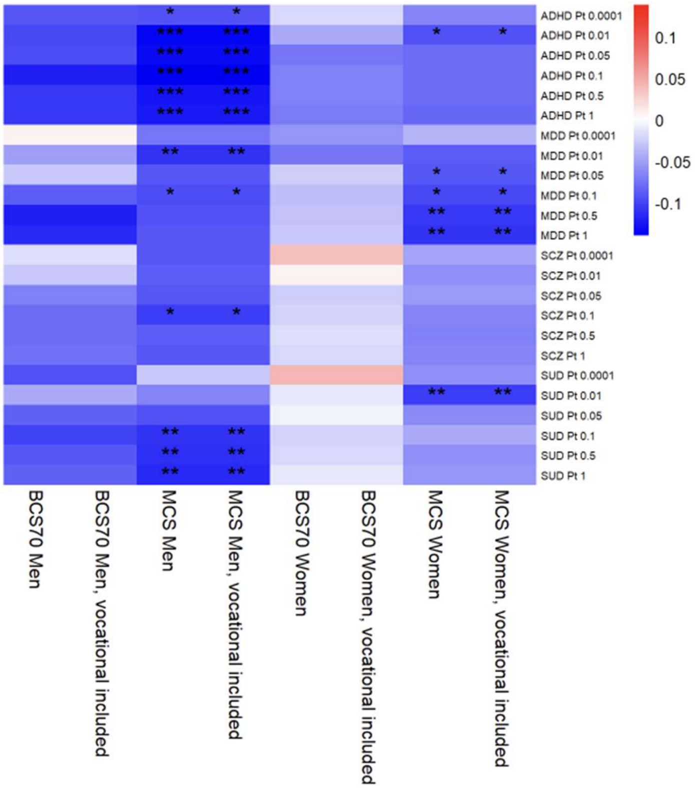


Supplementary figure 20. Associations between polygenic indices (PGIs) and family intact. Sensitivity analysis results are shown for BCS70 and MCS using an alternative measure of mother's education which includes vocational qualifications. Main analysis results are also shown. * = p < 0.05/41, ** = p < 0.01/41, *** = p < 0.001/41. ADHD = attention deficit hyperactivity disorder, BCS70 = the 1970 British Cohort Study, BHPS = British Household Panel Survey, MCS = Millennium Cohort Study, MDD = major depressive disorder, Pt = p value threshold, SCZ = schizophrenia, SUD = substance use disorders.

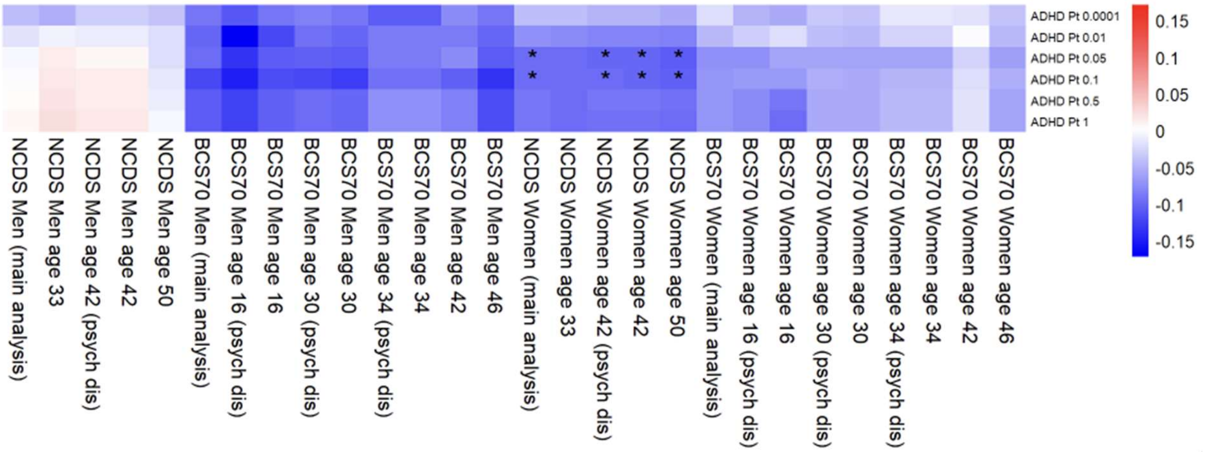


Supplementary figure 21. Associations between polygenic indices (PGIs) and family intact. Sensitivity analysis results for NCDS and BCS70 using mental health outcomes at different ages. * = p < 0.05/41, ** = p < 0.01/41, *** = p < 0.001/41. ADHD = attention deficit hyperactivity disorder, BCS70 = the 1970 British Cohort Study, NCDS = the 1958 National Child Development Study, Pt = p value threshold, psych dis = psychological distress.

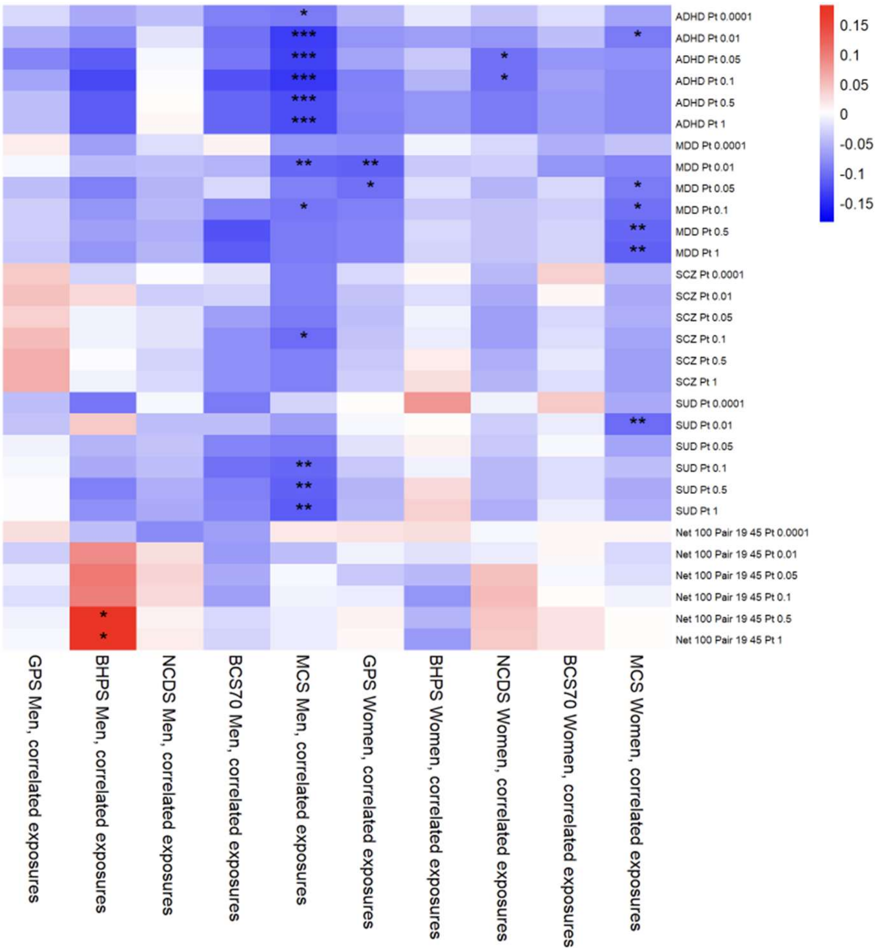


Supplementary figure 22. Associations between polygenic indices (PGIs) and family intact. Sensitivity analysis results for path models with pairwise correlation terms for the three exposures. Results are identical to main analysis. * = p < 0.05/41, ** = p < 0.01/41, *** = p < 0.001/41. ADHD = attention deficit hyperactivity disorder, BCS70 = the 1970 British Cohort Study, BHPS = British Household Panel Survey, GPS = general population sample of Understanding Society, MCS = Millennium Cohort Study, MDD = major depressive disorder, NCDS = the 1958 National Child Development Study, $P_t$ = p value threshold, SCZ = schizophrenia, SUD = substance use disorders.

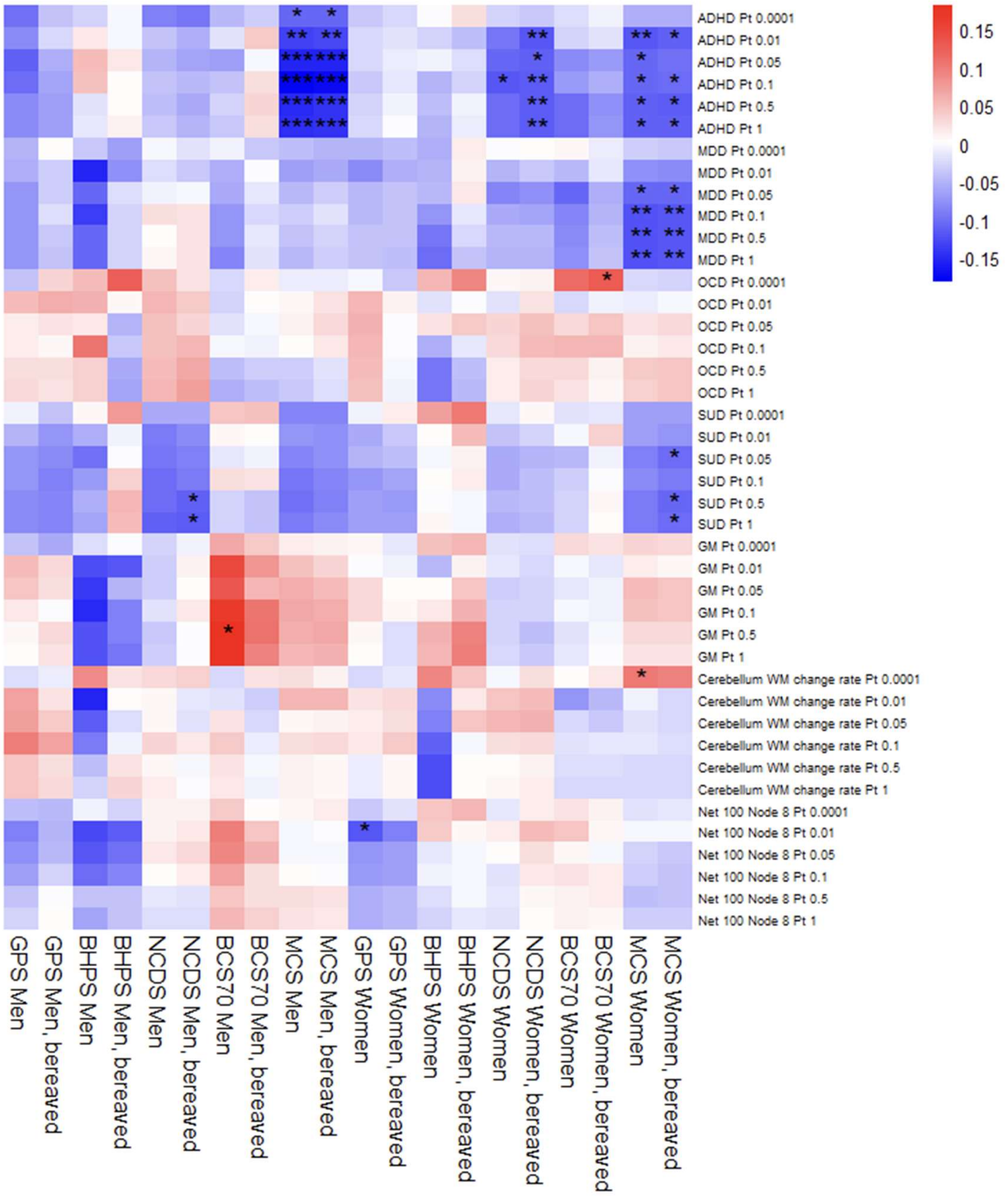


Supplementary figure 23. Associations between polygenic indices (PGIs) and father employed. Sensitivity analysis results for analyses which include participants whose parent(s) passed away during youth. Main analysis results are also shown. SUD PGI becomes associated with father employed in two samples, while non-robust associations are gained or lost for OCD PGI and three endophenotype PGIs. * = p < 0.05/41, ** = p < 0.01/41, *** = p < 0.001/41. ADHD = attention deficit hyperactivity disorder, BCS70 = the 1970 British Cohort Study, BHPS = British Household Panel Survey, GM = total grey matter volume, GPS = general population sample of Understanding Society, MCS = Millennium Cohort Study, MDD = major depressive disorder, NCDS = the 1958 National Child Development Study, OCD = obsessive-compulsive disorder, Pt = p value threshold, SUD = substance use disorders, WM = white matter.

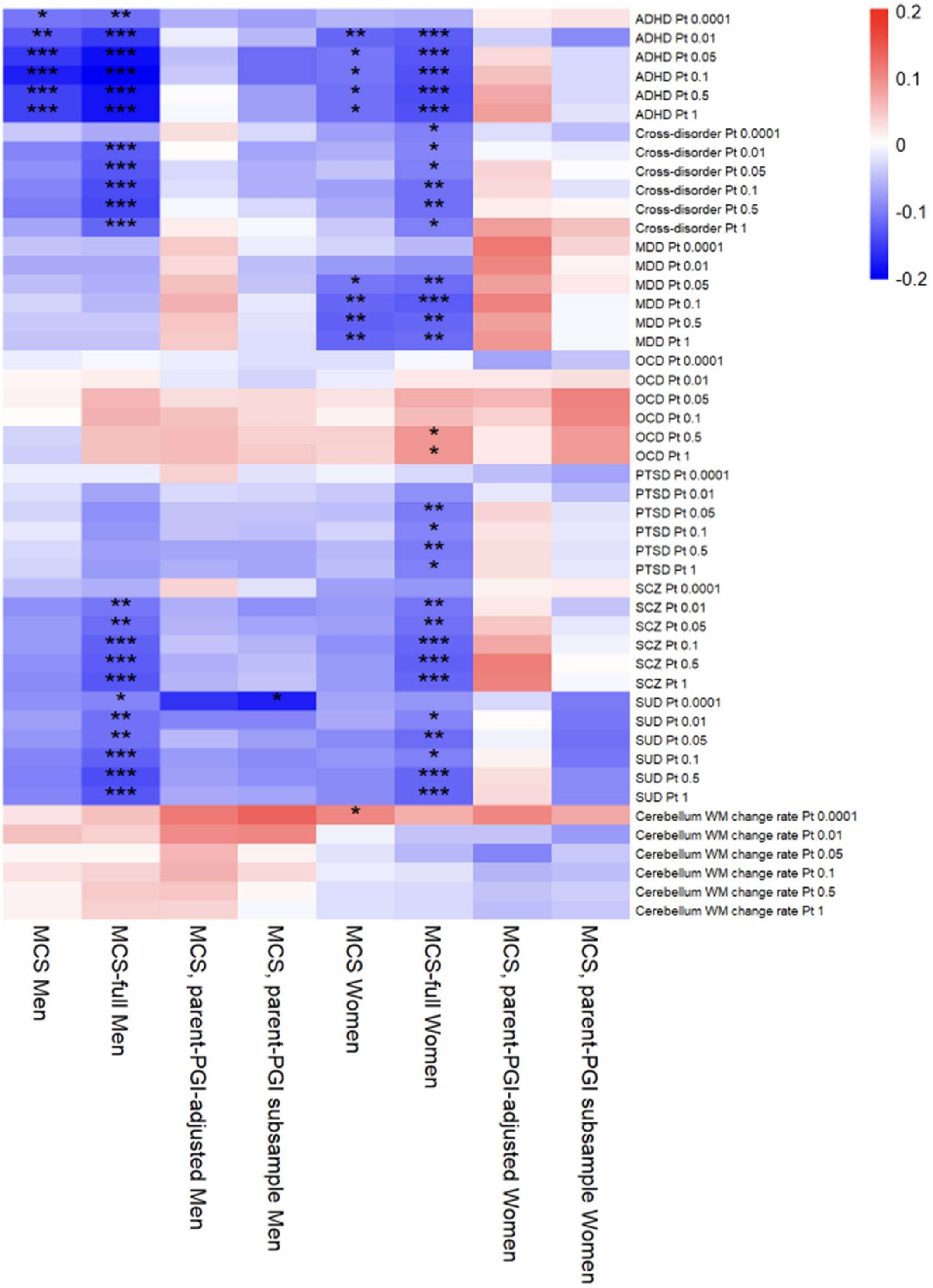


Supplementary figure 24. Associations between polygenic indices (PGIs) and father employed. Results for MCS-specific sensitivity analyses, and MCS main analysis, are shown. Including all ancestries leads to greater gene-environment correlation (rGE), while restricting to participants with parents' PGIs removes most rGE. * = p < 0.05/41, ** = p < 0.01/41, *** = p < 0.001/41. ADHD = attention deficit hyperactivity disorder, MCS = Millennium Cohort Study, MDD = major depressive disorder, OCD = obsessive-compulsive disorder, PTSD = post-traumatic stress disorder, PGI = polygenic index, Pt = p value threshold, SCZ = schizophrenia, SUD = substance use disorder, WM = white matter.

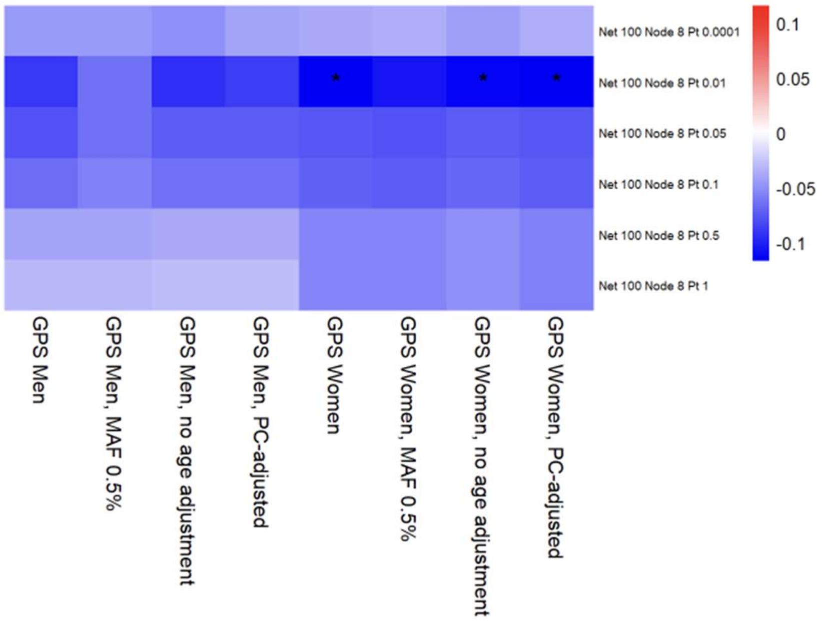


Supplementary figure 25. Associations between polygenic indices (PGIs) and father employed. Results for GPS-specific sensitivity analyses, and GPS main analysis, are shown. * = p < 0.05/41, ** = p < 0.01/41, *** = p < 0.001/41. GPS = general population sample of Understanding Society, MAF = minor allele frequency (filtering), PC = principal components (of ancestry), Pt = p value threshold.

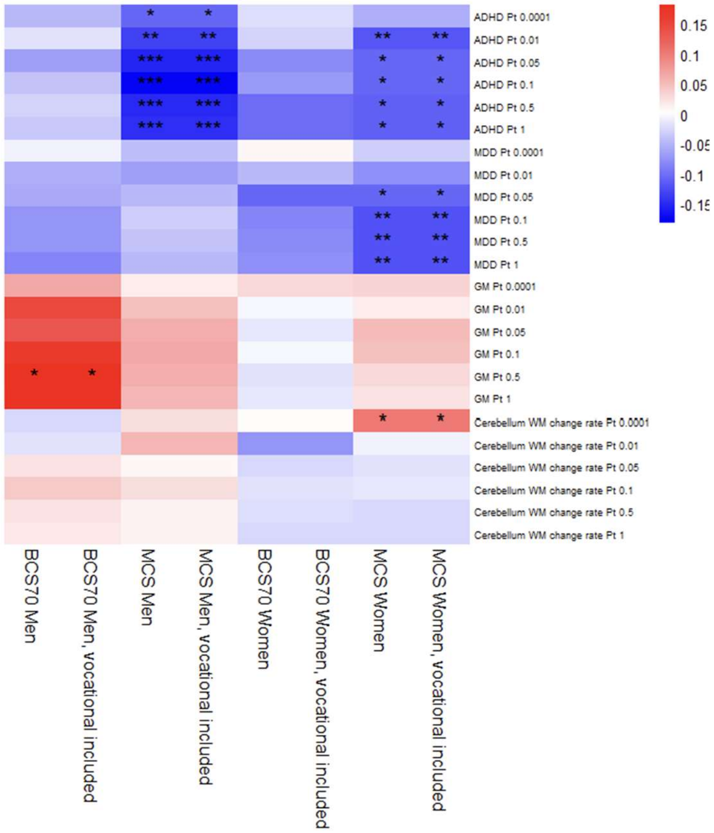


Supplementary figure 26. Associations between polygenic indices (PGIs) and father employed. Sensitivity analysis results are shown for BCS70 and MCS using an alternative measure of mother's education which includes vocational qualifications. Main analysis results are also shown. * = p < 0.05/41, ** = p < 0.01/41, *** = p < 0.001/41. ADHD = attention deficit hyperactivity disorder, BCS70 = the 1970 British Cohort Study, GM = total grey matter volume, MCS = Millennium Cohort Study, MDD = major depressive disorder, Pt = p value threshold, WM = white matter.

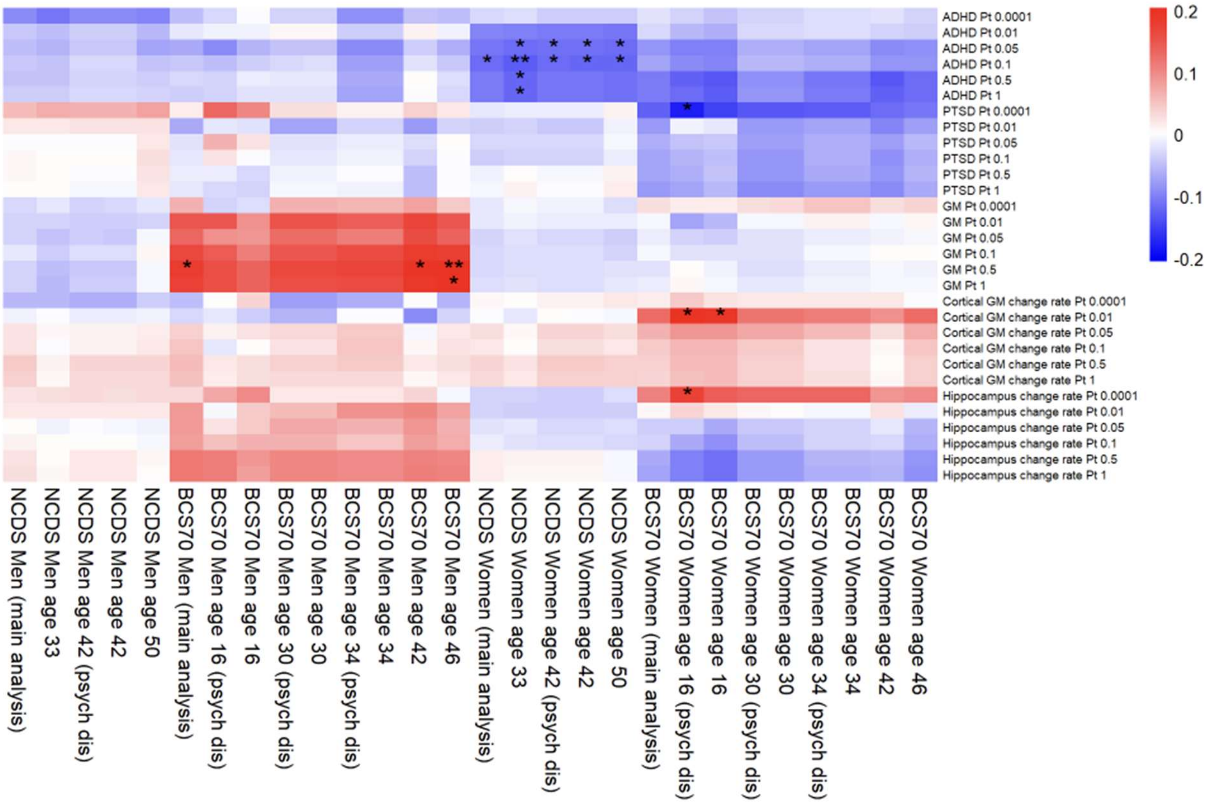


Supplementary figure 27. Associations between polygenic indices (PGIs) and father employed. Sensitivity analysis results for NCDS and BCS70 using mental health outcomes at different ages. * = p < 0.05/41, ** = p < 0.01/41, *** = p < 0.001/41. ADHD = attention deficit hyperactivity disorder, BCS70 = the 1970 British Cohort Study, GM = grey matter volume, NCDS = the 1958 National Child Development Study, psych dis = psychological distress, PTSD = post-traumatic stress disorder.

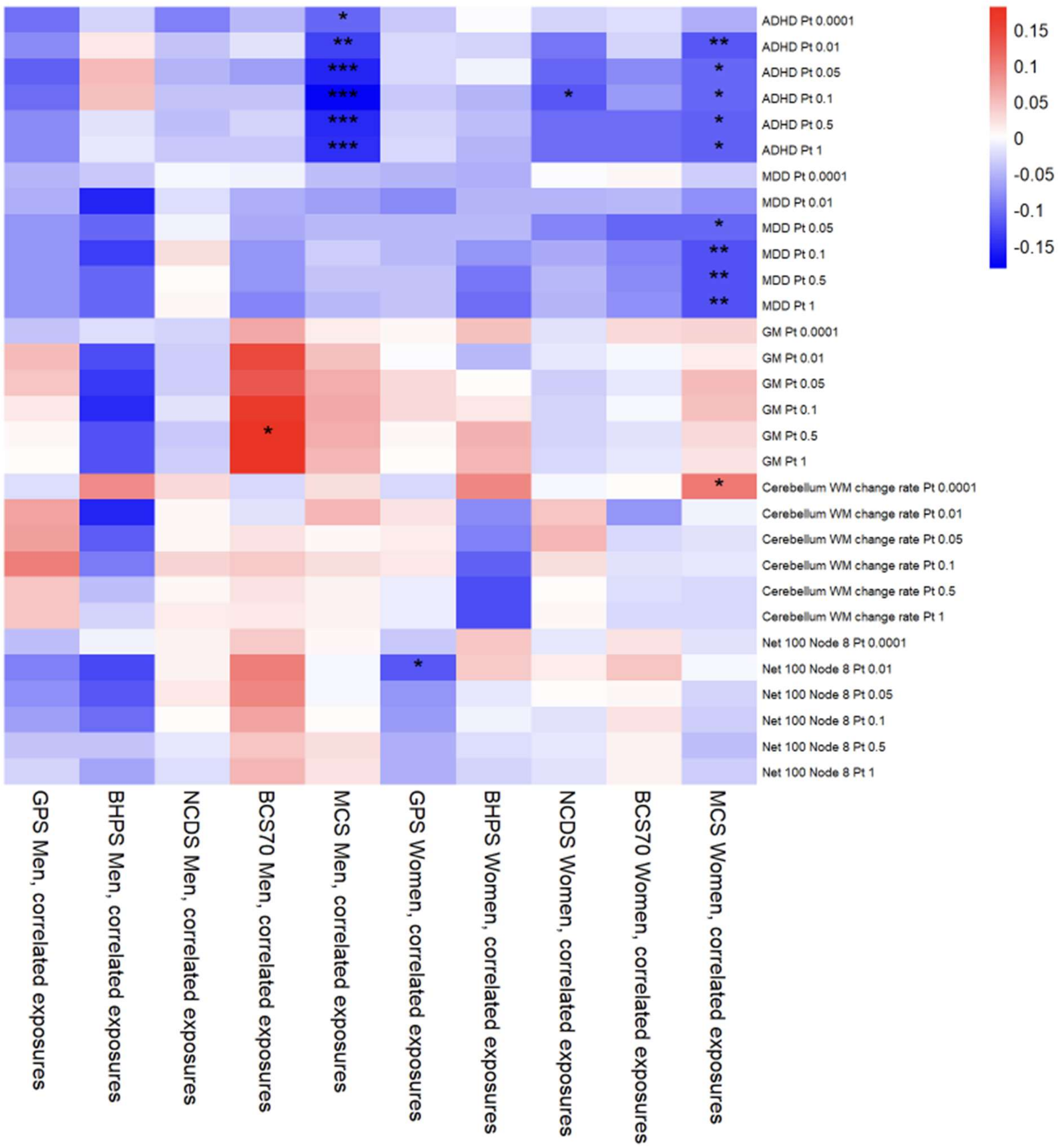


Supplementary figure 28. Associations between polygenic indices (PGIs) and father employed. Sensitivity analysis results for path models with pairwise correlation terms for the three exposures. Results are identical to main analysis. * = p < 0.05/41, ** = p < 0.01/41, *** = p < 0.001/41. ADHD = attention deficit hyperactivity disorder, BCS70 = the 1970 British Cohort Study, BHPS = British Household Panel Survey, GM = total grey matter volume, GPS = general population sample of Understanding Society, MCS = Millennium Cohort Study, MDD = major depressive disorder, NCDS = the 1958 National Child Development Study, Pt = p value threshold, WM = white matter.

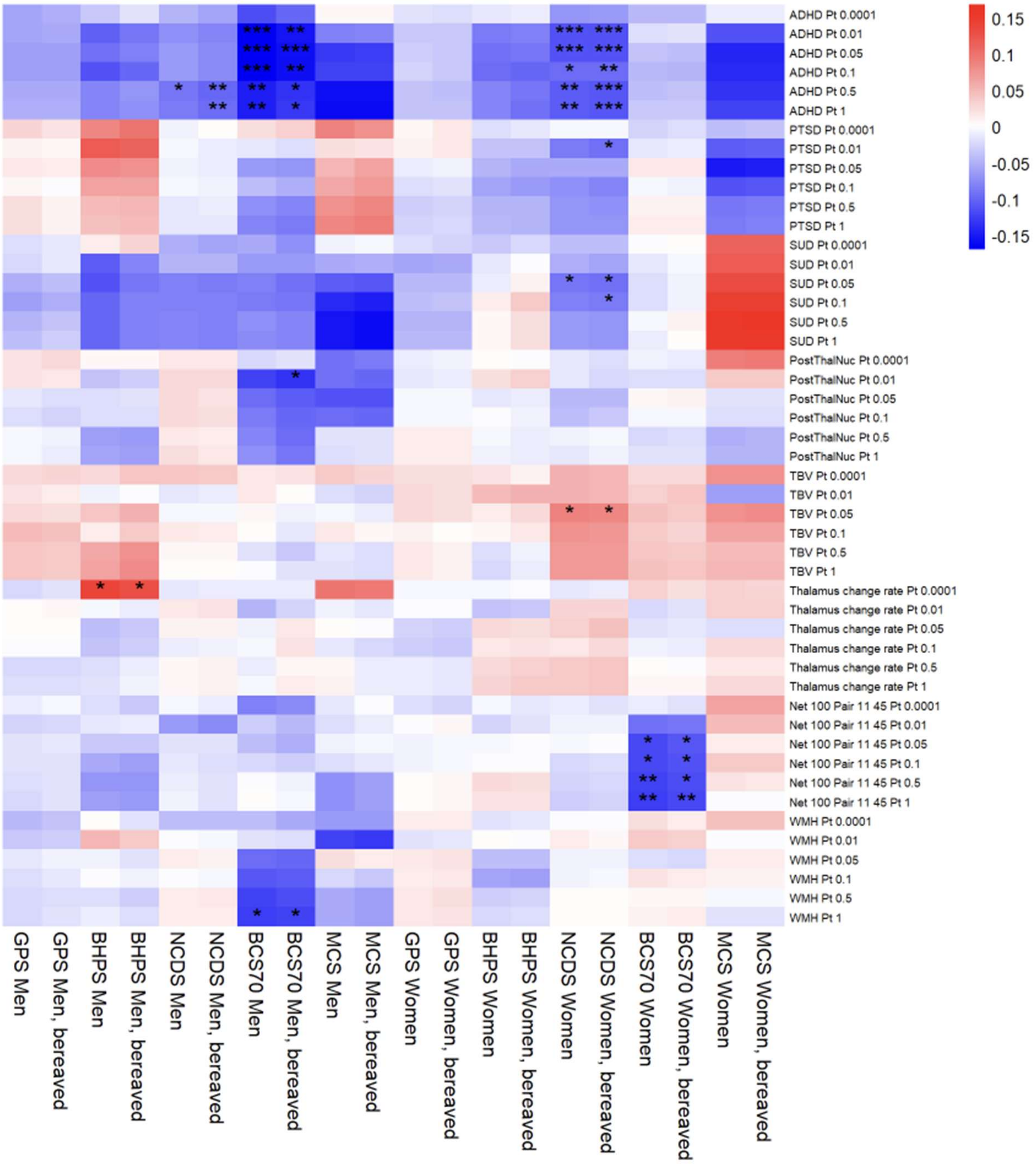


Supplementary figure 29. Associations between polygenic indices (PGIs) and mother educated. Sensitivity analysis results for analyses which include participants whose parent(s) passed away during youth. Main analysis results are also shown. PTSD PGI becomes associated in NCDS women. * = p < 0.05/41, ** = p < 0.01/41, *** = p < 0.001/41. ADHD = attention deficit hyperactivity disorder, BCS70 = the 1970 British Cohort Study, BHPS = British Household Panel Survey, GM = total grey matter volume, GPS = general population sample of Understanding Society, MCS = Millennium Cohort Study, NCDS = the 1958 National Child Development Study, PostThalNuc = posterior thalamic nuclei volume, Pt = p value threshold, PTSD = post-traumatic stress disorder, SUD = substance use disorders, TBV = total brain volume, WMH = white matter hyperintensities.

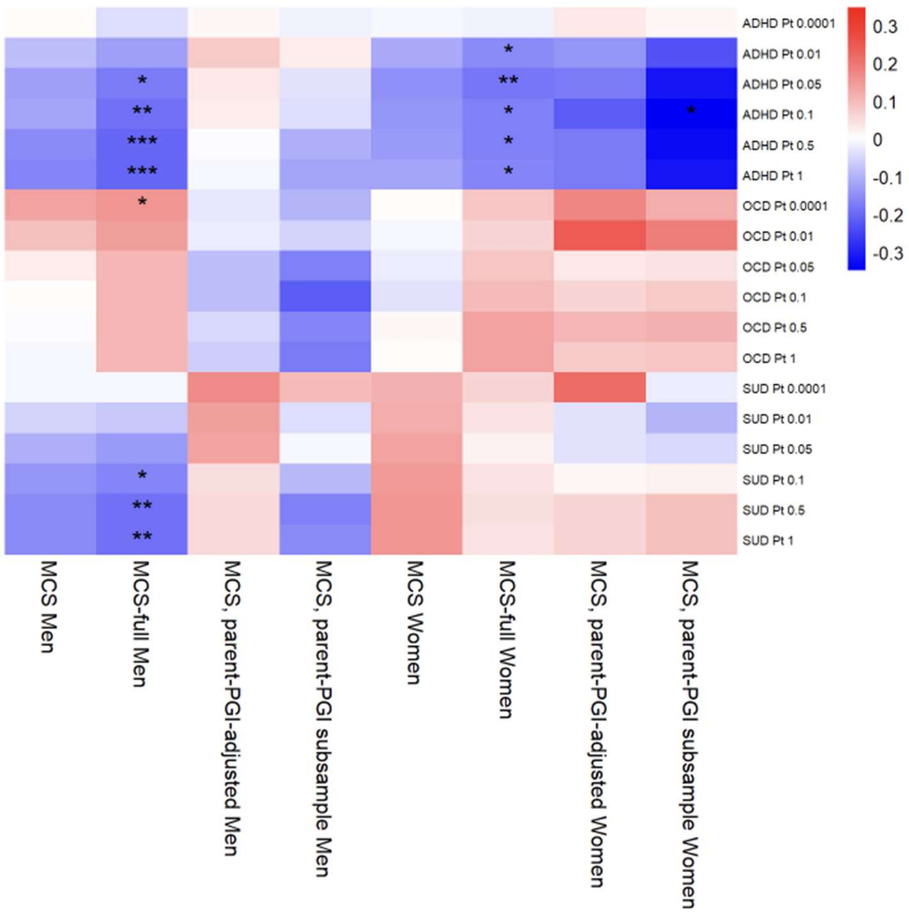


Supplementary figure 30. Associations between polygenic indices (PGIs) and mother educated. Results for MCS-specific sensitivity analyses, and MCS main analysis, are shown. Including all ancestries leads to greater rGE, while restricting to participants with parents' PGIs removes all rGE. * = p < 0.05/41, ** = p < 0.01/41, *** = p < 0.001/41. ADHD = attention deficit hyperactivity disorder, MCS = Millennium Cohort Study, OCD = obsessive-compulsive disorder, PGI = polygenic index, Pt = p value threshold, SUD = substance use disorders.

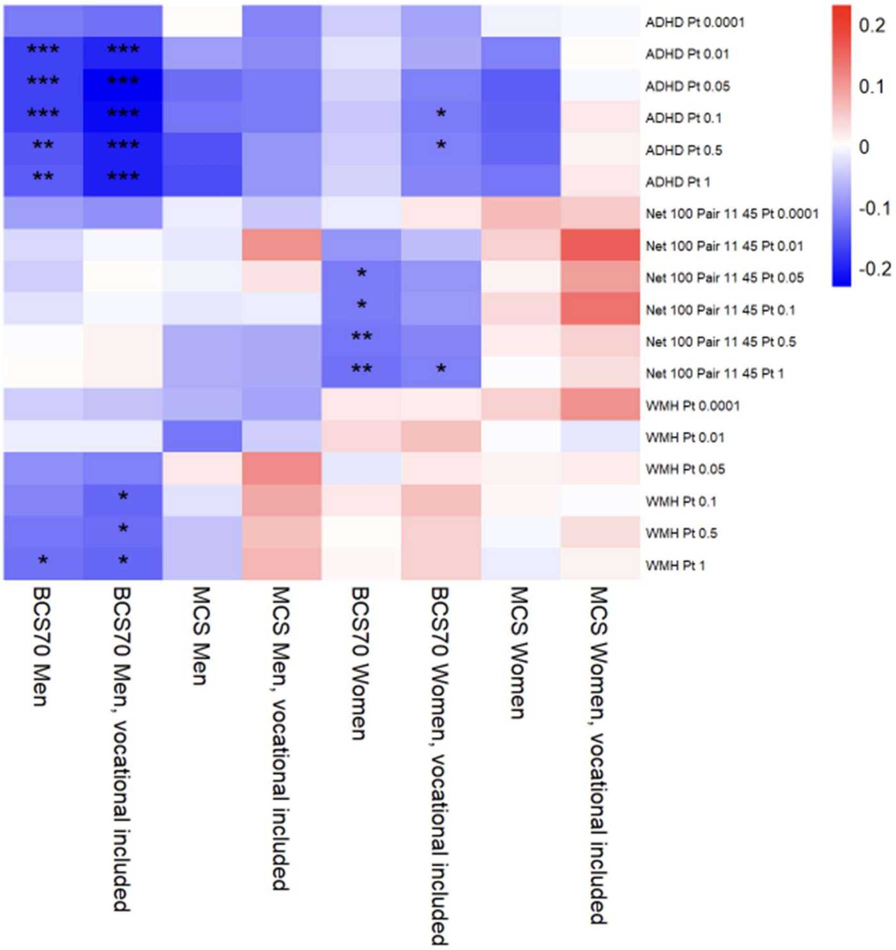


Supplementary figure 31. Associations between polygenic indices (PGIs) and mother educated. Sensitivity analysis results are shown for BCS70 and MCS using an alternative measure of mother's education which includes vocational qualifications. Main analysis results are also shown. * = p < 0.05/41, ** = p < 0.01/41, *** = p < 0.001/41. ADHD = attention deficit hyperactivity disorder, BCS70 = the 1970 British Cohort Study, MCS = Millennium Cohort Study, Pt = p value threshold, WMH = white matter hyperintensities.

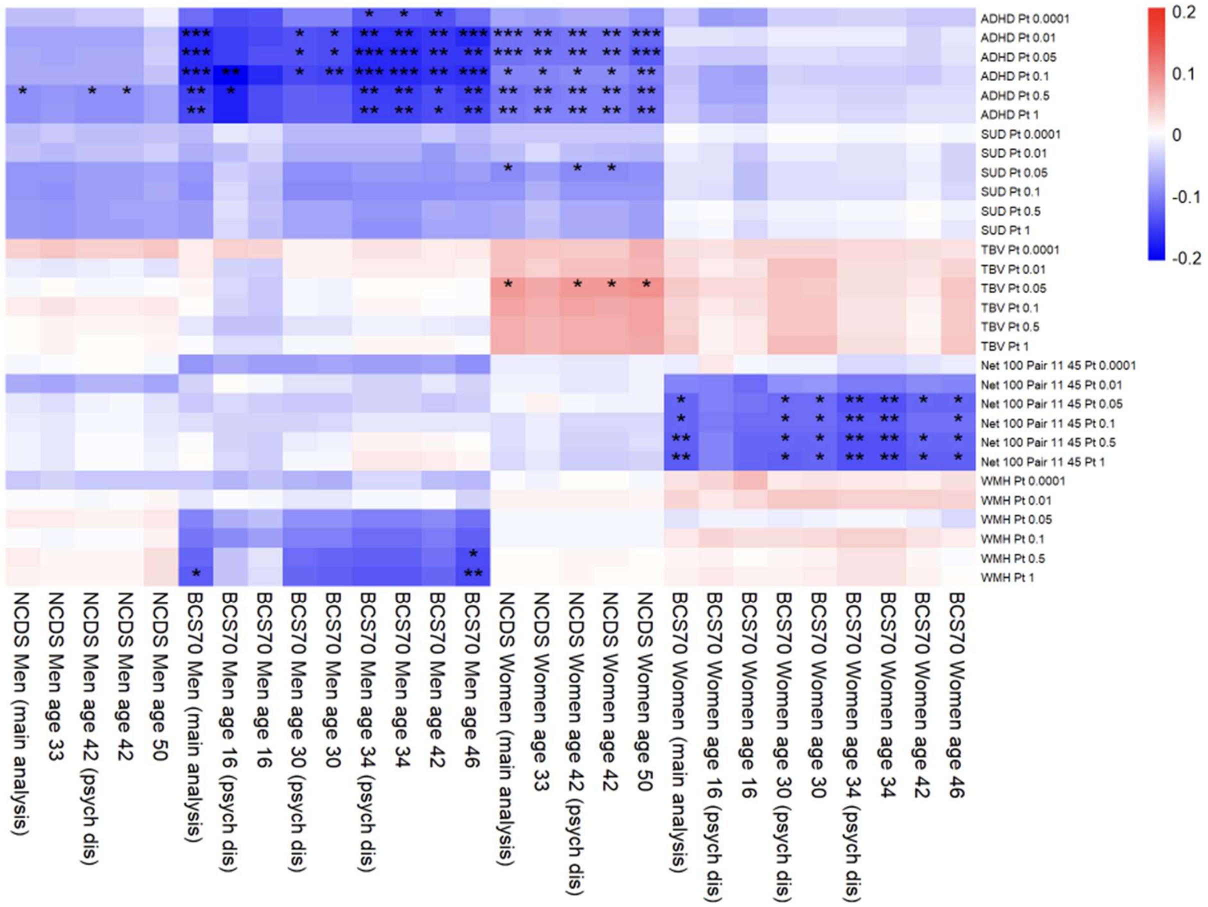


Supplementary figure 32. Associations between polygenic indices (PGIs) and mother educated. Sensitivity analysis results for NCDS and BCS70 using mental health outcomes at different ages. * = p < 0.05/41, ** = p < 0.01/41, *** = p < 0.001/41. ADHD = attention deficit hyperactivity disorder, BCS70 = the 1970 British Cohort Study, NCDS = the 1958 National Child Development Study, psych dis = psychological distress, Pt = p value threshold, SUD = substance use disorders, TBV = total brain volume, WMH = white matter hyperintensities.

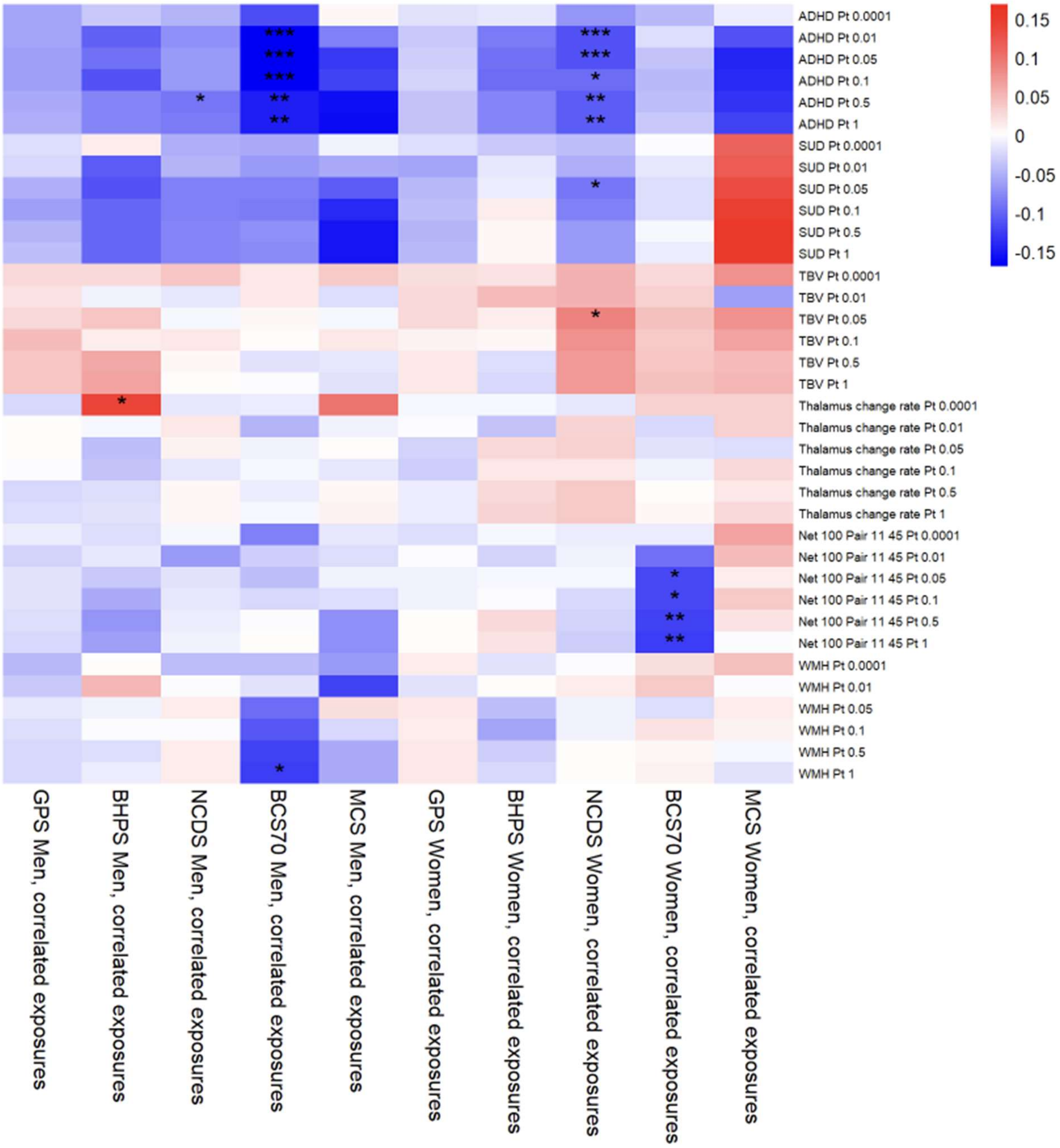


Supplementary figure 33. Associations between polygenic indices (PGIs) and mother educated. Sensitivity analysis results for path models with pairwise correlation terms for the three exposures. Results are identical to main analysis. * = p < 0.05/41, ** = p < 0.01/41, *** = p < 0.001/41. ADHD = attention deficit hyperactivity disorder, BCS70 = the 1970 British Cohort Study, BHPS = British Household Panel Survey, GPS = general population sample of Understanding Society, MCS = Millennium Cohort Study, NCDS = the 1958 National Child Development Study, Pt = p value threshold, SUD = substance use disorders, TBV = total brain volume, WMH = white matter hyperintensities.
